# Evaluating the utility of tumour mutational signatures for identifying hereditary colorectal cancer and polyposis syndrome carriers

**DOI:** 10.1101/2019.12.11.19014597

**Authors:** Peter Georgeson, Bernard J. Pope, Christophe Rosty, Mark Clendenning, Khalid Mahmood, Jihoon E. Joo, Romy Walker, Ryan Hutchinson, Susan Preston, Julia Como, Sharelle Joseland, Aung K. Win, Finlay A. Macrae, John L. Hopper, Dmitry Mouradov, Peter Gibbs, Oliver M. Sieber, Dylan E. O’Sullivan, Darren R. Brenner, Steve Gallinger, Mark A. Jenkins, Ingrid M. Winship, Daniel D. Buchanan

## Abstract

**Objective:** Germline pathogenic variants (PVs) in the DNA mismatch repair (MMR) genes and in the base excision repair gene *MUTYH* underlie hereditary colorectal cancer (CRC) and polyposis syndromes. We evaluated the robustness and discriminatory potential of tumour mutational signatures in CRCs for identifying germline PV carriers.

**Design:** Whole exome sequencing of formalin-fixed paraffin embedded (FFPE) CRC tissue was performed on 33 MMR germline PV carriers, 12 biallelic *MUTYH* germline PV carriers, 25 sporadic *MLH1* methylated MMR-deficient CRCs (MMRd controls) and 160 sporadic MMR-proficient CRCs (MMRp controls) and included 498 TCGA CRC tumours. COSMIC V3 single base substitution (SBS) and indel (ID) mutational signatures were assessed for their ability to differentiate CRCs that developed in carriers from non-carriers.

**Results:** The combination of mutational signatures SBS18 and SBS36 contributing >30% of a CRC’s signature profile was able to discriminate biallelic *MUTYH* carriers from all other non-carrier control CRCs with 100% accuracy (area under the curve (AUC) 1.0). SBS18 and SBS36 were associated with specific *MUTYH* variants p.Gly396Asp (p=0.025) and p.Tyr179Cys (p=5×10^−5^), respectively. The combination of ID2 and ID7 could discriminate the 33 MMR PV carrier CRCs from the MMRp control CRCs (AUC 0.99), however, SBS and ID signatures, alone or in combination, could not provide complete discrimination (AUC 0.79) between CRCs from MMR PV carriers and sporadic MMRd controls.

**Conclusion:** Assessment of SBS and ID signatures can discriminate CRCs from biallelic *MUTYH* carriers and MMR PV carriers from non-carriers with high accuracy, demonstrating utility as a potential diagnostic and variant classification tool.

**SIGNIFICANCE OF THE STUDY:** *What is already known about this subject?:*

- Identifying carriers of pathogenic variants (PVs) in moderate/high-risk colorectal cancer (CRC) and polyposis susceptibility genes has clinical relevance for diagnosis, targeted screening and prevention strategies, prognosis, and treatment options. However, challenges still remain in the identification of carriers and the classification of rare variants in these genes.
- Previous studies have identified tumour mutational signatures that result from defective DNA repair including DNA mismatch repair (MMR) deficiency and base excision repair defects, DNA repair mechanisms that underlie the common hereditary CRC and polyposis syndromes but their diagnostic utility in CRC is unknown.

*What are the new findings?:*

- Single base substitution (SBS)-related mutational signatures derived from whole exome sequencing of formalin-fixed paraffin embedded (FFPE)-derived CRC tissue DNA can effectively discriminate CRCs that developed in biallelic *MUTYH* PV carriers from CRC-affected non-carriers.
- CRCs that develop in MMR PV carriers (Lynch syndrome) can be effectively differentiated from sporadic MMR-proficient CRC by a combination of indel (ID) signatures, but the SBS and ID tumour mutational signatures are less effective at discriminating Lynch syndrome-related CRC from sporadic MMR-deficient CRC resulting from *MLH1* gene promoter hypermethylation.
- The SBS and ID mutational signatures associated with biallelic *MUTYH* PV carriers and MMR PV carriers are robust to changes in experimental settings.
- We demonstrate the optimal experimental settings for calculating mutational signatures and define thresholds that optimise sensitivity and specificity for classifying CRC aetiology as hereditary or non-hereditary.

*How might it impact on clinical practice in the foreseeable future?:*

- Deriving SBS- and ID-related mutational signatures from CRCs can identify carriers of PVs in hereditary CRC and polyposis susceptibility genes.
- The application of mutational signatures has the potential to improve the diagnosis of hereditary CRC and aid in variant classification, leading to improved clinical management and CRC prevention.

## INTRODUCTION

Colorectal cancer (CRC) is the third most commonly diagnosed cancer worldwide and is a leading cause of cancer-related morbidity and mortality [1]. Currently, 5-10% of CRCs develop in individuals who carry a pathogenic variant (PV) in a known hereditary CRC and/or polyposis susceptibility gene, including the DNA mismatch repair (MMR) genes (*MLH1, MSH2, MSH6, PMS2*) and the base excision repair (BER) gene *MUTYH* [2] (reviewed in [3]). Identifying carriers of PVs in these susceptibility genes has important implications for preventing subsequent primary cancers in the proband [4,5] and for the prevention of CRC in relatives through targeted screening approaches such as colonoscopy with polypectomy [6,7].

Currently, the most common strategy to identify MMR gene PV carriers (Lynch syndrome) starts with testing the tumour for microsatellite instability (MSI) and/or loss of MMR protein expression by immunohistochemistry [8]. However, loss of MMR protein expression (MMR-deficiency) in a CRC is not diagnostic for carrying a PV because MMR-deficiency can be caused by epigenetic inactivation of the MMR genes (hypermethylation of the *MLH1* gene promoter) or biallelic somatic mutations of MMR genes [9,10]. Moreover, no tumour-based approach is routinely applied to identify biallelic carriers in *MUTYH*; currently germline testing is guided by the presence of associated phenotypic features [3], although the phenotype for biallelic *MUTYH* carriers has been shown to be variable, making this approach suboptimal [11]. Furthermore, the transition of clinical genetics to multi-gene panel testing for identifying hereditary CRC and polyposis syndromes has led to further challenges with the classification of variants of uncertain clinical significance (VUS)[12].

The determination of tumour mutational signatures (TMS) is an emerging approach that integrates the somatic mutation landscape within a single tumour to identify patterns associated with distinct oncogenic pathways [13–16]. Most commonly, each TMS is derived from compositional changes of single base substitutions (SBS), indels (ID), and doublets. Version 3 of the predominant TMS framework published on the Catalogue of Somatic Mutations in Cancer (COSMIC) website defines 95 signatures, of which a proposed aetiology is available for 63 (66%) [17]. To date, multiple TMS have been identified that relate to defective DNA repair including MMR (SBS6, SBS14, SBS15, SBS20, SBS21, SBS26, SBS44, ID1, ID2, ID7), and BER defects caused by dysfunctional *NTHL1* (SBS30) [18] and *MUTYH* (SBS18 and SBS36) [19,20].

Although TMS show substantial promise in the translational setting, clinical adoption has been limited [21]. In this study, we evaluated the SBS and ID TMS landscapes in CRCs caused by germline PVs in the MMR genes or by biallelic PVs in the *MUTYH* gene, to test the clinical utility of TMS for predicting PVs in these specific genes. We assessed the effect of experimental settings and quantified the discriminatory potential of TMS for identifying PV carriers from non-carrier CRCs.

## MATERIALS AND METHODS

### Study Cohort

CRC-affected individuals with tumour and matched germline whole exome sequencing (WES) data from five studies were included in the analysis: the Australasian (ACCFR) and Ontario (OCCFR) sites of the Colon Cancer Family Registry [22,23], The ANGELS study, WEHI study and The Cancer Genome Atlas (TCGA) colon adenocarcinoma (COAD) and rectum adenocarcinoma (READ) study [24] (**Figure 1, Supplementary Table 1, Supplementary Table 2**). Formalin-fixed paraffin embedded (FFPE) CRCs from carriers of germline PVs in *MLH1, MSH2* or *MSH6* (n=33), from biallelic or monoallelic carriers of PVs in *MUTYH* (n=21) and from an individual carrying a PV and a VUS in *MUTYH* were included. Tumour MMR status was determined by immunohistochemistry with details of the tumour and germline characterisation undertaken described previously [25,26]. Two groups of FFPE CRCs were selected as non-hereditary controls: 1) MMR-proficient (MMRp) CRCs without a known germline mutation in the *MUTYH* or MMR genes were included as “MMRp controls” (n=160), and 2) MMR-deficient (MMRd) CRCs resulting from somatic *MLH1* gene promoter hypermethylation were included as “MMRd controls” (n=25). Tumours were then separated into discovery (n=142) and validation groups (n=97) based on their recruitment origin from either clinic-based (discovery) or population-based (validation). Furthermore, 498 CRC tumours from TCGA COAD and READ studies [24] were included as an additional set of fresh-frozen, non-hereditary CRCs (**Supplementary Table 2**). MMR status was determined using MSIseq [27], enabling stratification into 446 MMRp (“TCGA MMRp controls”) and 52 MMRd (“TCGA MMRd controls”), where each of the 52 MMRd controls were confirmed to have tumour hypermethylation of the *MLH1* gene promoter (**Supplementary Methods**).

**Table 1.**
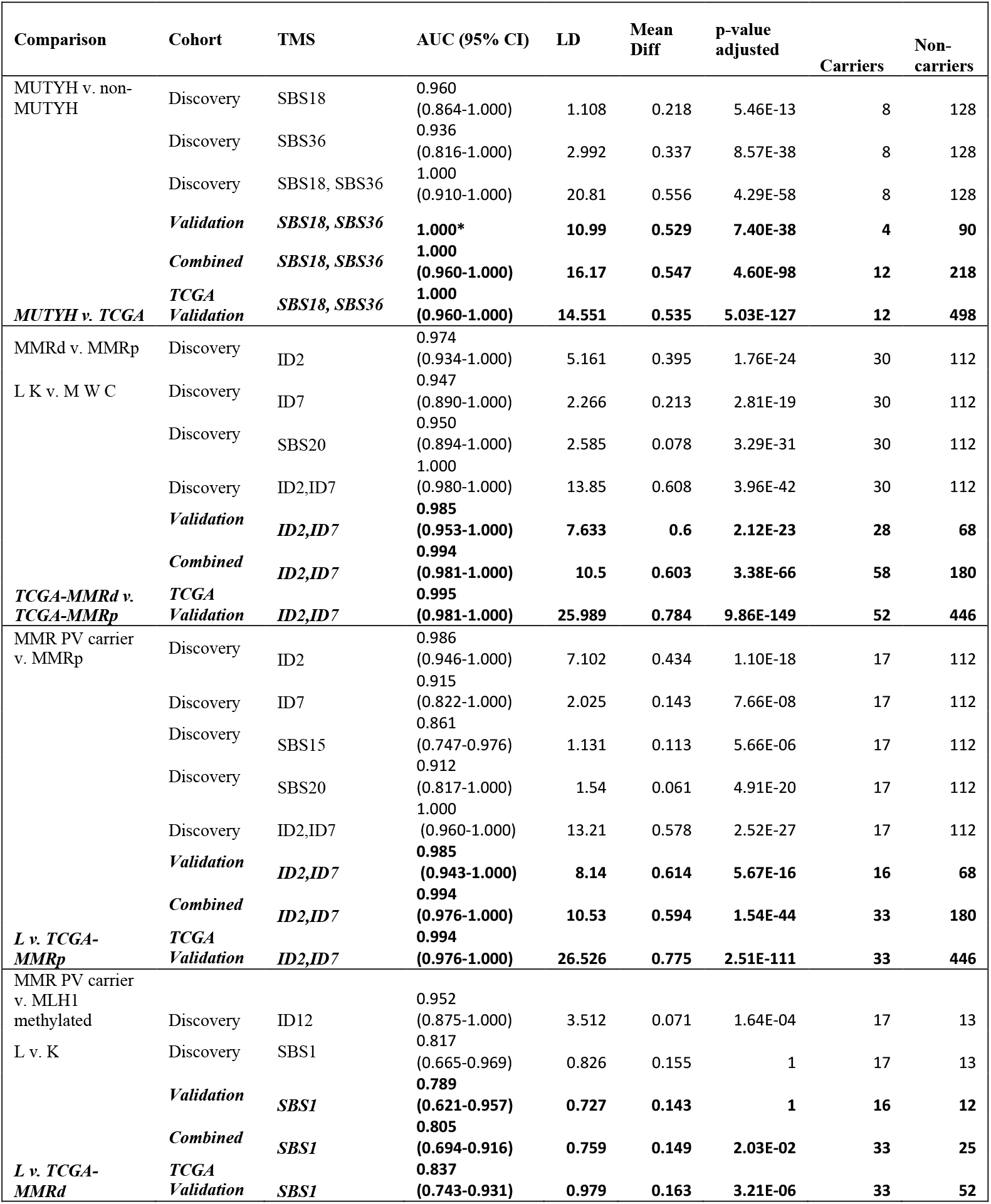
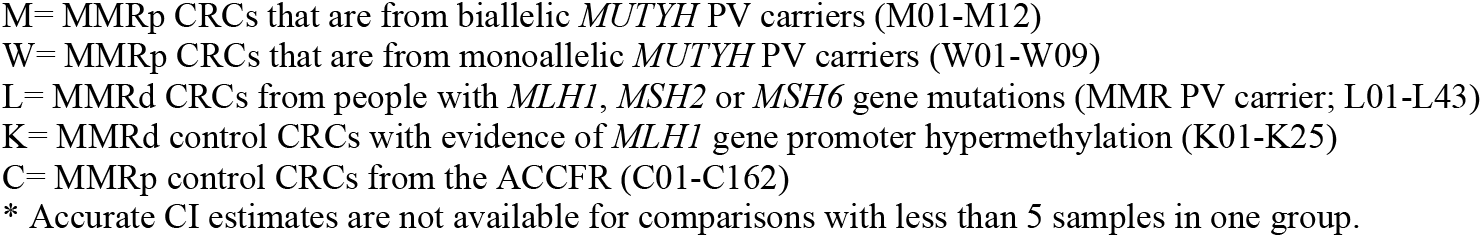
Individual tumour mutational signatures (TMS) identified to be associated with a hereditary CRC group from the analysis of 737 CRCs. Individual TMS with an AUC>0.90 and/or mean difference >10pp are shown, as well as the best combination of TMS determined using forward selection applied to the discovery group. The robustness of these results are illustrated by the accuracy obtained when applied to an independent validation set, and when our MMRp and MMRd control groups were replaced with TCGA MMRp and MMRd CRCs.

**Figure 1.**
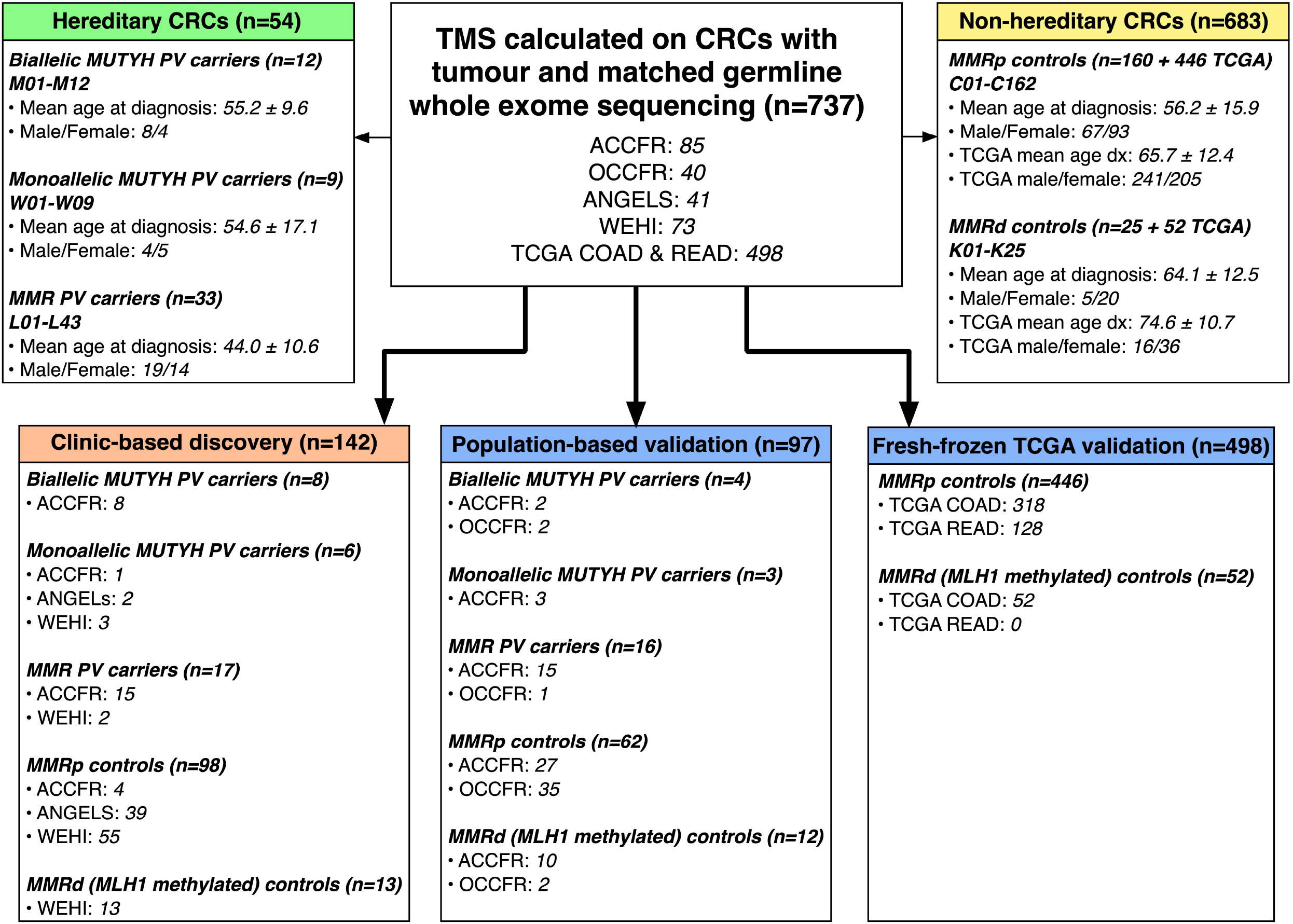
Breakdown of tumours, their male/female ratio and mean age at diagnosis (±SD) included in this study from the ACCFR, OCCFR, ANGELS, WEHI and TCGA studies. In total, 737 CRCs were analysed, consisting of 54 hereditary CRCs and 683 non-hereditary CRCs. A discovery group of 142 CRCs were selected from clinic-based studies, while the validation group of 97 CRCs was selected from population-based studies. An additional 498 tumours sourced from colorectal adenocarcinoma (COAD) and rectal adenocarcinoma (READ) studies of TCGA were included as an additional comparison group. Non-hereditary CRCs were stratified into mismatch repair-proficient (MMRp controls) and mismatch repair-deficient (MMRd controls) tumours.

### Whole Exome Sequencing and Analysis

FFPE CRC tissues were macrodissected and sequenced as tumour with matching peripheral blood-derived DNA sequenced as germline (**Supplementary Methods**). Somatic single-nucleotide variants (SNVs) and short insertion and deletions (indels) were called with Strelka 2.9.2 and used to calculate TMS using the simulated annealing method previously described by SignatureEstimation [28] from the set of COSMIC version 3 signatures [17] (**Supplementary Methods**). The impact of experimental settings was explored by filtering variants based on depth of coverage (DP) and the variant allele fraction (VAF) in the tumour, then calculating TMS at each filter point. Details of the TCGA analysis are provided in the **Supplementary Methods**.

We measured the separability of each hereditary CRC group from the non-hereditary CRC group, at each filtering setting, for each relevant TMS. We measured the difference in the means of the two groups, and statistical significance was determined using a one-sided t-test. To further discriminate groups with high (or perfect) area under the curve (AUC) of the receiver operator curve, we also calculated Fisher’s linear discriminant (LD) [29], which measures the distance between the means relative to the average standard deviation of the groups, which provides a quantitative measure of separability even when the groups have an AUC of 1.0 [30] (**Supplementary Methods**).

To find the optimal subset of TMS for identifying each hereditary group, we applied forward selection, an algorithm that is suitable for datasets with many potentially explanatory variables [31]. We maximised AUC and the mean difference between the groups, with stringent requirements to reduce the likelihood of overfitting. We adjusted and report p-values with Bonferroni correction applied [32] (**Supplementary Methods**).

### Patient and Public Involvement

There has been no patient or public involvement in the generation of this research report.

## RESULTS

### Overall TMS Results

The overview of all 737 CRCs analysed in this study and their breakdown by hereditary (n=54) and non-hereditary (n=683) subtype and distribution in the discovery (n=142) and validation (n=595) sets are shown in **Figure 1** and **Supplementary Table 3**. The hereditary CRC group were not significantly different in their age at diagnosis compared with the non-hereditary CRCs (48.2±12.6 years versus 51.5±16.9 years, p=0.19) but were significantly younger when compared with the 498 TCGA tumours (66.6±12.5 years, p=1×10^−22^). The characteristics of each individual and their CRC are provided in **Supplementary Table 1** and **Supplementary Table 2**.

The calculated SBS and ID TMS compositions for each of the 142 CRCs in the discovery set and for each of the 97 CRCs in the validation set are shown in **Figure 2** and **Supplementary Figure 1**, respectively. The mean±SD for each SBS and ID TMS were compared between the CRCs from either the biallelic *MUTYH* carriers or from the MMR PV carriers with the non-hereditary CRCs to identify hereditary CRC associated SBS or ID TMS (**Figure 3**).

**Figure 2.**
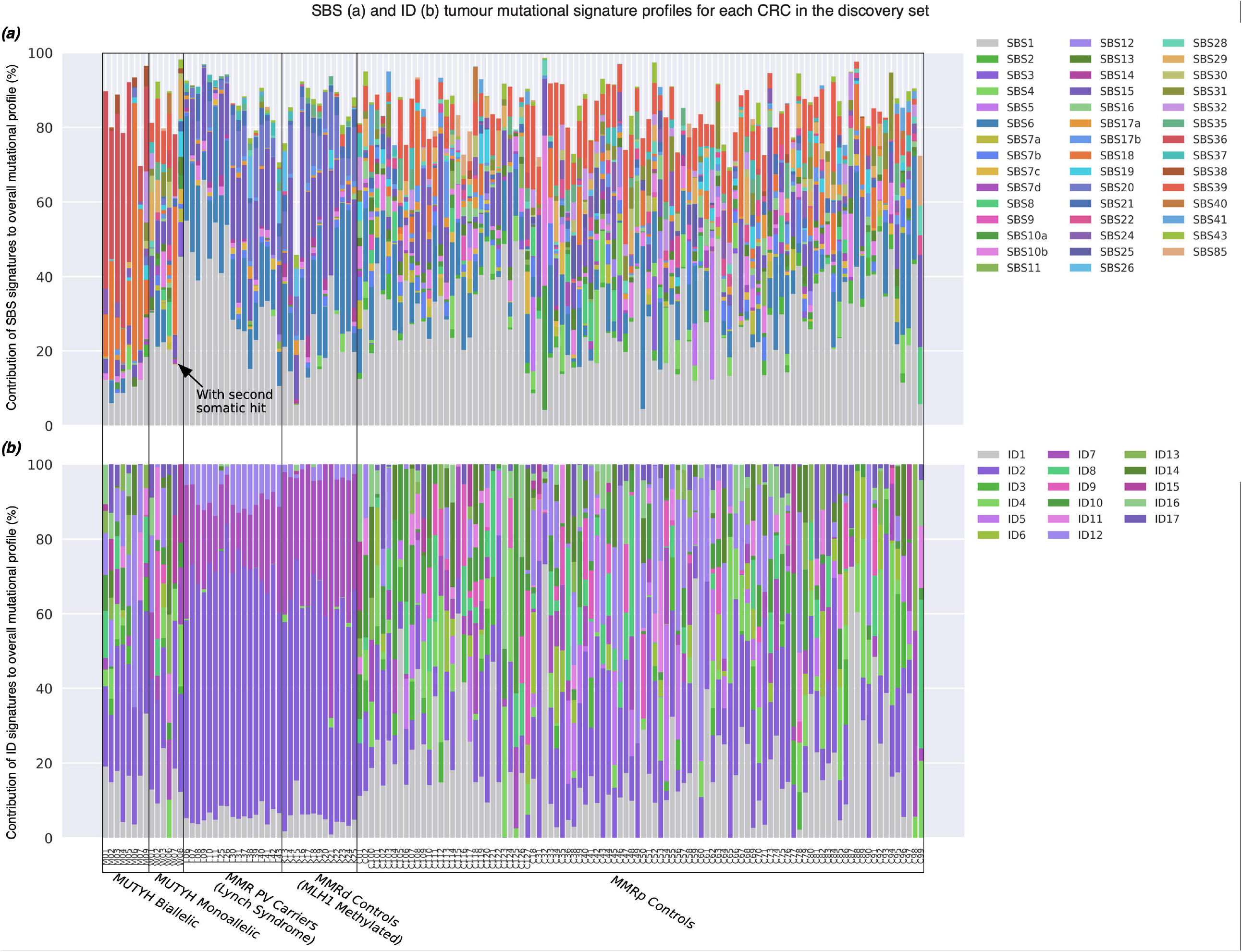
The tumour mutational signature (TMS) profiles based on the COSMIC v3 signature set for each of the 142 CRCs tested by whole exome sequencing and included in the discovery group: (a) Single base substitution (SBS)-derived TMS and (b) insertion and deletion (ID)-derived TMS profiles. CRCs were grouped by subtype: (i) biallelic *MUTYH* pathogenic variant carriers, (ii) monoallelic *MUTYH* pathogenic variant carriers, (iii) mismatch repair (MMR) gene pathogenic variant carriers (Lynch syndrome), (iv) MMR-deficient (MMRd) controls related to *MLH1* gene promoter hypermethylation and (v) MMR-proficient (MMRp) controls. Individual SBS or ID TMS with proportional values below 5% across all the CRC samples were excluded.

**Figure 3.**
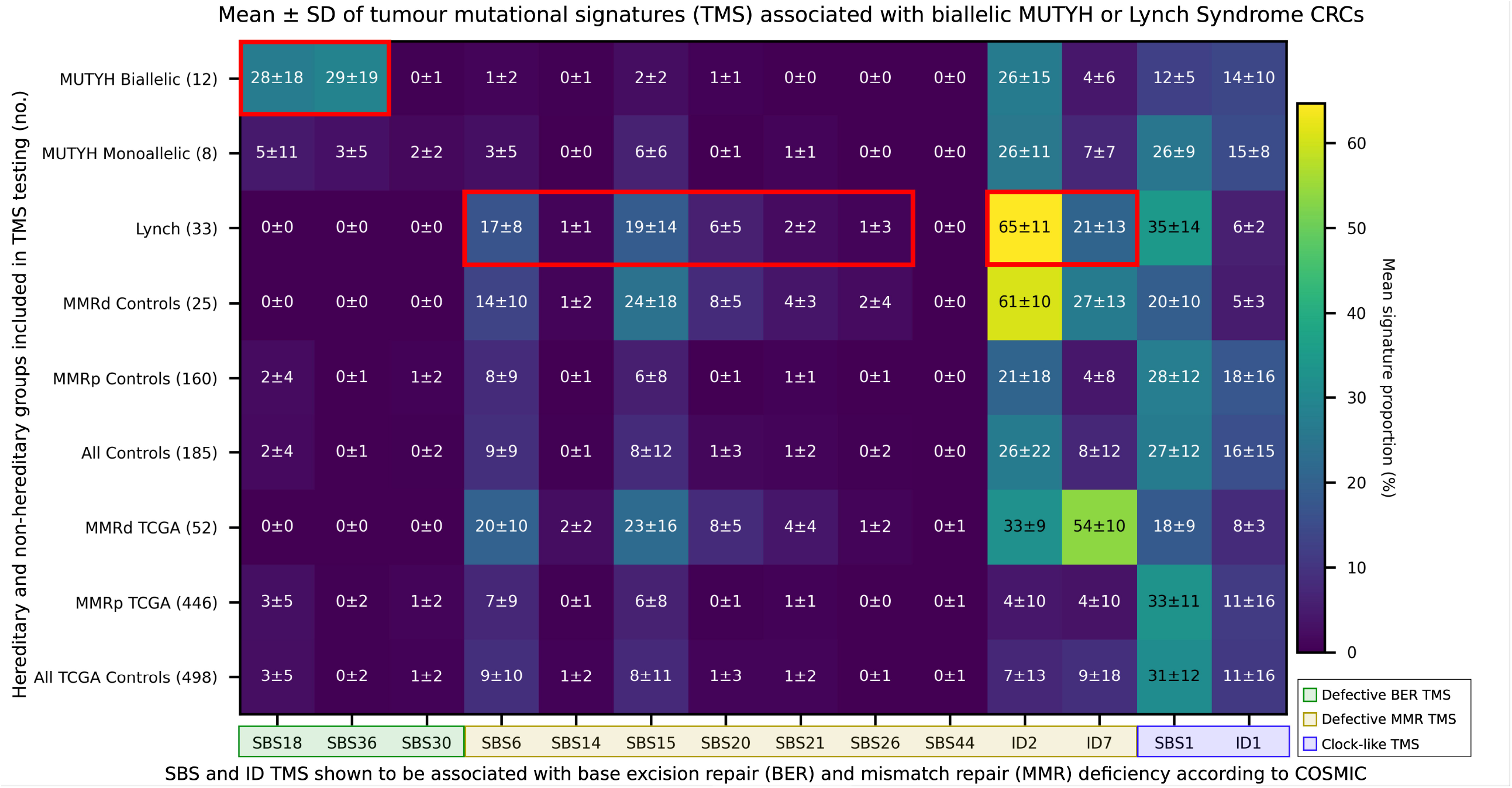
Heatmap showing the mean ± SD of each tumour mutational signature (TMS) in the hereditary and non-hereditary CRC groups included in the study. *X-axis*: COSMIC v3 TMS reported to be associated with defective base excision repair (BER; SBS18, SBS36, SBS30), with defective mismatch repair (MMR; SBS6, SBS14, SBS15, SBS20, SBS21, SBS26, SBS44, ID2, and ID7) and with ageing or “clock-like” signatures (SBS1 and ID1). *Y-axis*: hereditary CRC groups (biallelic *MUTYH* and monoallelic *MUTYH* pathogenic variant carriers and *MLH1, MSH2* and *MSH6* gene pathogenic variant carriers (Lynch)) and the non-hereditary and TCGA CRCs stratified by MMRd and MMRp controls. Red boxes highlight TMS signatures found in this study to be significantly associated with CRCs from biallelic *MUTYH* pathogenic variant carriers or with CRCs from Lynch syndrome carriers when compared with all CRC controls (n=185) or with MMRp control CRCs (n=160), respectively.

### Identifying biallelic *MUTYH* PV carrier TMS in CRC

For the 12 biallelic *MUTYH* carrier CRCs, SBS18 (mean±SD; 27.5±18.2%) and SBS36 (29.0±18.8%) were significantly enriched when compared with non-*MUTYH* CRCs (groups C, K and L) (1.6±3.9%; p=1×10^−34^ and 0.1±0.7%; p=2×10^−56^, respectively; **Figure 3**) and when compared with the TCGA tumours (2.7±5.0%; p=5×10^−42^ and 0.3±1.5%; p=2×10^−115^, respectively; **Figure 3**).

The SBS18 and SBS36 TMS differed by the underlying germline *MUTYH* PV. The CRCs from homozygous carriers of the c.536A>G p.Tyr179Cys PV (n=5) showed significantly higher proportions of SBS36 (p=5×10^−5^), while the CRCs from homozygous carriers of the c.1187G>A p.Gly396Asp variant (n=4) showed higher proportions of SBS18 (p=0.025) (**Figure 4a** and **Figure 4b**). SNV-derived mutational burden was higher in CRCs from biallelic *MUTYH* carriers compared with MMRp controls (p=1×10^−6^) (**Figure 4e**), but no association was observed for the indel-derived mutation burden (**Figure 4f**). When stratified by PV, homozygous or compound heterozygous carriers of the c.536A>G p.Tyr179Cys (n=8) showed a significantly higher SNV-derived mutational burden compared with the p.Gly396Asp homozygous carriers (8.98±2.34 versus 4.27±0.82 mutations/Mb; p=0.0051).

**Figure 4.**
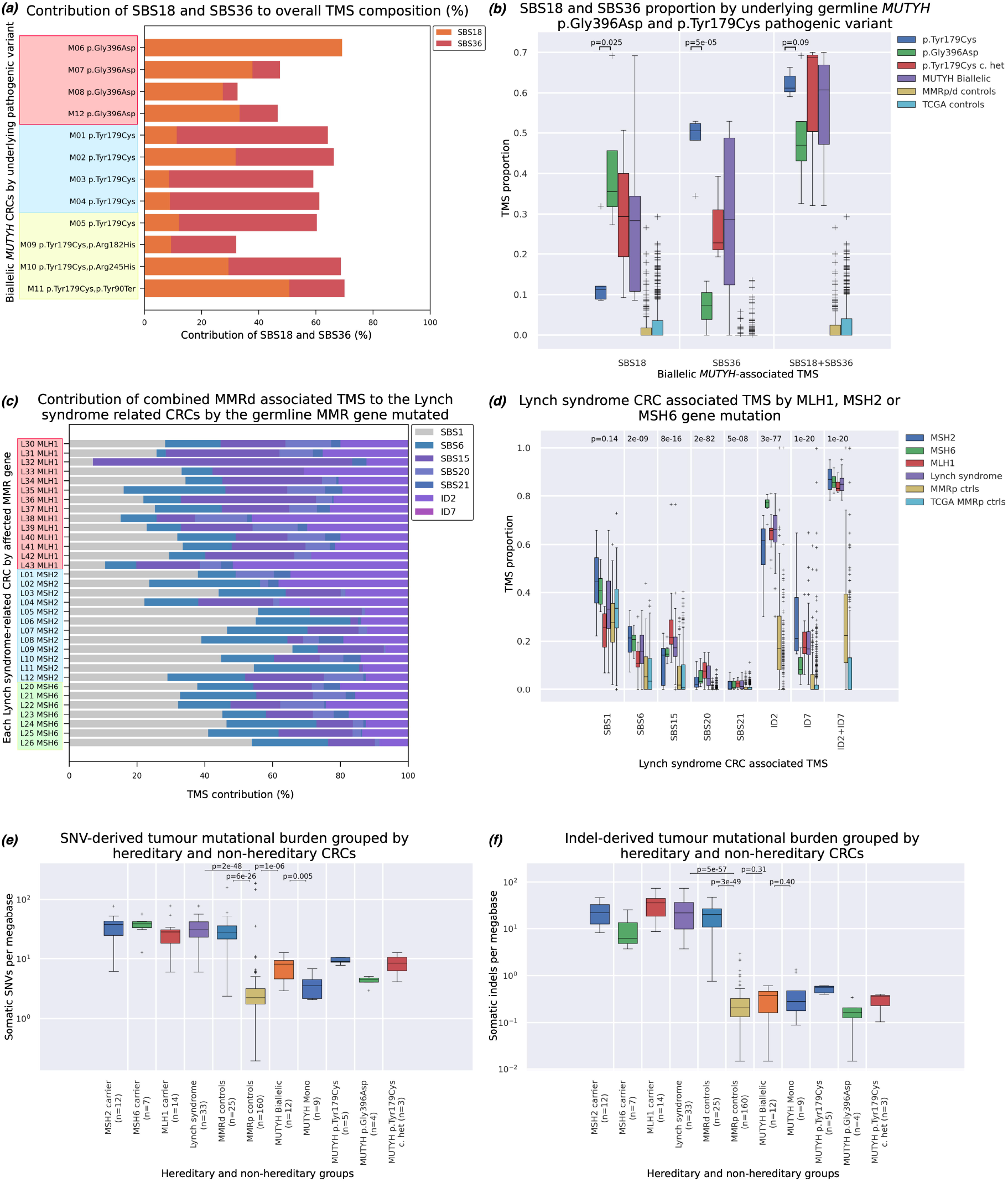
The investigation of *MUTYH* pathogenic variant (PV) and MMR gene specific tumour mutational signatures (TMS) profiles: **(a)** The contribution of the biallelic MUTYH-associated signatures SBS18 and SBS36 to each of the 12 biallelic *MUTYH* related CRCs by their underlying germline PVs: (i) homozygous p.Gly396Asp carriers, (ii) homozygous p.Tyr179Cys carriers and (iii) compound heterozygous p.Tyr179Cys carriers, **(b)** The mean±SD of SBS18, SBS36 and combined SBS18/SBS36 by underlying germline *MUTYH* pathogenic variant carrier group and CRC control groups. P-values indicate difference between CRCs from homozygous p.Gly396Asp carriers and homozygous p.Tyr179Cys carriers, **(c)** The contribution of the Lynch syndrome-associated TMS identified in our study for each of the 33 CRCs from the *MLH1, MSH2* and *MSH6* gene pathogenic variant carriers, **(d)** The distribution of each of the Lynch syndrome-associated TMS across the 14 CRCs from *MLH1* carriers, 12 CRCs from *MSH2* carriers, and 7 CRCs from the *MSH6* carriers and controls groups. P-values indicate differences between the six groups for each individual TMS as measured by ANOVA. **(e)** Somatic single nucleotide variant (SNV)-derived tumour mutational burden for hereditary and non-hereditary CRCs. CRCs from both biallelic *MUTYH* carriers and Lynch syndrome carriers exhibited significantly higher SNV-derived mutational burden compared with CRCs from MMRp controls (p=1.1×10^−6^ and p=2×10^−48^, respectively; excluding three ultra-hypermutated CRCs related to somatic mutations within the exonuclease domain of *POLE*), **(f)** Somatic insertion and deletion (Indel)-derived tumour mutational burden for hereditary and non-hereditary CRCs. CRCs from Lynch syndrome carriers exhibited significantly higher Indel-derived mutational burden compared with CRCs from MMRp controls (p=5×10^−57^, while CRCs from biallelic *MUTYH* carriers were not found to have a significant difference in Indel-derived mutational burden compared with CRCs from MMRp controls (p=0.31).

We investigated the ability of TMS to differentiate CRCs from biallelic *MUTYH* carriers from non-carriers by training a classifier using forward selection on the discovery set (n=136) and applying this to the validation sets of FFPE CRCs (n=94) and fresh-frozen CRCs from TCGA (n=498). The combination of SBS18 and SBS36 provided the most effective discrimination (AUC 1.0) and highest difference between TMS means (55.6 percentage points(pp)) of CRCs from biallelic *MUTYH* carriers from non-carriers in the discovery set (**Table 1**). The combination of SBS18 and SBS36 was similarly effective in discriminating the four *MUTYH* biallelic CRCs in the validation and TCGA set each with an AUC of 1.0 (**Table 1**). The sum of SBS18 and SBS36 accounted for, on average, more than half of the total signature composition for all 12 biallelic *MUTYH* CRCs (56.5±13.6%; range 32.1% to 70.0%) but had a negligible contribution to all other CRCs in the study (Non-MUTYH carrier CRCs: 1.8±3.9%; range 0.0% to 26.6%, p=5×10^−98^; TCGA: 3.0±5.2%; range 0.0% to 29.3%, p=9×10^−130^; **Figure 4b**).

We found that the robustness of TMS was dependent on different minimum VAF and minimum DP variant filtering settings. A minimum VAF threshold between 5% and 20% (**Figure 5a** and **Figure 5c**), and a minimum DP threshold of at least 30 reads (**Figure 5b** and **Figure 5d)** effectively separated biallelic *MUTYH* carrier CRCs from non-carrier CRCs. A minimum VAF of 10% and minimum DP of 50 reads were selected to maximise the capacity of TMS to identify CRCs from biallelic *MUTYH* carriers. Our analysis of the discovery set predicts that a CRC with a combined SBS18 and SBS36 proportion of >34% has a >95% likelihood of having biallelic *MUTYH* (**Figure 5c** and **Figure 5d**). Applied to the validation set, the 95% likelihood threshold achieved 100% specificity, but marginally misclassified one of the four biallelic carrier CRCs (M09; SBS18/SBS36 observed signature profile 32.6%). With all 12 biallelic carriers combined, 100% accuracy could be achieved with a threshold of 30% (**Supplementary Table 4**).

**Figure 5.**
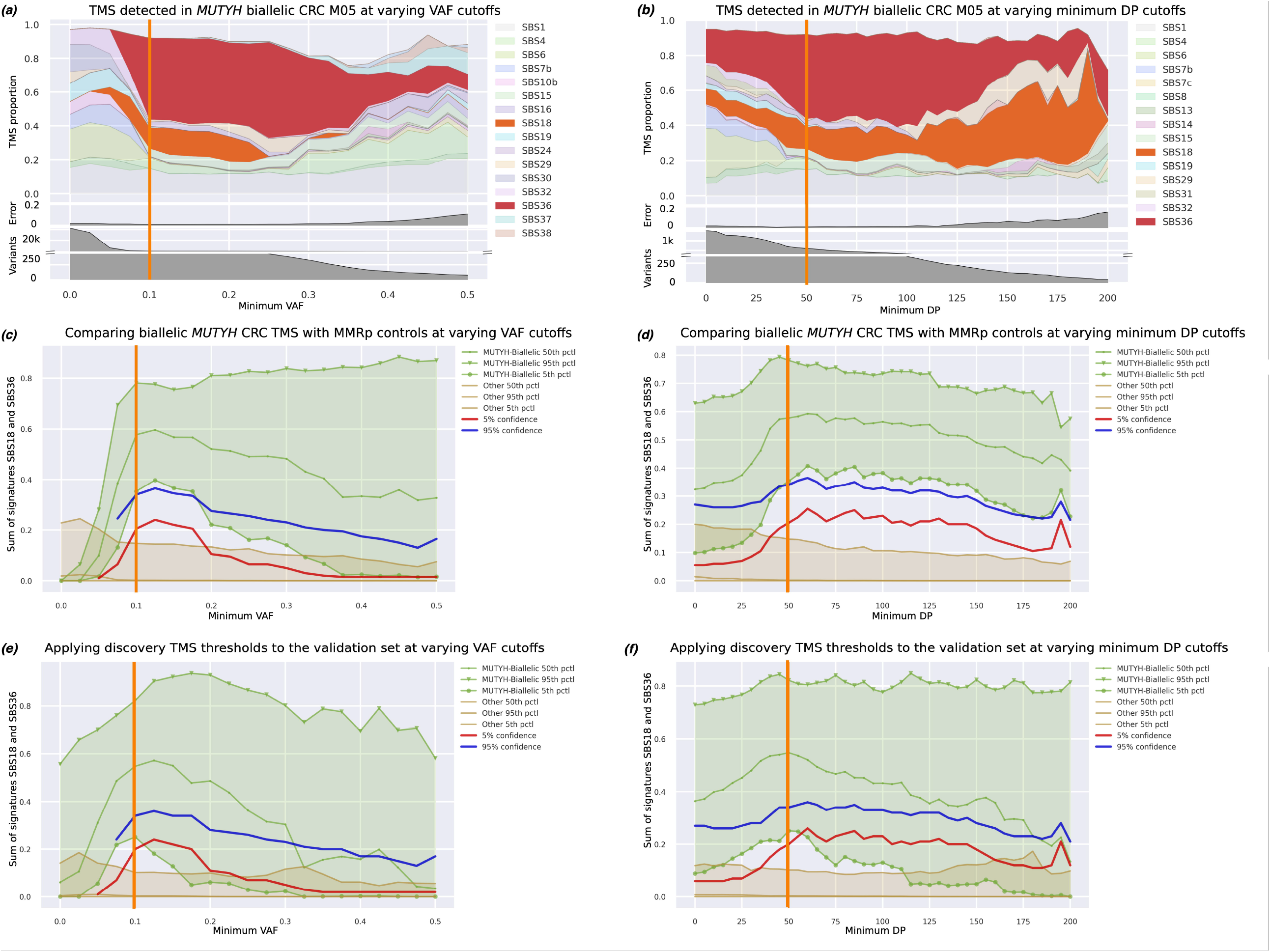
Tumour mutational signature (TMS) assessment for biallelic *MUTYH*-carrier CRCs. A representative CRC from a biallelic *MUTYH*-carrier (M05) was selected to demonstrate the changes in the SBS18 and SBS36 TMS profile related to changes in experimental conditions namely, **(a)**, changes in variant allele fraction (VAF) filtering (at a minimum read depth (DP) fixed at 50bp), and (b) changes in minimum DP (at a VAF fixed at 0.1). The SBS18 and SBS36 TMS were robust and dominant with VAF thresholds between 0.075 and 0.25 **(a)**, and with DP thresholds between 25-100bp **(b)** across all 12 biallelic *MUTYH* carrier CRCs (images not shown), with the upper thresholds of VAF and DP corresponding to limits where the number of somatic variants started to decline and the TMS error increased. The sum of SBS18 and SBS36 separated CRCs from biallelic *MUTYH* carriers (light green) from CRCs from the MMRp and MMRd controls (brown) from the discovery group when the VAF was between 0.075 and 0.175 **(c)** and when DP was >25 **(d)**. The calculated 95% (blue line), and 5% (red line) probabilities reflect the likelihood that a CRC is from a biallelic *MUTYH* pathogenic variant carrier. When applied to the validation set of CRCs, the sum of SBS18 and SBS36 similarly provided the best separation of CRCs from biallelic *MUTYH* carriers from the MMRp and MMRd control CRCs at VAF thresholds between 0.075 and 0.15 **(e)** and at DP thresholds between 25bp and 100bp **(f)**.

We applied these thresholds in two scenarios: 1) evaluating monoallelic *MUTYH* PV carriers, and 2) classifying a VUS. The sum of SBS18 and SBS36 was evaluated in eight monoallelic *MUTYH* PV carriers (W01-W08): with the exception of W07, the combined SBS18/SBS36 TMS was not observed (1.7±2.9%; range 0.0 to 7.6%), suggesting that monoallelic *MUTYH* PVs do not alone result in BER deficiency. No identifiable second somatic “hits” (SNV, indel or loss of heterozygosity) in *MUTYH* were evident in these seven monoallelic CRCs, however W07 exhibited combined SBS18/SBS36 of 51%, highly indicative of biallelic inactivation of *MUTYH* in this CRC. In addition to the germline NM_001128425.2:c.504+19_504+31del PV (ClinVar [33]), a somatic loss of heterozygosity event spanning the *MUTYH* gene was subsequently identified in W07 confirming biallelic *MUTYH* inactivation (**Supplementary Figure 2**).

We then applied our combined SBS18/SBS36 TMS threshold to a CRC (W09) from a person who carried a heterozygous *MUTYH* c.1187G>A p.Gly396Asp PV and a heterozygous *MUTYH* c.912C>G p.Ser304Arg variant classified as a VUS in ClinVar [33]. Both SBS18 and SBS36 were 0% and, therefore, there is a <1% likelihood that this CRC has biallelic *MUTYH* inactivation (**Figure 5c** and **Figure 5d**). This suggests that the c.912C>G p.Ser304Arg variant is likely to be benign or is *in cis* with the c.1187G>A p.Gly396Asp PV.

### Identifying a MMR gene carrier TMS in CRC

We investigated the utility of TMS related to defective MMR for identifying MMR PV carriers from both MMRp CRCs and sporadic MMRd CRCs. The MMR PV carriers (L01-L43) showed significantly higher levels of SBS6, SBS15, SBS20, ID2 and ID7 (p=1×10^−5^, 4×10^−11^, 2×10^−32^, 4×10^−28^, p=1×10^−14^ respectively) compared with the non-hereditary MMRp control CRCs (**Figure 3**). When comparing the MMR PV carriers to the MMRd control group of CRCs (K01-K25), only SBS1 showed a significant difference between the groups (34.6±13.7% v. 19.7±10.3%, p=0.020) (**Figure 3**). TMS of interest were found to be consistent at VAF thresholds between 0.00 and 0.15, and DP thresholds between 10 and 150 (**Figure 6, 7**).

**Figure 6.**
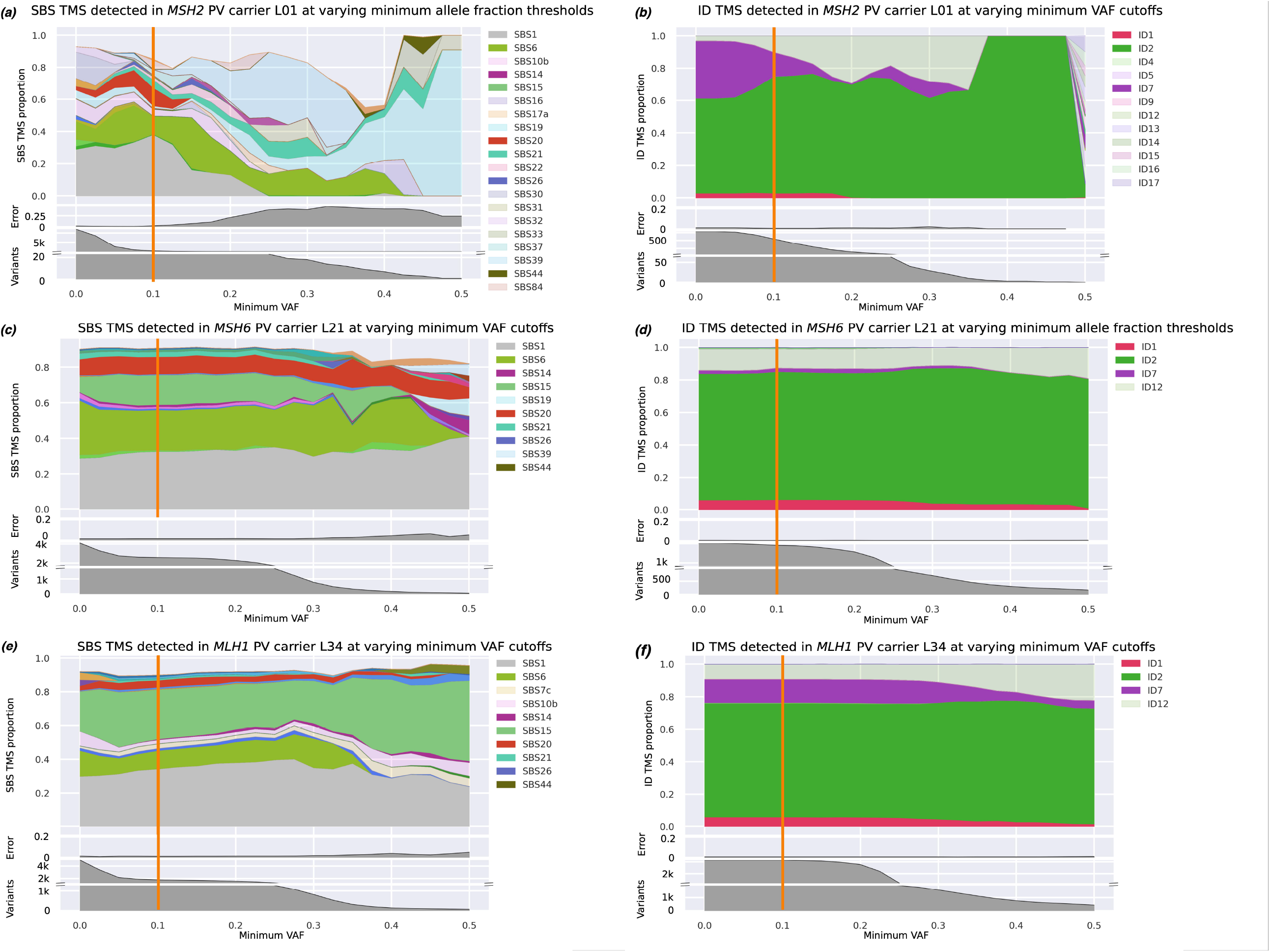
Assessment of single base substitution (SBS) and indel (ID) tumour mutational signatures (TMS) while varying the VAF threshold, for samples L01 **(a, b)**, L21 **(c, d)**, and L34 **(e, f)**, representing a CRC from a carrier of a germline pathogenic variant in the *MSH2, MSH6*, and *MLH1* genes, respectively. In all cases, ID signatures that have previously been associated with MMRd dominate at most VAF thresholds **(b, d, f)**, while relevant SBS signatures are also present but less dominant, particularly at highly stringent settings **(a, c, e)**.

**Figure 7.**
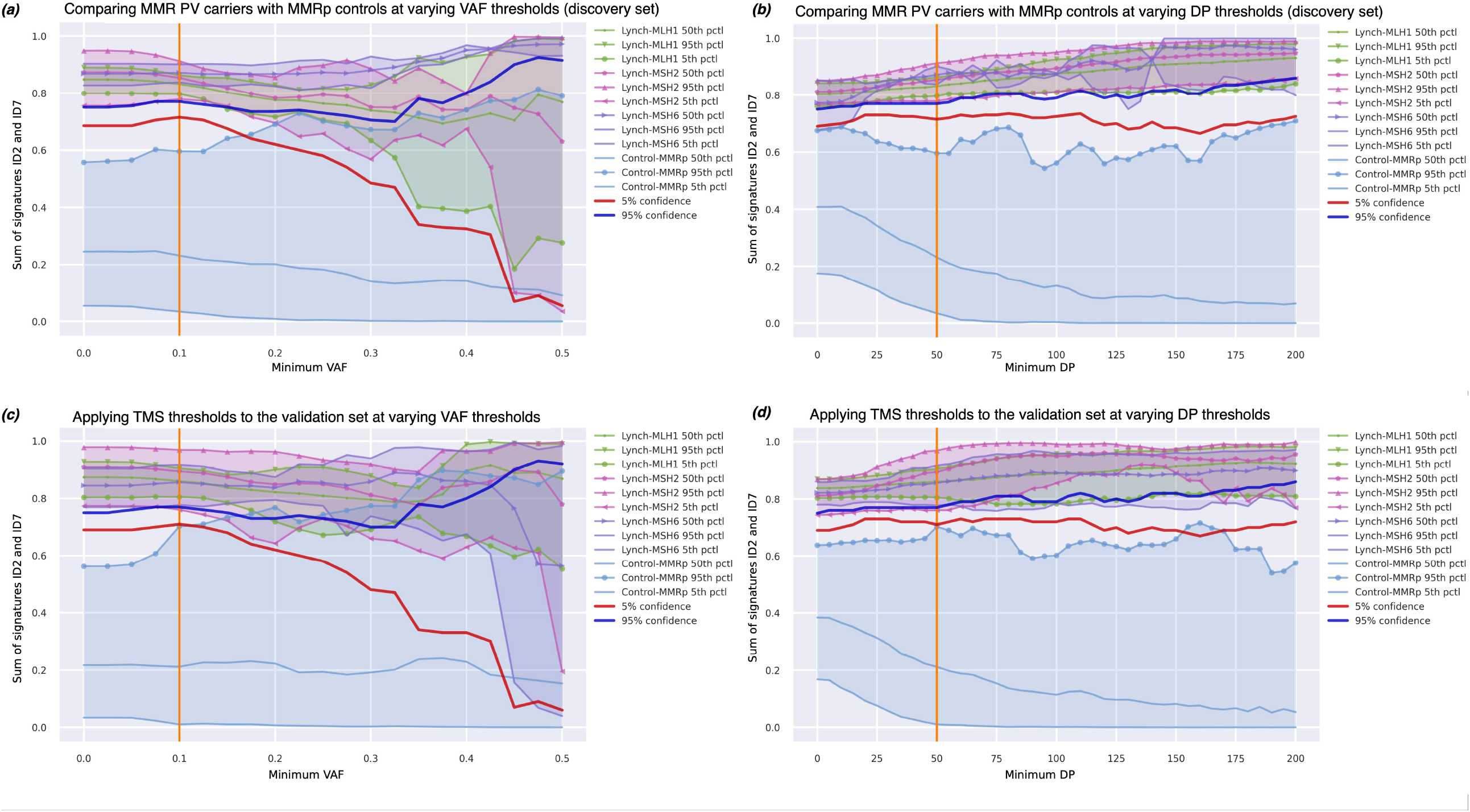
The sum of ID2 and ID7 effectively separated CRCs from MMR pathogenic variant carriers (n=17) from the MMRp control CRCs (n=98) at low somatic VAF thresholds (<0.2) **(a)** and across a broad range of minimum DP thresholds (>25bp) **(b)** when applied to the CRCs in the discovery set. When applied to the validation set of CRCs, we similarly observe a combined ID2 and ID7 TMS could effectively discriminate CRCs from MMR pathogenic variant carriers from CRCs from the MMRp controls at low VAF thresholds (≤0.1) **(c)** and across broad range of minimum DP thresholds **(d)**.

Determining MMRd and MMRp CRCs is a common molecular stratification in the diagnosis and treatment of CRC. We applied the forward selection classifier to discriminate MMRd CRC, comprising both Lynch- and *MLH1* methylated-CRCs (n=30) from MMRp CRCs (n=112) in the discovery group. This identified ID2 and ID7 as the most informative combination of TMS, achieving an AUC of 1.0, and difference of 60.8pp between the means of the groups and a high degree of replication in the validation and TCGA tumour groups (**Table 1**). The mean number of indel mutations in the MMRd and MMRp CRCs from both discovery and validation CRCs combined was significantly different (23.0±14.7 versus 0.29±0.35; p=6×10^−50^; **Figure 4f**).

We then applied forward selection to discriminate MMR PV carriers (n=17) from the MMRp CRCs in the discovery group, also identifying ID2 and ID7 as the most informative combination of TMS, achieving an AUC of 1.0, and difference of 57.8pp between the means of the groups (**Table 1, Figure 4d**), a result that showed a high degree of replication in the validation and TCGA MMRp control groups (**Table 1, Figure 4d**). When considering the sum of ID2 and ID7, the likelihood of a MMR PV carrier relative to a MMRp CRC was >95% at values >77% (**Figure 7a** and **Figure 7b**). Applied to the validation set, the sum of ID2 and ID7 yielded an AUC of 0.986 (**Table 1**). The 95% threshold exhibited 100% sensitivity and 98.8% specificity (**Supplementary Table 5**). Two MMRp tumours incorrectly classified as MMRd – C31 and C150 – had TMS sums of 74% and 100% respectively. We found no evidence of a germline MMR gene PV in these two people. Both tumours did, however, exhibit low indel counts: 4 and 1 respectively, which increases the uncertainty in calculated TMS (high reconstruction error) as has previously been noted [34,35], therefore, these two MMRp CRCs are likely false positives.

A common diagnostic challenge is to differentiate hereditary (Lynch syndrome) from sporadic (*MLH1* methylated) MMRd CRC. Forward selection showed no significant TMS in the discovery group (n=30), but when applied to the full dataset (n=58), SBS1 was the best performing and only significant TMS (p=0.02) that could discriminate MMR PV carriers (L01-L43) from the MMRd control CRCs (K01-K25). However, SBS1 only achieved an AUC of 0.805 (**Table 1**). This result improved marginally (AUC 0.837) when a larger group of TCGA MMRd controls (n=52) were utilised (**Table 1**). Consequently, the MMR PV carriers and sporadic MMRd CRCs could not be separated with high confidence.

## DISCUSSION

Germline PVs in the DNA MMR genes and biallelic PVs in *MUTYH* result in a high risk of developing CRC as well as extra-colonic cancers [6,7,36], therefore, the identification of carriers is of great importance for cancer prevention [18,37]. In this study, we demonstrated the utility of a combined SBS18 and SBS36 TMS for discriminating CRCs from biallelic *MUTYH* carriers from non-carriers and the utility of a combined ID2 and ID7 TMS for discriminating both MMRd and MMR PV carriers from MMRp CRCs. Our analysis showed that a combined SBS18 and SBS36 TMS proportion threshold of 30% provided 100% sensitivity and specificity for identification of CRCs from biallelic *MUTYH* carriers in both discovery and validation sets in this study. Although the sum of SBS18 and SBS36 was more effective than each of these signatures alone in being able to differentiate biallelic *MUTYH* carriers from non-carriers, associations between individual TMS and PV type were observed, namely the p.Tyr179Cys PV and SBS36 and p.Gly396Asp and SBS18. CRCs that developed in MMR PV carriers could be effectively differentiated from non-hereditary MMRp CRCs by a combination of TMS derived from somatic indel mutations (ID2 and ID7), achieving 100% sensitivity and 99.4% specificity in the pooled discovery and validation sets when the combined ID2 and ID7 TMS proportion threshold was 75%. TMS were less effective at discriminating MMR PV carriers from sporadic MMR-deficient CRCs resulting from *MLH1* gene promoter hypermethylation (SBS1 validation set AUC: 0.789). The findings for a combined SBS18 and SBS36 TMS and for a combined ID2 and ID7 TMS were consistent when assessed against the TCGA MMRp and MMRd fresh-frozen tumours, highlighting both the robustness and reproducibility of these findings and supporting the utility of deriving TMS from CRCs to identify carriers of PVs in *MUTYH* and MMR genes, respectively.

SBS18 and SBS36 were individually associated with CRCs from biallelic *MUTYH* carriers in our cohort, but neither TMS alone completely separated the carriers from the non-*MUTYH* CRCs (AUC 0.981 and 0.956 respectively). Combining SBS18 and SBS36 resulted in complete separation from the non-*MUTYH* carrier CRCs. This was also observed for the MMR PV carriers where the combination of ID2 and ID7 resulted in improved separation from the MMRp CRCs in the validation set (combined ID2 and ID7 AUC 0.986 v. 0.939 (ID2) and 0.922 (ID7)), highlighting the benefit of combining TMS for improved discrimination.

This study confirms previous reports of the association of SBS18 and SBS36 with CRCs from biallelic *MUTYH* carriers [19,20], however we have demonstrated several novel findings in our study: We 1) showed the effectiveness of a combined SBS18/SBS36 TMS profile for identifying biallelic *MUTYH* carriers, 2) quantified the capacity of combined SBS18/SBS36 TMS of 30% which could identify *MUTYH* biallelic carriers with 100% accuracy, 3) identified a significant association between germline *MUTYH* PV carrier CRCs and individual SBS18 and/or SBS36 TMS, 4) observed a significantly elevated SNV-derived mutational burden in CRCs from homozygous p.Tyr179Cys PV carriers relative to the homozygous p.Gly396Asp PV carriers (p=0.0051) and to the MMRp controls (excluding 3 ultra-hypermutated CRCs each with a pathogenic *POLE* exonuclease domain somatic mutations, p=1×10^−6^), and 5) provided an example of the application of the SBS18/SBS36 TMS for variant classification.

Monoallelic *MUTYH* PV carriers are reported to have a small but significant increased risk of CRC [25,38]. A previous study identified high levels of SBS18 in the CRCs from two monoallelic germline *MUTYH* PV carriers where loss of the wildtype allele was observed in the tumour [19]. In this study, SBS18 and/or SBS36 were not increased in the CRCs from seven of the eight monoallelic *MUTYH* carriers, nor did we find evidence of a second somatic hit in their CRCs. One monoallelic NM_001128425.2:c.504+19_504+31del PV carrier (W07) that exhibited high levels of SBS18/36 also harboured a large somatic deletion across the *MUTYH* gene region, thus providing the second inactivating event. Our findings support the notion that biallelic *MUTYH* inactivation is necessary to promote BER deficiency-related CRC tumourigenesis. This approach could be applied to rare variant classification, including VUSs. We explored this in a CRC (W09) from a carrier of a *MUTYH* PV (p.Gly396Asp) and a VUS (p.Ser304Arg). The absence of both SBS18 and SBS36 suggests that this VUS is not pathogenic and could be further supported by exclusion of the variant being *in cis* with the p.Gly396Asp PV through parent genotyping.

The current proposed mechanism for the accumulation of MMRd associated somatic mutations is based on polymerase slippage, particularly during replication of low complexity regions such as homopolymers, followed by defective repair of these errors [39]. Our results reflect this underlying molecular mechanism, showing that ID signatures more effectively differentiated MMRd CRCs from MMRp CRCs compared with SBS TMS. ID2 and ID7 were the most relevant TMS out of the 10 MMRd-related TMS assessed, and are primarily composed of 1bp homopolymer deletions. The combined ID2 and ID7 TMS profile was able to differentiate MMRd CRC from MMRp CRC with high accuracy (validation set AUC 0.986), a result that is comparable to MMR immunohistochemistry [40,41], which is recommended to be performed on all newly diagnosed CRCs to screen for Lynch syndrome [8]. As tumour sequencing becomes more widely implemented, calculating TMS on CRCs could supersede the need for MMR immunohistochemistry. When comparing MMR PV carriers to the MMRd control group, SBS1 had the greatest capacity to separate the two groups, but with substantial overlap (AUC 0.805; p=0.020). From our analysis we find that the current TMS framework cannot completely differentiate MMR PV carriers from *MLH1* methylated CRCs, but note that age of diagnosis was significantly different between these two groups (p=1.4×10^−8^), and may be a confounder for the association observed with SBS1, as this particular TMS is known to be associated with aging (referred to as the “clock-like signature” in COSMIC). A larger age-matched study could determine if SBS1 is informative for this comparison or rather, a proxy for age. Investigating novel approaches to derive TMS that are focused on differentiating these two important MMRd subtypes may provide a better discrimination tool.

We demonstrated experimental settings where our TMS findings were robust. A minimum VAF threshold from 0.05 to 0.2 and minimum DP threshold from 25 to 100 resulted in stable somatic mutation counts and consequently provided robustness of the hereditary CRC-associated TMS. Filtering VAF and DP too leniently (increased artefacts) or too stringently (reduced number of variants) resulted in substantial changes to the TMS proportions, including loss of the biallelic *MUTYH* and Lynch syndrome-related TMS in the CRC. To overcome a potential limitation of selecting a set of fixed optimal filtering settings we generated progression graphs that illustrate the robustness of a tumour’s TMS profile across different filtering settings (**Figure 5a-b, 6a-f**), providing a visual confirmation of a single tumour’s TMS robustness when compared to presenting a tumour’s profile as a “mutagraph” (**Figure 2**). The progression graphs demonstrate that these potential *MUTYH* and MMR PV diagnostic TMS are not diminished by subtle changes in experimental conditions related to tumour purity, differences in variant calling settings and DNA sequencing technology that might be experienced in different laboratories across the world, adding to the likelihood that TMS analysis can be implemented in routine diagnostic tumour testing. Furthermore, our findings were replicated between FFPE CRCs and fresh-frozen CRCs from TCGA, providing further evidence of the robustness of our findings and the potential implementation of this approach.

This study has some limitations. Doublet signatures were excluded from this study due to low numbers in WES data, and high reconstruction error, although doublet signatures are not reported to be a significant component of CRC [16]. We developed this TMS profiling approach and thresholds using WES data, however, further calibration of these TMS thresholds may be required when applied to whole genome tumour sequencing or targeted tumour sequencing of CRCs, where greater and fewer numbers of somatic mutations, respectively, may influence TMS resolution and reconstruction error. In addition, biallelic *MUTYH* and Lynch syndrome carriers develop extra-colonic cancers, however, extrapolation of our CRC TMS findings to extra-colonic cancers is unknown. Validation of our findings in relation to PV-specific TMS, and elevated SNV-derived mutational burden in CRCs from homozygous p.Tyr179Cys PV, is needed. It remains to be determined if the TMS composition will differ for carriers of *MUTYH* PVs other than these two PVs that are relatively common in individuals with European ancestry. Additional application of the biallelic *MUTYH* SBS18/SBS36 TMS for variant classification needs to be demonstrated.

## CONCLUSIONS

Understanding the somatic mutational landscape can enhance precision oncology by enabling us to pinpoint biomarkers relevant to diagnosis and targeted treatment [42]. As access to tumour sequencing increases, the opportunity to derive TMS, at minimal additional cost, can provide improved diagnostic yield and guide therapeutic options [21]. We have shown that CRCs from biallelic *MUTYH* carriers exhibit TMS profiles that distinguish them from CRCs from non-carriers, evidenced by the combination of SBS18 and SBS36. These distinct TMS profiles have the potential to aid in variant classification for these genes. Furthermore, we identified a novel association between both the proportion of SBS18 and SBS36 and the SNV-derived TMB for specific *MUTYH* PVs. Our results highlight the additional utility of ID-derived TMS for determining defective DNA MMR where the combination of ID2 and ID7 effectively differentiated MMR PV carriers from MMRp CRCs. We have shown that TMS generated from WES of FFPE-CRCs can effectively identify carriers of hereditary CRC and polyposis syndromes and provides a functional assay to aid in the clinical genetics of CRC. Further work is needed to distinguish germline MMR-deficiency from somatic MMR-deficiency in order to improve the diagnosis of Lynch syndrome.

### Funding/Support

Funding by a National Health and Medical Research Council of Australia (NHMRC) project grant GNT1125269 (PI-Daniel Buchanan), supported the design, analysis and interpretation of data. PG is supported by an Australian Government Research Training Program Scholarship. DDB is supported by a NHMRC R.D. Wright Career Development Fellowship (GNT1125268) and funding from the University of Melbourne Research at Melbourne Accelerator Program (R@MAP). AKW is a NHMRC Career Development Fellow. MAJ is a NHMRC Senior Research Fellow. JLH is a NHMRC Senior Principal Research Fellow. O.M.S. is a NHMRC Senior Research Fellow (APP1136119). BJP is supported by a Victorian Health and Medical Research Fellowship.

Research reported in this publication was supported by the National Cancer Institute of the National Institutes of Health under Award Number U01CA167551 and through a cooperative agreement with the Australasian Colorectal Cancer Family Registry (NCI/NIH U01 CA074778 and U01/U24 CA097735) and by the Victorian Cancer Registry, Australia and Ontario Familial Colorectal Cancer Registry (U01/U24 CA074783). This research was performed under CCFR approved projects C-AU-0818-01, C-AU-1014-02, C-AU-0312-01, C-AU-1013-02.

### Financial disclosure

DDB served as a consultant on the Tumour Agnostic (dMMR) Advisory Board of Merck Sharp and Dohme in 2017 and 2018 for Pembrolizumab.

## Data Availability

Data are available upon reasonable request.

## Acknowledgments

The authors thank members of the Colorectal Oncogenomics Group for their support of this manuscript. We thank the participants and staff from the Australasian and Ontario sites of the Colon-CFR in particular, Nicole Mealey, Maggie Angelakos, Samantha Fox and Allyson Templeton for their support of this manuscript. We thank the participants from the ANGELS study and the Family Cancer Clinics across Australia. The authors thank the Victorian Cancer BioBank and Biogrid Australia for provision of patient specimens and clinical data for the WEHI group. This research was undertaken using the LIEF HPC-GPGPU Facility hosted at the University of Melbourne. This Facility was established with the assistance of LIEF Grant LE170100200. The authors thank the Australian Genome Research Facility (AGRF) and Melbourne Bioinformatics for their support of this work.

“The content of this manuscript does not necessarily reflect the views or policies of the National Cancer Institute or any of the collaborating centers in the Colon Cancer Family Registry (Colon-CFR), nor does mention of trade names, commercial products, or organizations imply endorsement by the US Government or the Colon-CFR.”

## Author contributions

DDB, PG, MC and IMW conceived the original study concept and design and designed the analysis. CR, FAM, IMW, AKW, JLH, MAJ, SG, DB, DOS, NM, PG, OMS, DM contributed to the acquisition of study data. The sample curation and laboratory testing was performed by MC, RW, RH, JJ, SP, SJ, JC. PG, BJP, KM implemented the bioinformatics analysis pipeline. PG, DDB, BJP, MAJ designed and performed the statistical analyses. PG and DDB prepared the manuscript. All authors provided critical revisions to the manuscript for important intellectual content and have read and approved of the final manuscript.

## Ethics

Informed consent was obtained from all ACCFR and OCCFR study participants, and the study protocol was approved by the institutional human ethics committees at both study sites. All ANGELS study participants provided informed consent and the study was approved by the University of Melbourne Human Research Ethics Committee (HREC#1750748) and Institutional review boards at each Family Cancer Clinic. All WEHI patients gave written informed consent, and the study was approved by human research ethics committees at both sites (HREC 12/19).

## Abbreviations

AUC: area under the curve
ACCFR: Australasian Colorectal Cancer Family Registry
BER: base excision repair
COAD: colon adenocarcinoma
CRCs: colorectal cancers
DP: depth of coverage
FFPE: formalin-fixed paraffin embedded
GCPS: Genetics of Colonic Polyposis Study
ID: indel
LD: Fisher’s linear discriminant
MSI: microsatellite instability
MMR: mismatch repair
MMRd: MMR-deficient
MMRp: MMR-proficient
MAP: *MUTYH*-associated polyposis
OCCFR: Ontario Colorectal Cancer Family Registry
PV: pathogenic variant
pp: percentage points
ROC: receiver operating curve
SBS: single base substitution
TCGA: The Cancer Genome Atlas
TMB: tumour mutational burden
TMS: tumour mutational signatures
VAF: variant allele fraction
VUS: variants of uncertain clinical significance
WES: whole exome sequencing

