## Supplementary Material for "Evaluating the utility of tumour mutational signatures for identifying hereditary colorectal cancer and polyposis syndrome carriers"

### SUPPLEMENTARY METHODS

#### Study Participants

The study population included men and women diagnosed with incident invasive primary colon or rectal cancer (CRC) who were enrolled in the Australasian and Ontario sites of the Colon Cancer Family Registry (ACCFR and OCCFR), the ANGELS study and the WEHI study. The CRC-affected participants included in the *hereditary CRC group* for this study were those identified to carry germline biallelic *MUTYH* or monoallelic *MUTYH* pathogenic variants or germline *MLH1*, *MSH2* or *MSH6* pathogenic variants confirmed by ClinVar. The CRC-affected participants included in the *non-hereditary CRC group* for this study were those identified not to carry a germline *MUTYH* or MMR gene pathogenic variant from clinic testing and/or interrogation of the non-tumour reference DNA sample used for whole exome sequencing (WES). Eligible participants from each of the studies included those with CRC tumour and non-tumour DNA tested by WES and that passed quality control assessment (**Figure 1** and **Supplementary Table 3**).

Population-based and clinic-based recruitment from The University of Melbourne (UoM), Victoria, Australia (ACCFR) and Population-based recruitment from Cancer Care Ontario (CCO), Toronto, Canada (OCCFR) included CRC-affected participants between 1998 and 2008. Population-based recruitment for ACCFR was restricted to people with CRC diagnosed before age 60 years. The ACCFR and OCCFR recruitment protocols and eligibility criteria have been previously detailed [1,2] (http://coloncfr.org). Details of the germline MMR and *MUTYH* gene testing and for the MMR IHC and MSI as well as *MLH1* gene promoter DNA methylation testing of CRC tumours have been described [3–7]. Informed consent was obtained from all study participants, and the study protocol was approved by the institutional human ethics committees at both study sites.

CRC-affected participants from the ANGELS study (Applying Novel Genomic approaches to Early-onset and suspected Lynch Syndrome colorectal and endometrial cancers: NHMRC-funding GNT1125269) included in this study were people with early-onset CRC (<50yrs) recruited from a Family Cancer Clinic in Australia and who had undergone germline multi-gene panel testing for CRC and polyposis susceptibility genes with no identified pathogenic variants related to hereditary CRC syndromes (apart from two participants who were monoallelic *MUTYH* pathogenic variant carriers)(**Figure 1**). All included early-onset CRCs were MMR-proficient determined from the Pathology laboratory

MMR IHC testing reports. All ANGELS study participants provided informed consent and the study was approved by the University of Melbourne Human Research Ethics Committee (HREC#1750748) and Institutional review boards at each Family Cancer Clinic.

The WEHI study cohort comprised 82 CRC patients who were treated at the Royal Melbourne Hospital (Parkville, VIC, Australia) and Western Hospital Footscray (Footscray, VIC, Australia), between Jan 1, 1993, and Dec 31, 2009. Fresh-frozen tumour and matched normal tissue specimens were obtained at surgery and accessed via hospital tissue banks. All patients gave written informed consent, and the study was approved by human research ethics committees at both sites (HREC 12/19).

**Whole Exome Sequencing**

For the ACCFR and ANGELS study participants, formalin-fixed paraffin embedded (FFPE) tissues from CRCs were macrodissected and DNA extracted using the QIAamp DNA FFPE Tissue kit (Qiagen, Hilden, Germany) using standard protocols. Peripheral blood-derived DNAs were extracted using DNeasy blood and tissue kit (Qiagen) and sequenced as germline references. Capture of the whole exome was performed using Agilent Clinical Research Exome V2 (Agilent, Santa Clara, CA) with sequencing performed on an Illumina NovaSeq 6000 (San Diego, CA) comprising 150bp paired-end reads at the Australian Genome Research Facility. Mean on-target coverage was 280.5 ± 192.0 (mean ± SD) for FFPE tumour DNA samples and 164.4 ± 79.0 for blood-derived DNA samples. For the WEHI CRCs, exome-enrichment was performed using the TruSeq Exome Enrichment Kit (Illumina) and 100 bp paired-end read sequencing performed on an Illumina HiSeq 2000 at the Australian Genome Research Facility.

**Bioinformatics Pipeline and Analysis**

WES data from each of the studies were processed using the same pipeline and analysis. Adapter sequences were trimmed from the raw FASTQ files using trimmomatic 0.38 [8], then aligned to the GRCh37 human reference genome using BWA 0.7.12. Somatic single-nucleotide variants (SNVs) and short insertions and deletions (indels) were called with Strelka 2.9.2 [9] using Illumina's recommended workflow. Tumour mutational signatures (TMS) were calculated using the pre-defined set of 67 SBS, 11 doublet, and 17 ID signatures published on the COSMIC website as version 3 [10]. Doublet variant counts were too low to reliably reconstruct doublet signatures, so were not analysed further in this study. MSI status was assessed using the method described by MSIseq [11].

The impact and suitability of experimental settings was explored by filtering somatic SNVs and indel variants based on depth of coverage (DP) in the tumour (calculated by the GATK tool AnnotateVcfWithBamDepth) and the somatic variant allele fraction (VAF). TMS were calculated for each sample at each filter setting. We quantitatively evaluated the ability of TMS to separate classes of samples at different filtering settings, for each relevant TMS. We calculated three measures of separation, which were then used to select the best TMS or combination of TMS. We calculated: 1) Fisher's linear discriminant (LD) [12], which measures the ratio of "between-group" variability and "within-group" variability to find filtering settings that produce tightly clustered groups that are well-separated (described in detail below); 2) the difference between the means of the two groups; and 3) p-values were calculated using a one-sided t-test.

**P-value calculations**

When determining the discriminatory utility of TMS (**Table 1, Supplementary Table 5**), p-values were calculated using one-sided t-tests. The test is one-sided given that we were only interested in identifying TMS showing presence of a signal (higher than average), rather than absence of a signal.

We reported adjusted p-values with Bonferroni correction applied [13]. We performed 1095 individual tests for significance; consequently, a raw p-value must be below $4.6 \times 10^{-5}$ to be considered significant. **Supplementary Table 5** indicates all TMS observed to have significant unadjusted p-values, with significant adjusted p-values highlighted.

AUC confidence intervals were calculated using the method described by Hanley and McNeil [14], unless the AUC was measured as 1.0, in which case this method does not provide a confidence interval. For these instances, the confidence interval was estimated using the method suggested by Obuchowski and Lieber [15].

**Distribution of samples in a group**

To determine 5$^{th}$ and 95$^{th}$ percentiles shown on the discrimination graphs (**Figures 5c, 5d, 5e, 5f, 6a, 6b, 6c, 6d, 6e, 6f, 7a, 7b, 7c, 7d**), the calculated TMS from each CRC in the group were fitted to a beta distribution. The beta distribution is a continuous probability distribution defined on the interval from zero to one. Given that TMS values are normalized to a percentage, the possible range of TMS values is bounded, suggesting the beta distribution as an appropriate distribution [16,17].

**Determining confidence levels**

The discrimination graphs report 5% and 95% confidence levels, indicating that if we observed a tumour with the specified combined SBS18 and SBS36 or combined ID2 and ID7 TMS percentage, based on our cohort of samples, the tumour would be 5% or 95% likely to belong to the biallelic *MUTYH* or Lynch syndrome hereditary group of CRCs, respectively.

An equivalent question is, given a measured TMS $x$ in a sample of CRCs, how likely is that TMS value to have been generated by each of the two possible source distributions: the distribution of TMS from CRCs *with* the syndrome of interest, or the distribution of TMS from CRCs *without* the syndrome of interest? Here, we label the distribution of values with the syndrome and those without the syndrome $D_1$ and $D_2$ respectively. We assume that both $D_1$ and $D_2$ follow a beta distribution as discussed previously: $D_1 \sim Beta(\alpha_1, \beta_1^2)$ and $D_2 \sim Beta(\alpha_2, \beta_2^2)$.

Given the two overlapping possible distributions from which the point may have originated, we aim to calculate the conditional probability $P(D=D_1|X=x)$. Given that the probability of observing an exact value is zero, we instead calculate the probability of observing $x$ within a range, that is $P(X=x\pm\delta|D=D_1)$, which can be calculated as the difference in the cumulative distribution function (cdf) at $x+\delta$ and $x-\delta$. i.e. $P(X=x\pm\delta|D=D_1)=cdf(x+\delta, D_1) - cdf(x-\delta, D_1)$.

The application of Bayes' theorem enables the calculation of $P(X=x\pm\delta|D=D_1)$ in terms of the cdf of each distribution:

$$P(D_1|X = x \pm \delta) = \frac{P(X = x \pm \delta|D_1)P(D_1)}{P(X = x \pm \delta)}$$

$$= \frac{cdf(X = x \pm \delta, D_1)P(D_1)}{P(X = x \pm \delta|D_1)P(D_1) + P(X = x \pm \delta|D_2)P(X = x \pm \delta|D_2)}$$

$$= \frac{cdf(X = x \pm \delta, D_1)P(D_1)}{cdf(X = x \pm \delta, D_1)P(D_1) + cdf(X = x \pm \delta, D_2)P(D_2)}$$

This approach requires prior probabilities to be assigned indicating the likelihood of a tumour belonging to each of the two distributions, given no prior information. For this analysis we assume an unbiased prior probability of $P(D_1) = P(D_2) = 0.5$.

We then calculated the confidence levels of interest by considering the probability at TMS values of $x$ across each possible TMS value.

**Fisher's Linear Discriminant**

**Table 1** includes measures of separation between groups, including Fisher's Linear Discriminant (LD), which is based on the distance between the means measured in terms of the average of the standard deviations of the two groups. i.e. given groups A and B with means $\mu_A$ and $\mu_B$ and standard deviations $\sigma_A$ and $\sigma_B$, LD is given by:

$$LD = \frac{\mu_A + \mu_B}{\frac{1}{2}(\sigma_A + \sigma_B)}$$

**Selection of the best combination of TMS**

We applied a greedy algorithm based on stepwise forward selection [18] to find the best combination of TMS to identify the hereditary CRC group of interest while reducing the likelihood of overfitting.

Starting with each individual COSMIC V3 TMS (excluding artefact signatures), we select the TMS with the highest AUC that also has a minimum difference in means between the groups of at least 10%. We then iterate by assessing the AUC when each possible TMS is added to the current best combination of TMS, provided this new combination improves the AUC by at least 10%, and the absolute difference in means between the groups also improves by at least 10%. If the AUC has reached the maximum possible value (100%), then instead the margin between the groups must improve by at least 10%. These requirements ensure that only TMS with a strong additive benefit with the hereditary group are selected. Although it is possible that TMS with legitimate but weaker associations are excluded with this method, this stringency reduces the likelihood of spurious TMS arising due to overfitting.

As an additional verification of the selection algorithm, we employed logistic regression with L1 penalty to select TMS, an approach that favours simple and interpretable models [19]. This algorithm selected the same set of TMS that were selected by forward selection; that is, SBS18 and SBS36 were selected to best differentiate biallelic *MUTYH* carrier CRCs, and ID2 and ID7 were selected to best differentiate both Lynch syndrome-related CRCs from MMRp CRCs, and MMRd from MMRp CRCs. This algorithm did not select any TMS when attempting to classify Lynch syndrome from MMRd CRCs in the discovery set, which is concordant with the findings from the forward selection method.

### Comparison with The Cancer Genome Atlas (TCGA)

The non-hereditary ("sporadic") control groups from the ACCFR included in the study were younger than the mean age of diagnosis in the general population [20]. To compare our hereditary CRC results with CRCs that are more reflective of the age at diagnosis of CRC in the general population we included TCGA colon adenocarcinomas (COAD) and rectal adenocarcinoma (READ) tumours [21] and stratified them as MMR-proficient (MMRp) and MMR-deficient (MMRd).

#### *Determining MMR status from the TCGA COAD dataset*

MSI status was determined for 399 COAD tumours and 137 READ tumours using MSIseq, with thresholds determined using published MSI results [22]. Overall, 446 tumours (318 COAD, 128 READ) were classified as MSS or MSI-L with MSIseq values <0.18, which formed the group of TCGA MMRp controls, and 74 tumours (70 COAD, 4 READ) were classified as MSI-H with MSIseq values >1.9.

*Deriving MLH1 hypermethylated tumours from the TCGA*

For the 74 MSI-H tumours, we determined which of these resulted from hypermethylation of the *MLH1* gene promoter using either TCGA Infinium HM450k methylation data, or Infinium HM27k methylation data. We considered a tumour to be *MLH1* hypermethylated if it met *both* of the following conditions:

1. For HM450k data, the mean value of the *MLH1* probes cg23658326, cg11600697, cg21490561 was greater than 0.2. For HM27k data, the value of the *MLH1* probe cg00893636 was greater than 0.2; and
2. At least three of the genes *CACNA1G, RUNX3, SOCS1, NEUROG1* and *IGF2* exhibited mean methylation levels above 0.2, thus indicating high levels of CIMP (CpG Island Methylator Phenotype).

This method identified 52 MMRd tumours that showed *MLH1* hypermethylation and high levels of CIMP and were thus classified as TCGA MMRd controls for this study.

The mean age of diagnosis of the TCGA MMRp and MMRd groups is 65.7±12.4 years and 74.6±10.7 years, respectively (**Supplementary Table 2**).

*Variant Calling*

Somatic SNVs and indels for the TCGA dataset were generated using Mutect2, then filtered with our established thresholds of minimum depth 50 and minimum variant allele fraction 0.1, to match the filtering strategy employed for the other CRCs in the study. We then generated TMS for each tumour as previously described.

*Validation of hereditary CRC mutational signatures using TCGA tumours*

To further validate our hereditary CRC TMS findings, we tested the effect of the later-onset and predominantly fresh-frozen tumours from TCGA controls. We repeated our analysis, replacing the MMRp cohort (n=160) with the TCGA MMRp cohort (n=446), and the MMRd cohort (n=25) with the TCGA MMRd cohort (n=52).

***Statistical Analyses***

All statistical analyses were performed using Python 3.6.1. We utilised the NumPy scientific programming package v1.16.2 [23] for numerical processing, SciPy v1.1.0 [24] for performing the statistical analyses, as well as scikit-learn v0.20.2 [25] for machine learning and classification algorithms.

AUC was calculated using the Python software package scikit-learn [25], specifically, the method *sklearn.metrics.roc_auc_score*, which accepts as arguments the calculated TMS values for the hereditary CRCs, and the calculated TMS values for the control CRCs, to generate an AUC value based on the trapezoid method.

**Supplementary Table 1.** Metadata and clinical observations associated with each CRC included in this study from the ACCFR, OCCFR, ANGELS and WEHI cohorts.

| ID | Study | Sex | Age Dx | Category | Gene | Germline findings for LS and BER | MMR IHC Status | Tumour site | Histological Type | Grade | MSIseq | Somatic POLE Exonuclease Mutations |
|---|---|---|---|---|---|---|---|---|---|---|---|---|
| C01 | ACCFR Clinic | Male | 50-59 | MMRp control | CON | MMR-prof | Proficient | rectal | adenocarcinoma | Poorly differentiated | 0.09 | |
| C02 | ACCFR Clinic | Male | 60-69 | MMRp control | CON | MMR-prof | Proficient | ascending | adenocarcinoma | Moderately differentiated | 0.10 | |
| C03 | ACCFR Pop | Male | 40-49 | MMRp control | CON | MMR-prof | Proficient | rectal | adenocarcinoma | Well differentiated | 0.03 | |
| C04 | ACCFR Pop | Male | 40-49 | MMRp control | CON | MMR-prof | Proficient | sigmoid | signet ring | Poorly differentiated | 0.05 | |
| C05 | ACCFR Pop | Male | 30-39 | MMRp control | CON | MMR-prof | Proficient | ascending | adenocarcinoma | Moderately differentiated | 0.13 | |
| C06 | ACCFR Pop | Female | 50-59 | MMRp control | CON | MMR-prof | Proficient | rectal | adenocarcinoma | Moderately differentiated | 0.09 | |
| C07 | ACCFR Pop | Male | 20-29 | MMRp control | CON | MMR-prof | Proficient | descending | adenocarcinoma | Moderately differentiated | 0.12 | |
| C08 | ACCFR Pop | Female | 20-29 | MMRp control | CON | MMR-prof | Proficient | sigmoid | adenocarcinoma | Poorly differentiated | 0.06 | |
| C09 | ACCFR Pop | Male | 20-29 | MMRp control | CON | MMR-prof | Proficient | transverse colon | adenocarcinoma | Poorly differentiated | 0.02 | |
| C10 | ACCFR Pop | Male | 40-49 | MMRp control | CON | MMR-prof | Proficient | ascending | adenocarcinoma | Moderately differentiated | 0.10 | |
| C11 | ACCFR Pop | Female | 40-49 | MMRp control | CON | MMR-prof | Proficient | caecum | adenocarcinoma | Moderately differentiated | 0.07 | |
| C12 | ACCFR Pop | Female | 40-49 | MMRp control | CON | MMR-prof | Proficient | transverse colon | adenocarcinoma | Moderately differentiated | 0.07 | |
| C13 | ACCFR Pop | Male | 50-59 | MMRp control | CON | MMR-prof | Proficient | transverse colon | adenocarcinoma | Moderately differentiated | 0.05 | |
| C14 | ACCFR Pop | Male | 50-59 | MMRp control | CON | MMR-prof | Proficient | transverse colon | adenocarcinoma | Moderately differentiated | 0.05 | |
| C15 | ACCFR Pop | Male | 40-49 | MMRp control | CON | MMR-prof | Proficient | sigmoid | adenocarcinoma | Moderately differentiated | 0.07 | |
| C16 | ACCFR Pop | Female | 40-49 | MMRp control | CON | MMR-prof | Proficient | rectal | undifferentiated | Poorly differentiated | 0.05 | |
| C17 | ACCFR Pop | Female | 30-39 | MMRp control | CON | MMR-prof | Proficient | transverse colon | adenocarcinoma | Poorly differentiated | 0.19 | |

| | | | | | | | | | | | | |
|---|---|---|---|---|---|---|---|---|---|---|---|---|
| C18 | ACCFR Clinic | Female | 40-49 | MMRp control | CON | MMR-prof | Proficient | transverse colon | adenocarcinoma | Moderately differentiated | 0.06 | |
| C19 | ACCFR Pop | Female | 20-29 | MMRp control | CON | MMR-prof | Proficient | Sigmoid | adenocarcinoma | Moderately differentiated | 0.22 | |
| C20 | ACCFR Pop | Male | 30-39 | MMRp control | CON | MMR-prof | Proficient | Rectum | adenocarcinoma | Moderately differentiated | 1.61 | c.1231G>T p.Val411Leu |
| C21 | ACCFR Pop | Female | 40-49 | MMRp control | CON | MMR-prof | Proficient | Sigmoid Colon | adenocarcinoma | Moderately differentiated | 0.00 | |
| C22 | ACCFR Pop | Female | 30-39 | MMRp control | CON | MMR-prof | Proficient | Rectum | adenocarcinoma | Poorly differentiated | 0.09 | |
| C23 | ACCFR Pop | Female | 30-39 | MMRp control | CON | MMR-prof | Proficient | Caecum | adenocarcinoma | Moderately differentiated | 0.02 | |
| C24 | ACCFR Pop | Female | 30-39 | MMRp control | CON | MMR-prof | Proficient | Rectum | adenocarcinoma | Moderately differentiated | 0.02 | |
| C25 | ACCFR Pop | Female | 10-19 | MMRp control | CON | MMR-prof | Proficient | Transverse | adenocarcinoma | Poorly differentiated | 0.03 | |
| C26 | ACCFR Pop | Male | 30-39 | MMRp control | CON | MMR-prof | Proficient | Sigmoid | adenocarcinoma | Moderately differentiated | 0.05 | |
| C27 | ACCFR Pop | Male | 30-39 | MMRp control | CON | MMR-prof | Proficient | Caecum | adenocarcinoma | Moderately differentiated | 0.09 | |
| C29 | ACCFR Pop | Female | 30-39 | MMRp control | CON | MMR-prof | Proficient | Rectum | adenocarcinoma | Well to moderately-differentiated | 0.70 | c.857C>G p.Pro286Arg |
| C30 | ACCFR Pop | Female | 30-39 | MMRp control | CON | MMR-prof | Proficient | Ascending | adenocarcinoma | Moderately differentiated | 0.12 | |
| C31 | ANGELs | Male | 40-49 | MMRp control | CON | MMR-prof | Proficient | Rectum | adenocarcinoma | Moderately differentiated | 0.00 | |
| C32 | ANGELs | Female | 30-39 | MMRp control | CON | MMR-prof | Proficient | Rectum | adenocarcinoma | Moderately differentiated | 1.65 | c.1376C>T p.Ser459Phe |
| C33 | ANGELs | Male | 20-29 | MMRp control | CON | MMR-prof | Proficient | Sigmoid | adenocarcinoma | Poorly differentiated | 0.07 | |
| C34 | ANGELs | Male | 40-49 | MMRp control | CON | MMR-prof | Proficient | Rectum | adenocarcinoma | Moderately differentiated | 0.07 | |
| W01 | ANGELs | Female | 40-49 | MUTYH monoallelic | MUTYH | p.Gly396Asp | Proficient | Rectum | adenocarcinoma | Moderately differentiated | 0.06 | |
| C35 | ANGELs | Female | 30-39 | MMRp control | CON | MMR-prof | Proficient | Rectum | adenocarcinoma | Moderately differentiated | 0.07 | |
| C36 | ANGELs | Female | 20-29 | MMRp control | CON | MMR-prof | Proficient | Ascending | adenocarcinoma | Moderately differentiated | 0.05 | |
| C38 | ANGELs | Male | 30-39 | MMRp control | CON | MMR-prof | Proficient | Rectum | adenocarcinoma | Moderately differentiated | 0.03 | |
| C39 | ANGELs | Female | 30-39 | MMRp control | CON | MMR-prof | Proficient | Sigmoid | adenocarcinoma | Moderately differentiated | 0.18 | |
| C40 | ANGELs | Female | 30-39 | MMRp control | CON | MMR-prof | Proficient | Caecum | adenocarcinoma | Poorly differentiated | 0.02 | |

| | | | | | | | | | | | |
|---|---|---|---|---|---|---|---|---|---|---|---|
| C41 | ANGELs | Female | 30-39 | MMRp control | CON | MMR-prof | Proficient | Descending | adenocarcinoma | Moderately differentiated | 0.05 |
| C42 | ANGELs | Male | 30-39 | MMRp control | CON | MMR-prof | Proficient | Transverse | adenocarcinoma | Poorly differentiated | 0.02 |
| C43 | ANGELs | Female | 30-39 | MMRp control | CON | MMR-prof | Proficient | Transverse | adenocarcinoma | Moderately differentiated | 0.03 |
| C44 | ANGELs | Female | 40-49 | MMRp control | CON | MMR-prof | Proficient | Sigmoid | adenocarcinoma | Moderately differentiated | 0.10 |
| C45 | ANGELs | Female | 20-29 | MMRp control | CON | MMR-prof | Proficient | Sigmoid | adenocarcinoma | Moderately differentiated | 0.05 |
| C46 | ANGELs | Female | 30-39 | MMRp control | CON | MMR-prof | Proficient | Sigmoid | adenocarcinoma | Moderately differentiated | 0.06 |
| C47 | ANGELs | Female | 30-39 | MMRp control | CON | MMR-prof | Proficient | Sigmoid | adenocarcinoma | Moderately differentiated | 0.02 |
| C48 | ANGELs | Male | 30-39 | MMRp control | CON | MMR-prof | Proficient | Transverse | adenocarcinoma | Moderately differentiated | 0.02 |
| C49 | ANGELs | Female | 30-39 | MMRp control | CON | MMR-prof | Proficient | Caecum | adenocarcinoma | Poorly differentiated | 0.02 |
| C50 | ANGELs | Female | 30-39 | MMRp control | CON | MMR-prof | Proficient | Rectum | adenocarcinoma | Moderately differentiated | 0.06 |
| C51 | ANGELs | Female | 30-39 | MMRp control | CON | MMR-prof | Proficient | Rectum | adenocarcinoma | Moderately differentiated | 0.12 |
| W02 | ANGELs | Female | 40-49 | MUTYH monoallelic | MUTYH | p.Arg245His | Proficient | Caecum | adenocarcinoma | Moderately differentiated | 0.07 |
| C52 | ANGELs | Female | 30-39 | MMRp control | CON | MMR-prof | Proficient | Sigmoid | adenocarcinoma | Poorly differentiated | 0.07 |
| C53 | ANGELs | Female | 30-39 | MMRp control | CON | MMR-prof | Proficient | Sigmoid | adenocarcinoma | Moderately differentiated | 0.03 |
| C54 | ANGELs | Female | 30-39 | MMRp control | CON | MMR-prof | Proficient | Ascending | adenocarcinoma | Moderately differentiated | 0.12 |
| C55 | ANGELs | Female | 40-49 | MMRp control | CON | MMR-prof | Proficient | Sigmoid | adenocarcinoma | Moderately differentiated | 0.07 |
| C56 | ANGELs | Female | 20-29 | MMRp control | CON | MMR-prof | Proficient | Rectum | adenocarcinoma | Moderately differentiated | 0.03 |
| C57 | ANGELs | Female | 40-49 | MMRp control | CON | MMR-prof | Proficient | Ascending | adenocarcinoma | Moderately differentiated | 0.12 |
| C58 | ANGELs | Female | 40-49 | MMRp control | CON | MMR-prof | Proficient | Rectum | adenocarcinoma | Moderately differentiated | 0.10 |
| C59 | ANGELs | Female | 30-39 | MMRp control | CON | MMR-prof | Proficient | Ascending | adenocarcinoma | Moderately differentiated | 0.70 |
| C60 | ANGELs | Male | 40-49 | MMRp control | CON | MMR-prof | Proficient | Rectum | adenocarcinoma | Moderately differentiated | 0.05 |
| C61 | ANGELs | Female | 40-49 | MMRp control | CON | MMR-prof | Proficient | Sigmoid | adenocarcinoma | Moderately differentiated | 0.03 |

| | | | | | | | | | | | |
|---|---|---|---|---|---|---|---|---|---|---|---|
| C62 | ANGELs | Female | 30-39 | MMRp control | CON | MMR-prof | Proficient | Sigmoid | adenocarcinoma | Moderately differentiated | 0.24 |
| C63 | ANGELs | Female | 30-39 | MMRp control | CON | MMR-prof | Proficient | Sigmoid | adenocarcinoma | Moderately differentiated | 0.06 |
| C64 | ANGELs | Male | 20-29 | MMRp control | CON | MMR-prof | Proficient | Sigmoid | adenocarcinoma | Poorly differentiated | 0.09 |
| C65 | ANGELs | Male | 20-29 | MMRp control | CON | MMR-prof | Proficient | Caecum | adenocarcinoma | Poorly differentiated | 0.19 |
| C66 | ANGELs | Female | 40-49 | MMRp control | CON | MMR-prof | Proficient | Sigmoid | adenocarcinoma | Moderately differentiated | 0.05 |
| C67 | ANGELs | Female | 40-49 | MMRp control | CON | MMR-prof | Proficient | Ascending | adenocarcinoma | Moderately differentiated | 0.16 |
| C68 | ANGELs | Female | 30-39 | MMRp control | CON | MMR-prof | Proficient | Sigmoid | mucinous | Moderately differentiated | 0.09 |
| C69 | ANGELs | Female | 30-39 | MMRp control | CON | MMR-prof | Proficient | Rectum | adenocarcinoma | Moderately differentiated | 0.03 |
| C70 | ANGELs | Female | 30-39 | MMRp control | CON | MMR-prof | Proficient | Transverse | adenocarcinoma | Poorly differentiated | 0.07 |
| C71 | ACCFR Clinic | Female | 60-69 | MMRp control | NTHL1 | p.Gln287Ter | Proficient | rectal | mucinous | Poorly differentiated | 0.02 |
| C72 | ACCFR Pop | Male | 40-49 | MMRp control | NTHL1 | p.Gln90Ter | Proficient | rectal | adenocarcinoma | Moderately differentiated | 0.07 |
| K01 | ACCFR Pop | Female | 70-79 | MMRd control | MLH1 | MLH1 methylation | Deficient | N/A | adenocarcinoma | Moderately differentiated | 18.63 |
| K02 | ACCFR Pop | Female | 40-49 | MMRd control | MLH1 | MLH1 methylation | Deficient | transverse colon | adenocarcinoma | Poorly differentiated | 14.25 |
| K03 | ACCFR Pop | Female | 60-69 | MMRd control | MLH1 | MLH1 methylation | Deficient | N/A | mucinous | Moderately differentiated | 4.87 |
| K04 | ACCFR Pop | Male | 50-59 | MMRd control | MLH1 | MLH1 methylation | Deficient | caecum | adenocarcinoma | Moderately differentiated | 13.55 |
| K05 | ACCFR Pop | Female | 50-59 | MMRd control | MLH1 | MLH1 methylation | Deficient | transverse colon | mucinous | Moderately differentiated | 20.91 |
| K06 | ACCFR Pop | Female | 50-59 | MMRd control | MLH1 | MLH1 methylation | Deficient | caecum | adenocarcinoma | Moderately differentiated | 13.49 |
| K07 | ACCFR Pop | Female | 40-49 | MMRd control | MLH1 | MLH1 methylation | Deficient | transverse colon | mucinous | Poorly differentiated | 13.79 |
| K08 | ACCFR Pop | Female | 50-59 | MMRd control | MLH1 | MLH1 methylation | Deficient | caecum | mucinous | Poorly differentiated | 30.69 |
| K09 | ACCFR Pop | Female | 50-59 | MMRd control | MLH1 | MLH1 methylation | Deficient | caecum | adenocarcinoma | Moderately differentiated | 23.33 |
| K10 | ACCFR Pop | Female | 40-49 | MMRd control | MLH1 | MLH1 methylation | Deficient | Caecum | adenocarcinoma | Moderately differentiated | 9.11 |
| L01 | ACCFR Pop | Male | 30-39 | Lynch | MSH2 | del x8 | Deficient | transverse colon | mucinous | Poorly differentiated | 7.10 |

| | | | | | | | | | | | |
|---|---|---|---|---|---|---|---|---|---|---|---|
| L02 | ACCFR Pop | Male | 50-59 | Lynch | MSH2 | p.Gly692CysfsX4 | Deficient | rectal | mucinous | Moderately differentiated | 18.80 |
| L20 | ACCFR Pop | Male | 30-39 | Lynch | MSH6 | p.Gln475Ter | Deficient | caecum | adenocarcinoma | Moderately differentiated | 4.55 |
| L21 | ACCFR Pop | Male | 20-29 | Lynch | MSH6 | p.Phe1104TrpfsX3 | Deficient | caecum | adenocarcinoma | Poorly differentiated | 7.59 |
| L22 | ACCFR Pop | Male | 30-39 | Lynch | MSH6 | p.Val653Ter | Deficient | sigmoid | mucinous | Moderately differentiated | 14.65 |
| L23 | ACCFR Pop | Male | 40-49 | Lynch | MSH6 | p.Ala1320GlufsX6 | Deficient | ascending | mucinous | Moderately differentiated | 3.00 |
| L30 | ACCFR Clinic | Male | 30-39 | Lynch | MLH1 | p.Ser225ValfsX4 | Deficient | caecum | mucinous | Moderately differentiated | 1.47 |
| L31 | ACCFR Clinic | Male | 40-49 | Lynch | MLH1 | p.Gln26Ter | Deficient | transverse colon | adenocarcinoma | Moderately differentiated | 1.38 |
| L32 | ACCFR Pop | Male | 20-29 | Lynch | MLH1 | p.Lys618del | Deficient | sigmoid | adenocarcinoma | Moderately differentiated | 17.12 |
| L33 | ACCFR Pop | Male | 20-29 | Lynch | MLH1 | p.(Ser225ValfsTer4) | Deficient | caecum | adenocarcinoma | Poorly differentiated | 13.64 |
| L34 | ACCFR Pop | Male | 40-49 | Lynch | MLH1 | p.Arg487Ter | Deficient | transverse colon | mucinous | Moderately differentiated | 18.38 |
| L35 | ACCFR Pop | Female | 40-49 | Lynch | MLH1 | p.Tyr548Ter | Deficient | caecum | adenocarcinoma | Poorly differentiated | 45.71 |
| L36 | ACCFR Pop | Female | 40-49 | Lynch | MLH1 | p.Thr117Met | Deficient | ascending | mucinous | Moderately differentiated | 30.46 |
| L03 | ACCFR Pop | Female | 50-59 | Lynch | MSH2 | c.2005+3_2005+14del12 | Deficient | ascending | adenocarcinoma | Moderately differentiated | 23.85 |
| L04 | ACCFR Pop | Female | 40-49 | Lynch | MSH2 | p.Pro622Leu | Deficient | Rectum | adenocarcinoma | Moderately differentiated | 20.43 |
| L24 | ACCFR Pop | Male | 50-59 | Lynch | MSH6 | p.Tyr524X | Deficient | Sigmoid Colon | mucinous | Moderately differentiated | 5.63 |
| L05 | ACCFR Pop | Male | 40-49 | Lynch | MSH2 | p.Gly630GlufsX4 | Deficient | Descending Colon | adenocarcinoma | Moderately differentiated | 14.28 |
| L37 | ACCFR Clinic | Female | 30-39 | Lynch | MLH1 | | Deficient | transverse colon | adenocarcinoma | Moderately differentiated | 35.37 |
| L38 | ACCFR Clinic | Female | 30-39 | Lynch | MLH1 | p.Thr117Met | Deficient | rectosigmoid colon | adenocarcinoma | Moderately differentiated | 23.39 |
| L39 | ACCFR Clinic | Female | 30-39 | Lynch | MLH1 | c.350C>T p.Thr117Met | Deficient | transverse colon | adenocarcinoma | Moderately differentiated | 10.15 |
| L40 | ACCFR Clinic | Male | 40-49 | Lynch | MLH1 | c.672delT p.Ser225ValfsX4 | Deficient | caecum | adenocarcinoma | Well differentiated | 13.88 |
| L41 | ACCFR Clinic | Male | 40-49 | Lynch | MLH1 | c.1975C>T p.Arg659X | Deficient | sigmoid | adenocarcinoma | Poorly differentiated | 21.64 |
| L06 | ACCFR Clinic | Female | 40-49 | Lynch | MSH2 | c.2038C>T p.Arg680X | Deficient | ascending colon | adenocarcinoma | Moderately differentiated | 12.97 |

| | | | | | | | | | | | |
|---|---|---|---|---|---|---|---|---|---|---|---|
| L07 | ACCFR Clinic | Male | 40-49 | Lynch | MSH2 | c.1165C>T p.Arg389X | Deficient | ascending colon | signet ring cell carcinoma | Poorly differentiated | 9.53 |
| L08 | ACCFR Clinic | Female | 40-49 | Lynch | MSH2 | IVS5+3A>T | Deficient | large bowel | adenocarcinoma | Poorly differentiated | 18.55 |
| L42 | ACCFR Clinic | Female | 50-59 | Lynch | MLH1 | c.420delA p.Lys140fs | Deficient | transverse colon | adenocarcinoma | Moderately differentiated | 7.77 |
| L09 | ACCFR Clinic | Male | 50-59 | Lynch | MSH2 | c.2285T>A p.Leu762X | Deficient | ileocolic anastomosis | adenocarcinoma | Moderately differentiated | 16.75 |
| L43 | ACCFR Clinic | Female | 60-69 | Lynch | MLH1 | c.1667G>A p.Ser556Asn | Deficient | Right Colon | adenocarcinoma | Poorly differentiated | 24.61 |
| L10 | ACCFR Clinic | Female | 60-69 | Lynch | MSH2 | | Deficient | hepatic flexure | adenocarcinoma | Moderately differentiated | 1.40 |
| L11 | ACCFR Clinic | Female | 50-59 | Lynch | MSH2 | c.2285T>A p.Leu762X | Deficient | ileocolic anastomosis | adenocarcinoma | Moderately differentiated | 25.32 |
| M01 | ACCFR Clinic | Male | 60-69 | MUTYH biallelic | MUTYH | p.Tyr179Cys | Proficient | ascending | adenocarcinoma | Moderately differentiated | 0.06 |
| M02 | ACCFR Clinic | Male | 60-69 | MUTYH biallelic | MUTYH | p.Tyr179Cys | Proficient | ascending | adenocarcinoma | Moderately differentiated | 0.06 |
| M03 | ACCFR Clinic | Male | 60-69 | MUTYH biallelic | MUTYH | p.Tyr179Cys | Proficient | caecum | adenocarcinoma | Moderately differentiated | 0.12 |
| M04 | ACCFR Clinic | Female | 50-59 | MUTYH biallelic | MUTYH | p.Tyr179Cys | Proficient | ascending | adenocarcinoma | Moderately differentiated | 0.07 |
| M05 | ACCFR Clinic | Female | 50-59 | MUTYH biallelic | MUTYH | p.Tyr179Cys | Proficient | ascending | adenocarcinoma | Moderately differentiated | 0.09 |
| M06 | ACCFR Clinic | Male | 60-69 | MUTYH biallelic | MUTYH | p.Gly396Asp | Proficient | caecum | adenocarcinoma | Unknown | 0.06 |
| M07 | ACCFR Clinic | Male | 60-69 | MUTYH biallelic | MUTYH | p.Gly396Asp | Proficient | unknown | adenocarcinoma | Moderately differentiated | 0.03 |
| M08 | ACCFR Pop | Male | 50-59 | MUTYH biallelic | MUTYH | p.Gly396Asp | Proficient | rectal | adenocarcinoma | Moderately differentiated | 0.07 |
| M09 | ACCFR Clinic | Male | 50-59 | MUTYH biallelic | MUTYH | p.Tyr179Cys; p.Arg182His | Proficient | sigmoid | adenocarcinoma | Moderately differentiated | 0.07 |
| M10 | ACCFR Pop | Male | 30-39 | MUTYH biallelic | MUTYH | p.Tyr179Cys; p.Arg245His | Proficient | ascending | adenocarcinoma | Moderately differentiated | 0.10 |
| W03 | ACCFR Clinic | Female | 60-69 | MUTYH monoallelic | MUTYH | p.Gly396Asp | Proficient | sigmoid | mucinous | Moderately differentiated | 0.10 |
| W09 | ACCFR Pop | Male | 40-49 | MUTYH VUS | MUTYH | p.Gly396Asp; p.Ser304Arg | Proficient | sigmoid | adenocarcinoma | Moderately differentiated | 0.09 |
| W04 | ACCFR Pop | Female | 30-39 | MUTYH monoallelic | MUTYH | p.Gly396Asp | Proficient | sigmoid | adenocarcinoma | Moderately differentiated | 0.06 |
| W05 | ACCFR Pop | Male | 40-49 | MUTYH monoallelic | MUTYH | p.Gly396Asp | Proficient | sigmoid | adenocarcinoma | Moderately differentiated | 0.05 |

| | | | | | | | | | | | |
|---|---|---|---|---|---|---|---|---|---|---|---|
| C73 | WEHI | Male | 70-79 | MMRp control | CON | MMR-prof | Proficient | Rectosigmoid | Adenocarcioma | Well/Moderate | 0.09 |
| W06 | WEHI | Male | 70-79 | MUTYH monoallelic | CON | p.Tyr179Cys | Proficient | Rectum | Adenocarcioma | Well/Moderate | 0.05 |
| C74 | WEHI | Female | 60-69 | MMRp control | CON | MMR-prof | Proficient | Ascending | Adenocarcioma | Well/Moderate | 0.05 |
| C75 | WEHI | Male | 70-79 | MMRp control | CON | MMR-prof | Proficient | Hepatic flexure | Adenocarcioma | Well/Moderate | 0.07 |
| C76 | WEHI | Female | 40-49 | MMRp control | CON | MMR-prof | Proficient | Transverse | Adenocarcioma | Well/Moderate | 0.15 |
| C77 | WEHI | Male | 60-69 | MMRp control | CON | MMR-prof | Proficient | Sigmoid | Adenocarcioma | Poor | 0.02 |
| C78 | WEHI | Male | 30-39 | MMRp control | CON | MMR-prof | Proficient | Sigmoid | Adenocarcioma | Well/Moderate | 0.05 |
| C79 | WEHI | Male | 70-79 | MMRp control | CON | MMR-prof | Proficient | Sigmoid | Adenocarcioma | Well/Moderate | 0.10 |
| C80 | WEHI | Female | 60-69 | MMRp control | CON | MMR-prof | Proficient | Sigmoid | Adenocarcioma | Well/Moderate | 0.07 |
| C81 | WEHI | Male | 70-79 | MMRp control | CON | MMR-prof | Proficient | Caecum | Adenocarcioma | Well/Moderate | 0.10 |
| C82 | WEHI | Male | 70-79 | MMRp control | CON | MMR-prof | Proficient | Caecum | Adenocarcioma | Poor | 0.07 |
| C83 | WEHI | Male | 80-89 | MMRp control | CON | MMR-prof | Proficient | Sigmoid | Adenocarcioma | Well/Moderate | 0.06 |
| C84 | WEHI | Male | 70-79 | MMRp control | CON | MMR-prof | Proficient | Ascending | Adenocarcioma | Well/Moderate | 0.21 |
| C85 | WEHI | Female | 70-79 | MMRp control | CON | MMR-prof | Proficient | Sigmoid | Adenocarcioma | Well/Moderate | 0.03 |
| C86 | WEHI | Male | 70-79 | MMRp control | CON | MMR-prof | Proficient | Sigmoid | Adenocarcioma | NA | 0.03 |
| C87 | WEHI | Male | 80-89 | MMRp control | CON | MMR-prof | Proficient | Sigmoid | Adenocarcioma | Well/Moderate | 0.09 |
| C88 | WEHI | Male | 70-79 | MMRp control | CON | MMR-prof | Proficient | Rectosigmoid | Adenocarcioma | Well/Moderate | 0.06 |
| C89 | WEHI | Male | 70-79 | MMRp control | CON | MMR-prof | Proficient | Caecum | Adenocarcioma | Well/Moderate | 0.09 |
| C90 | WEHI | Male | 60-69 | MMRp control | CON | MMR-prof | Proficient | Splenic flexure | Adenocarcioma | Poor | 0.03 |
| C91 | WEHI | Male | 50-59 | MMRp control | CON | MMR-prof | Proficient | Sigmoid | Adenocarcioma | Well/Moderate | 0.12 |
| C92 | WEHI | Female | 70-79 | MMRp control | CON | MMR-prof | Proficient | Caecum | Adenocarcioma | Well/Moderate | 0.07 |
| C93 | WEHI | Female | 70-79 | MMRp control | CON | MMR-prof | Proficient | Caecum | Adenocarcioma | Poor | 0.27 |

| | | | | | | | | | | | |
|---|---|---|---|---|---|---|---|---|---|---|---|
| C94 | WEHI | Male | 50-59 | MMRp control | CON | MMR-prof | Proficient | Rectum | Adenocarcioma | Well/Moderate | 0.05 |
| C95 | WEHI | Male | 40-49 | MMRp control | CON | MMR-prof | Proficient | Sigmoid | Adenocarcioma | Poor | 0.13 |
| C96 | WEHI | Male | 80-89 | MMRp control | CON | MMR-prof | Proficient | Rectosigmoid | Adenocarcioma | Well/Moderate | 0.18 |
| C97 | WEHI | Female | 60-69 | MMRp control | CON | MMR-prof | Proficient | Sigmoid | Adenocarcioma | Poor | 0.09 |
| C98 | WEHI | Female | 60-69 | MMRp control | CON | MMR-prof | Proficient | Ascending | Adenocarcioma | Poor | 0.05 |
| C99 | WEHI | Male | 60-69 | MMRp control | CON | MMR-prof | Proficient | Sigmoid | Adenocarcioma | Well/Moderate | 0.02 |
| C100 | WEHI | Female | 60-69 | MMRp control | CON | MMR-prof | Proficient | Caecum | Adenocarcioma | Well/Moderate | 0.10 |
| C101 | WEHI | Female | 60-69 | MMRp control | CON | MMR-prof | Proficient | Caecum | Adenocarcioma | Well/Moderate | 0.15 |
| W07 | WEHI | Male | 50-59 | MUTYH monoallelic | MUTYH | c.504+19_504+31 del | Proficient | Rectosigmoid | Adenocarcioma | Well/Moderate | 0.07 |
| C102 | WEHI | Male | 70-79 | MMRp control | CON | MMR-prof | Proficient | Ascending | Adenocarcioma | Well/Moderate | 0.09 |
| C103 | WEHI | Male | 60-69 | MMRp control | CON | MMR-prof | Proficient | Rectum | Adenocarcioma | Well/Moderate | 0.05 |
| W08 | WEHI | Female | 80-89 | MUTYH monoallelic | MUTYH | p.Gly396Asp | Proficient | Rectum | Adenocarcioma | Well/Moderate | 0.05 |
| C104 | WEHI | Male | 60-69 | MMRp control | CON | MMR-prof | Proficient | Sigmoid | Adenocarcioma | Poor | 0.10 |
| C105 | WEHI | Male | 80-89 | MMRp control | CON | MMR-prof | Proficient | Splenic flexure | Adenocarcioma | Well/Moderate | 0.12 |
| C106 | WEHI | Female | 30-39 | MMRp control | CON | MMR-prof | Proficient | Sigmoid | Adenocarcioma | Well/Moderate | 0.02 |
| C107 | WEHI | Male | 60-69 | MMRp control | CON | MMR-prof | Proficient | Rectum | Adenocarcioma | Well/Moderate | 0.15 |
| C108 | WEHI | Female | 60-69 | MMRp control | CON | MMR-prof | Proficient | Caecum | Adenocarcioma | Well/Moderate | 0.18 |
| C109 | WEHI | Male | 70-79 | MMRp control | CON | MMR-prof | Proficient | Splenic flexure | Adenocarcioma | Well/Moderate | 0.02 |
| C110 | WEHI | Female | 50-59 | MMRp control | CON | MMR-prof | Proficient | Sigmoid | Adenocarcioma | Well/Moderate | 0.06 |
| C111 | WEHI | Male | 70-79 | MMRp control | CON | MMR-prof | Proficient | Sigmoid | Adenocarcioma | Well/Moderate | 0.03 |
| C112 | WEHI | Male | 60-69 | MMRp control | CON | MMR-prof | Proficient | Rectosigmoid | Adenocarcioma | Poor | 0.03 |

| | | | | | | | | | | | |
|---|---|---|---|---|---|---|---|---|---|---|---|
| C113 | WEHI | Female | 70-79 | MMRp control | CON | MMR-prof | Proficient | Caecum | Adenocarcioma | NA | 0.06 |
| C114 | WEHI | Female | 50-59 | MMRp control | CON | MMR-prof | Proficient | Rectosigmoid | Adenocarcioma | Well/Moderate | 0.16 |
| C115 | WEHI | Female | 70-79 | MMRp control | CON | MMR-prof | Proficient | Rectum | Adenocarcioma | Well/Moderate | 0.18 |
| C116 | WEHI | Female | 40-49 | MMRp control | CON | MMR-prof | Proficient | Rectum | Adenocarcioma | Well/Moderate | 0.05 |
| C117 | WEHI | Female | 60-69 | MMRp control | CON | MMR-prof | Proficient | Rectosigmoid | Adenocarcioma | Well/Moderate | 0.05 |
| C118 | WEHI | Male | 70-79 | MMRp control | CON | MMR-prof | Proficient | Rectum | Adenocarcioma | Poor | 0.16 |
| C119 | WEHI | Female | 60-69 | MMRp control | CON | MMR-prof | Proficient | Caecum | Adenocarcioma | Well/Moderate | 0.22 |
| C120 | WEHI | Female | 70-79 | MMRp control | CON | MMR-prof | Proficient | Ascending | Adenocarcioma | Well/Moderate | 0.07 |
| C121 | WEHI | Female | 70-79 | MMRp control | CON | MMR-prof | Proficient | Rectosigmoid | Adenocarcioma | Well/Moderate | 0.07 |
| C122 | WEHI | Male | 70-79 | MMRp control | CON | MMR-prof | Proficient | Sigmoid | Adenocarcioma | Poor | 0.05 |
| C123 | WEHI | Female | 60-69 | MMRp control | CON | MMR-prof | Proficient | Rectosigmoid | Adenocarcioma | Well/Moderate | 0.02 |
| C124 | WEHI | Female | 50-59 | MMRp control | CON | MMR-prof | Proficient | Sigmoid | Adenocarcioma | Well/Moderate | 0.07 |
| C125 | WEHI | Male | 70-79 | MMRp control | CON | MMR-prof | Proficient | Caecum | Adenocarcioma | Well/Moderate | 0.07 |
| C126 | WEHI | Male | 80-89 | MMRp control | CON | MMR-prof | Proficient | Splenic flexure | Adenocarcioma | Well/Moderate | 0.09 |
| C127 | WEHI | Female | 60-69 | MMRp control | CON | MMR-prof | Proficient | Sigmoid | Adenocarcioma | Poor | 0.06 |
| M11 | OCCFR | Female | 30-39 | MUTYH biallelic | MUTY H | c.536A>G c.312C>A | Proficient | Caecum | Unknown | Moderately differentiated | 0.02 |
| C128 | OCCFR | Female | 40-49 | MMRp control | CON | MMR-prof | Proficient | Rectum | Adenocarcinoma | Unknown | 0.06 |
| C129 | OCCFR | Female | 40-49 | MMRp control | CON | MMR-prof | Proficient | Rectum | Other | Poorly differentiated | 0.05 |
| C130 | OCCFR | Male | 40-49 | MMRp control | CON | MMR-prof | Proficient | Sigmoid | Adenocarcinoma | Moderately differentiated | 0.02 |
| C131 | OCCFR | Female | 40-49 | MMRp control | CON | MMR-prof | Proficient | Sigmoid | Other | Moderately differentiated | 0.10 |
| C132 | OCCFR | Female | 30-39 | MMRp control | CON | MMR-prof | Proficient | Sigmoid | Adenocarcinoma | Moderately differentiated | 0.07 |
| C133 | OCCFR | Female | 40-49 | MMRp control | CON | MMR-prof | Proficient | Descending | Adenocarcinoma | Moderately differentiated | 0.10 |

| | | | | | | | | | | | |
|---|---|---|---|---|---|---|---|---|---|---|---|
| C134 | OCCFR | Female | 40-49 | MMRp control | CON | MMR-prof | Proficient | Sigmoid | Adenocarcinoma | Moderately differentiated | 0.07 |
| C135 | OCCFR | Female | 40-49 | MMRp control | CON | MMR-prof | Proficient | Sigmoid | Adenocarcinoma | Moderately differentiated | 0.06 |
| C136 | OCCFR | Male | 40-49 | MMRp control | CON | MMR-prof | Proficient | Transverse | Adenocarcinoma | Moderately differentiated | 0.09 |
| C137 | OCCFR | Male | 40-49 | MMRp control | CON | MMR-prof | Proficient | Caecum | Adenocarcinoma | Well differentiated | 0.09 |
| C138 | OCCFR | Female | 40-49 | MMRp control | CON | MMR-prof | Proficient | Ascending | Adenocarcinoma | Moderately differentiated | 0.10 |
| C139 | OCCFR | Female | 40-49 | MMRp control | CON | MMR-prof | Proficient | Transverse | Adenocarcinoma | Moderately differentiated | 0.05 |
| C140 | OCCFR | Female | 40-49 | MMRp control | CON | MMR-prof | Proficient | Caecum | Adenocarcinoma | Moderately differentiated | 0.09 |
| C141 | OCCFR | Female | 40-49 | MMRp control | CON | MMR-prof | Proficient | Sigmoid | Adenocarcinoma | Poorly differentiated | 0.19 |
| C142 | OCCFR | Male | 40-49 | MMRp control | CON | MMR-prof | Proficient | Sigmoid | Adenocarcinoma | Well differentiated | 0.07 |
| C143 | OCCFR | Female | 30-39 | MMRp control | CON | MMR-prof | Proficient | Rectum | Adenocarcinoma | Moderately differentiated | 0.06 |
| C144 | OCCFR | Female | 40-49 | MMRp control | CON | MMR-prof | Proficient | Rectum | Adenocarcinoma | Moderately differentiated | 0.06 |
| M12 | OCCFR | Female | 40-49 | MUTYH biallelic | MUTY H | p.Gly396Asp | Proficient | Caecum | Adenocarcinoma | Moderately differentiated | 0.00 |
| C145 | OCCFR | Female | 40-49 | MMRp control | CON | MMR-prof | Proficient | Ascending | Adenocarcinoma | Poorly differentiated | 0.05 |
| C146 | OCCFR | Male | 40-49 | MMRp control | CON | MMR-prof | Proficient | Sigmoid | Adenocarcinoma | Moderately differentiated | 0.03 |
| C147 | OCCFR | Male | 40-49 | MMRp control | CON | MMR-prof | Proficient | Rectum | Adenocarcinoma | Unknown | 0.03 |
| C148 | OCCFR | Male | 40-49 | MMRp control | CON | MMR-prof | Proficient | Caecum | Adenocarcinoma | Moderately differentiated | 0.13 |
| C149 | OCCFR | Female | 40-49 | MMRp control | CON | MMR-prof | Proficient | Rectum | Adenocarcinoma | Moderately differentiated | 0.06 |
| C150 | OCCFR | Female | 40-49 | MMRp control | CON | MMR-prof | Proficient | Sigmoid | Adenocarcinoma | Moderately differentiated | 0.00 |
| C151 | OCCFR | Male | 30-39 | MMRp control | CON | MMR-prof | Proficient | Caecum | Other | Well differentiated | 0.06 |
| C152 | OCCFR | Female | 40-49 | MMRp control | CON | MMR-prof | Proficient | Caecum | Adenocarcinoma | Moderately differentiated | 0.28 |
| C153 | OCCFR | Male | 30-39 | MMRp control | CON | MMR-prof | Proficient | Rectum | Adenocarcinoma | Moderately differentiated | 0.03 |
| C154 | OCCFR | Female | 40-49 | MMRp control | CON | MMR-prof | Proficient | Rectum | Adenocarcinoma | Moderately differentiated | 0.09 |

| | | | | | | | | | | | |
|---|---|---|---|---|---|---|---|---|---|---|---|
| C155 | OCCFR | Male | 40-49 | MMRp control | CON | MMR-prof | Proficient | Rectum | Adenocarcinoma | Moderately differentiated | 0.02 |
| C156 | OCCFR | Female | 40-49 | MMRp control | CON | MMR-prof | Proficient | Sigmoid | Mucinous | Well differentiated | 0.07 |
| C157 | OCCFR | Male | 40-49 | MMRp control | CON | MMR-prof | Proficient | Sigmoid | Adenocarcinoma | Well differentiated | 0.00 |
| C158 | OCCFR | Female | 40-49 | MMRp control | CON | MMR-prof | Proficient | Rectum | Adenocarcinoma | Unknown | 0.05 |
| C159 | OCCFR | Female | 40-49 | MMRp control | CON | MMR-prof | Proficient | Caecum | Unknown | Moderately differentiated | 0.07 |
| C160 | OCCFR | Female | 40-49 | MMRp control | CON | MMR-prof | Proficient | Rectum | Other | Poorly differentiated | 0.06 |
| C161 | OCCFR | Female | 40-49 | MMRp control | CON | MMR-prof | Proficient | Rectum | Adenocarcinoma | Moderately differentiated | 0.06 |
| C162 | OCCFR | Male | 40-49 | MMRp control | CON | MMR-prof | Proficient | Rectum | Unknown | Moderately differentiated | 0.12 |
| K11 | OCCFR | Female | 40-49 | MMRd control | MLH1 | MLH1 methylation | Deficient | Ascending | Adenocarcinoma | Poorly differentiated | 23.88 |
| K12 | OCCFR | Male | 40-49 | MMRd control | MLH1 | MLH1 methylation | Deficient | Ascending | Adenocarcinoma | Poorly differentiated | 9.72 |
| K13 | WEHI | Female | 70-79 | MMRd control | MLH1 | MLH1 methylation | Deficient | Transverse | Adenocarcioma | Well/Moderate | 13.57 |
| K14 | WEHI | Male | 70-79 | MMRd control | MLH1 | MLH1 methylation | Deficient | Ascending | Adenocarcioma | Poor | 20.27 |
| K15 | WEHI | Female | 80-89 | MMRd control | MLH1 | MLH1 methylation | Deficient | Caecum | Adenocarcioma | Poor | 22.38 |
| K16 | WEHI | Female | 90-99 | MMRd control | MLH1 | MLH1 methylation | Deficient | Caecum | Adenocarcioma | Well/Moderate | 21.76 |
| K17 | WEHI | Female | 60-69 | MMRd control | MLH1 | MLH1 methylation | Deficient | Hepatic flexure | Adenocarcioma | Well/Moderate | 17.76 |
| K18 | WEHI | Female | 70-79 | MMRd control | MLH1 | MLH1 methylation | Deficient | Ascending | Adenocarcioma | Poor | 10.34 |
| K19 | WEHI | Female | 60-69 | MMRd control | MLH1 | MLH1 methylation | Deficient | Caecum | Adenocarcioma | Well/Moderate | 18.37 |
| K20 | WEHI | Male | 60-69 | MMRd control | MLH1 | MLH1 methylation | Deficient | Transverse | Adenocarcioma | Poor | 7.80 |
| K21 | WEHI | Female | 60-69 | MMRd control | MLH1 | MLH1 methylation | Deficient | Ascending | Adenocarcioma | Well/Moderate | 6.11 |
| K22 | WEHI | Male | 70-79 | MMRd control | MLH1 | MLH1 methylation | Deficient | Ascending | Adenocarcioma | Poor | 19.50 |
| K23 | WEHI | Female | 70-79 | MMRd control | MLH1 | MLH1 methylation | Deficient | Ascending | Adenocarcioma | Poor | 9.84 |
| K24 | WEHI | Female | 70-79 | MMRd control | MLH1 | MLH1 methylation | Deficient | Hepatic flexure | Adenocarcioma | Poor | 19.35 |

| | | | | | | | | | | | |
|---|---|---|---|---|---|---|---|---|---|---|---|
| K25 | WEHI | Female | 60-69 | MMRd control | MLH1 | MLH1 methylation | Deficient | Transverse | Adenocarcioma | Well/Moderate | 13.61 |
| L25 | WEHI | Female | 60-69 | Lynch | MSH6 | c.2535dup | Deficient | Splenic flexure | Adenocarcioma | Well/Moderate | 4.35 |
| L26 | WEHI | Male | 50-59 | Lynch | MSH6 | c.4001G>A | Deficient | Hepatic flexure | Adenocarcioma | Well/Moderate | 3.80 |
| L12 | OCCFR | Male | 30-39 | Lynch | MSH2 | c.1786_1788del | Deficient | Caecum | Adenocarcinoma | Moderately differentiated | 22.14 |

**Supplementary Table 2.** Clinical data obtained from TCGA for the colon adenocarcinomas (COAD) and rectal adenocarcinomas (READ) tumours included in this study. MMR status (proficient or deficient) was determined from published MSI status generated by MSIseq [11]. MMRd CRCs were all determined to result from hypermethylation of the *MLH1* gene promoter as described in the **Supplementary Methods.**

| bcr_patient_barcode | MMR Status | AgeDx | TNM Stage | Anatomical Site | Sex | Histology | ICD | MSIseq |
|---|---|---|---|---|---|---|---|---|
| TCGA-3L-AA1B | Proficient | 61 | T2 | Cecum | FEMALE | Colon Adenocarcinoma | C18.0 | 0.07 |
| TCGA-4N-A93T | Proficient | 67 | T4a | Ascending Colon | MALE | Colon Adenocarcinoma | C18.2 | 0.07 |
| TCGA-4T-AA8H | Proficient | 42 | T3 | Descending Colon | FEMALE | Colon Mucinous Adenocarcinoma | C18.6 | 0.04 |
| TCGA-5M-AAT4 | Proficient | 74 | T3 | Ascending Colon | MALE | Colon Adenocarcinoma | C18.2 | 0.07 |
| TCGA-5M-AAT5 | Proficient | NA | NA | NA | FEMALE | Colon Adenocarcinoma | NA | 0.02 |
| TCGA-5M-AATA | Proficient | NA | NA | NA | MALE | Colon Adenocarcinoma | NA | 0.07 |
| TCGA-5M-AATE | Proficient | 76 | T3 | Ascending Colon | MALE | Colon Adenocarcinoma | C18.2 | 0.02 |
| TCGA-A6-2671 | Proficient | 85 | T3 | Sigmoid Colon | MALE | Colon Adenocarcinoma | C18.7 | 0.09 |
| TCGA-A6-2674 | Proficient | 71 | T3 | Sigmoid Colon | MALE | Colon Mucinous Adenocarcinoma | C18.7 | 0.00 |
| TCGA-A6-2675 | Proficient | 78 | T3 | Sigmoid Colon | MALE | Colon Adenocarcinoma | C18.7 | 0.00 |
| TCGA-A6-2677 | Proficient | 68 | T3 | Cecum | FEMALE | Colon Adenocarcinoma | C18.2 | 0.02 |
| TCGA-A6-2679 | Proficient | 73 | T3 | Ascending Colon | FEMALE | Colon Adenocarcinoma | C18.2 | 0.04 |
| TCGA-A6-2680 | Proficient | 72 | T3 | Hepatic Flexure | FEMALE | Colon Adenocarcinoma | C18.3 | 0.00 |
| TCGA-A6-2681 | Proficient | 73 | T3 | Cecum | FEMALE | Colon Adenocarcinoma | C18.9 | 0.02 |
| TCGA-A6-2682 | Proficient | 70 | T4b | NA | MALE | Colon Adenocarcinoma | C18.9 | 0.02 |
| TCGA-A6-2684 | Proficient | 75 | T2 | Cecum | FEMALE | Colon Adenocarcinoma | C18.9 | 0.04 |
| TCGA-A6-2685 | Proficient | 48 | T3 | Sigmoid Colon | FEMALE | Colon Adenocarcinoma | C18.9 | 0.00 |
| TCGA-A6-3807 | Proficient | 53 | T3 | Sigmoid Colon | FEMALE | Colon Adenocarcinoma | C18.7 | 0.07 |
| TCGA-A6-3808 | Proficient | 73 | T3 | Cecum | MALE | Colon Adenocarcinoma | C18.9 | 0.09 |
| TCGA-A6-3810 | Proficient | 62 | T3 | Sigmoid Colon | MALE | Colon Adenocarcinoma | C18.9 | 0.00 |
| TCGA-A6-4105 | Proficient | 79 | T3 | Ascending Colon | MALE | Colon Adenocarcinoma | C18.2 | 0.02 |
| TCGA-A6-4107 | Proficient | 57 | T3 | Ascending Colon | FEMALE | Colon Adenocarcinoma | C18.2 | 0.04 |
| TCGA-A6-5656 | Proficient | 74 | T2 | Sigmoid Colon | MALE | Colon Adenocarcinoma | C18.7 | 0.04 |
| TCGA-A6-5657 | Proficient | 65 | T3 | NA | MALE | Colon Adenocarcinoma | C18.9 | 0.00 |
| TCGA-A6-5659 | Proficient | 82 | T2 | Cecum | MALE | Colon Adenocarcinoma | C18.2 | 0.07 |
| TCGA-A6-5660 | Proficient | 73 | T3 | Cecum | MALE | Colon Adenocarcinoma | C18.2 | 0.07 |

| | | | | | | | | |
|---|---|---|---|---|---|---|---|---|
| TCGA-A6-5664 | Proficient | 80 | T4a | Cecum | MALE | Colon Adenocarcinoma | C18.2 | 0.00 |
| TCGA-A6-5666 | Proficient | 78 | T4b | Sigmoid Colon | MALE | Colon Adenocarcinoma | C18.9 | 0.09 |
| TCGA-A6-5667 | Proficient | 40 | T3 | Sigmoid Colon | FEMALE | Colon Adenocarcinoma | C18.7 | 0.09 |
| TCGA-A6-6137 | Proficient | 55 | T3 | Hepatic Flexure | MALE | Colon Adenocarcinoma | C18.2 | 0.07 |
| TCGA-A6-6138 | Proficient | 61 | T2 | Cecum | MALE | Colon Adenocarcinoma | C18.0 | 0.04 |
| TCGA-A6-6142 | Proficient | 56 | T3 | Sigmoid Colon | FEMALE | Colon Adenocarcinoma | C18.7 | 0.02 |
| TCGA-A6-6648 | Proficient | 56 | T3 | NA | MALE | Colon Adenocarcinoma | C18.6 | 0.00 |
| TCGA-A6-6649 | Proficient | 66 | T3 | NA | MALE | Colon Adenocarcinoma | C18.2 | 0.02 |
| TCGA-A6-6650 | Proficient | 69 | T3 | NA | FEMALE | Colon Adenocarcinoma | C18.2 | 0.09 |
| TCGA-A6-6651 | Proficient | 55 | T3 | Transverse Colon | FEMALE | Colon Adenocarcinoma | C18.9 | 0.04 |
| TCGA-A6-6652 | Proficient | 59 | T3 | Sigmoid Colon | MALE | Colon Adenocarcinoma | C18.7 | 0.07 |
| TCGA-A6-6654 | Proficient | 65 | T3 | Descending Colon | FEMALE | Colon Adenocarcinoma | C18.9 | 0.04 |
| TCGA-A6-6782 | Proficient | 82 | T4a | NA | MALE | Colon Adenocarcinoma | C18.2 | 0.02 |
| TCGA-A6-A566 | Proficient | 55 | T4 | Descending Colon | FEMALE | Colon Mucinous Adenocarcinoma | C18.6 | 0.07 |
| TCGA-A6-A567 | Proficient | 56 | T3 | Sigmoid Colon | MALE | Colon Adenocarcinoma | C18.7 | 0.02 |
| TCGA-A6-A56B | Proficient | 57 | T3 | Sigmoid Colon | MALE | Colon Adenocarcinoma | C18.7 | 0.04 |
| TCGA-AA-3488 | Proficient | 58 | T3 | Sigmoid Colon | MALE | Colon Adenocarcinoma | C19 | 0.02 |
| TCGA-AA-3489 | Proficient | 75 | T3 | Sigmoid Colon | MALE | Colon Adenocarcinoma | C18.9 | 0.04 |
| TCGA-AA-3494 | Proficient | 55 | T3 | Sigmoid Colon | MALE | Colon Adenocarcinoma | C18.9 | 0.04 |
| TCGA-AA-3495 | Proficient | 79 | T2 | Hepatic Flexure | MALE | Colon Adenocarcinoma | C18.2 | 0.02 |
| TCGA-AA-3496 | Proficient | 83 | T3 | Ascending Colon | FEMALE | Colon Adenocarcinoma | C18.2 | 0.13 |
| TCGA-AA-3502 | Proficient | 73 | T2 | Transverse Colon | MALE | Colon Adenocarcinoma | C18.9 | 0.09 |
| TCGA-AA-3506 | Proficient | 77 | T2 | Hepatic Flexure | MALE | Colon Adenocarcinoma | C18.9 | 0.13 |
| TCGA-AA-3509 | Proficient | 54 | T3 | Sigmoid Colon | FEMALE | Colon Adenocarcinoma | C18.7 | 0.04 |
| TCGA-AA-3510 | Proficient | 70 | T3 | Transverse Colon | MALE | Colon Adenocarcinoma | C18.2 | 0.11 |
| TCGA-AA-3511 | Proficient | 64 | T4 | Sigmoid Colon | MALE | Colon Adenocarcinoma | C18.9 | 0.02 |
| TCGA-AA-3530 | Proficient | 80 | T2 | Cecum | MALE | Colon Adenocarcinoma | C18.2 | 0.07 |
| TCGA-AA-3655 | Proficient | 68 | T3 | Sigmoid Colon | MALE | Colon Adenocarcinoma | C18.7 | 0.02 |
| TCGA-AA-3660 | Proficient | 51 | T3 | Sigmoid Colon | FEMALE | Colon Adenocarcinoma | C18.9 | 0.09 |
| TCGA-AA-3662 | Proficient | 80 | T4 | Sigmoid Colon | FEMALE | Colon Adenocarcinoma | C18.9 | 0.02 |
| TCGA-AA-3664 | Proficient | 74 | T3 | Cecum | FEMALE | Colon Adenocarcinoma | C18.2 | 0.04 |
| TCGA-AA-3666 | Proficient | 68 | T3 | Cecum | MALE | Colon Adenocarcinoma | C18.9 | 0.00 |
| TCGA-AA-3667 | Proficient | 36 | T2 | Sigmoid Colon | FEMALE | Colon Adenocarcinoma | C18.9 | 0.02 |
| TCGA-AA-3673 | Proficient | 53 | T3 | Transverse Colon | FEMALE | Colon Adenocarcinoma | C18.4 | 0.09 |
| TCGA-AA-3675 | Proficient | 84 | T3 | Hepatic Flexure | MALE | Colon Adenocarcinoma | C18.2 | 0.09 |
| TCGA-AA-3678 | Proficient | 60 | T2 | Sigmoid Colon | FEMALE | Colon Adenocarcinoma | C18.7 | 0.04 |

| | | | | | | | | |
|---|---|---|---|---|---|---|---|---|
| TCGA-AA-3679 | Proficient | 59 | T3 | Descending Colon | MALE | Colon Adenocarcinoma | C18.9 | 0.00 |
| TCGA-AA-3680 | Proficient | 67 | T4 | Cecum | FEMALE | Colon Adenocarcinoma | C18.0 | 0.02 |
| TCGA-AA-3681 | Proficient | 77 | T3 | Cecum | FEMALE | Colon Adenocarcinoma | C18.0 | 0.09 |
| TCGA-AA-3684 | Proficient | 65 | T4 | Cecum | FEMALE | Colon Mucinous Adenocarcinoma | C18.0 | 0.02 |
| TCGA-AA-3685 | Proficient | 69 | T3 | Sigmoid Colon | MALE | Colon Adenocarcinoma | C18.9 | 0.02 |
| TCGA-AA-3688 | Proficient | 80 | T3 | Sigmoid Colon | MALE | Colon Adenocarcinoma | C18.9 | 0.02 |
| TCGA-AA-3692 | Proficient | 47 | T3 | Splenic Flexure | FEMALE | Colon Mucinous Adenocarcinoma | C18.9 | 0.02 |
| TCGA-AA-3693 | Proficient | 77 | T4 | Sigmoid Colon | FEMALE | Colon Adenocarcinoma | C18.7 | 0.00 |
| TCGA-AA-3695 | Proficient | 63 | T3 | Cecum | FEMALE | Colon Adenocarcinoma | C18.0 | 0.09 |
| TCGA-AA-3696 | Proficient | 75 | T3 | Sigmoid Colon | FEMALE | Colon Adenocarcinoma | C18.7 | 0.07 |
| TCGA-AA-3697 | Proficient | 77 | T3 | Sigmoid Colon | MALE | Colon Adenocarcinoma | C18.9 | 0.13 |
| TCGA-AA-3712 | Proficient | 65 | T3 | Descending Colon | MALE | Colon Adenocarcinoma | C18.6 | 0.07 |
| TCGA-AA-3812 | Proficient | 82 | T3 | Sigmoid Colon | FEMALE | Colon Adenocarcinoma | C18.7 | 0.04 |
| TCGA-AA-3814 | Proficient | 85 | T3 | Ascending Colon | FEMALE | Colon Adenocarcinoma | C18.9 | 0.02 |
| TCGA-AA-3818 | Proficient | 78 | T3 | Hepatic Flexure | FEMALE | Colon Adenocarcinoma | C18.9 | 0.11 |
| TCGA-AA-3819 | Proficient | 41 | T3 | Sigmoid Colon | FEMALE | Colon Adenocarcinoma | C18.9 | 0.02 |
| TCGA-AA-3831 | Proficient | 66 | T3 | Sigmoid Colon | MALE | Colon Adenocarcinoma | C18.9 | 0.00 |
| TCGA-AA-3837 | Proficient | 67 | T3 | Hepatic Flexure | MALE | Colon Mucinous Adenocarcinoma | C18.9 | 0.13 |
| TCGA-AA-3841 | Proficient | 66 | T3 | Sigmoid Colon | MALE | Colon Adenocarcinoma | C18.9 | 0.04 |
| TCGA-AA-3842 | Proficient | 51 | T2 | Sigmoid Colon | MALE | Colon Adenocarcinoma | C18.9 | 0.04 |
| TCGA-AA-3844 | Proficient | 78 | T3 | Sigmoid Colon | FEMALE | Colon Adenocarcinoma | C18.9 | 0.00 |
| TCGA-AA-3846 | Proficient | 74 | T3 | Sigmoid Colon | FEMALE | Colon Adenocarcinoma | C18.7 | 0.04 |
| TCGA-AA-3848 | Proficient | 82 | T3 | Sigmoid Colon | FEMALE | Colon Adenocarcinoma | C18.7 | 0.02 |
| TCGA-AA-3850 | Proficient | 74 | T2 | Transverse Colon | MALE | Colon Adenocarcinoma | C18.4 | 0.02 |
| TCGA-AA-3851 | Proficient | 74 | T3 | Ascending Colon | MALE | Colon Adenocarcinoma | C18.9 | 0.18 |
| TCGA-AA-3852 | Proficient | 88 | T3 | Transverse Colon | MALE | Colon Mucinous Adenocarcinoma | C18.9 | 0.11 |
| TCGA-AA-3854 | Proficient | 71 | T2 | Sigmoid Colon | FEMALE | Colon Mucinous Adenocarcinoma | C18.9 | 0.02 |
| TCGA-AA-3855 | Proficient | 72 | T2 | Sigmoid Colon | MALE | Colon Adenocarcinoma | C18.9 | 0.02 |
| TCGA-AA-3856 | Proficient | 59 | T3 | Sigmoid Colon | MALE | Colon Adenocarcinoma | C18.9 | 0.02 |
| TCGA-AA-3858 | Proficient | 67 | T2 | Sigmoid Colon | MALE | Colon Adenocarcinoma | C18.9 | 0.02 |
| TCGA-AA-3860 | Proficient | 53 | T3 | Sigmoid Colon | FEMALE | Colon Adenocarcinoma | C18.7 | 0.02 |
| TCGA-AA-3861 | Proficient | 72 | T3 | Cecum | MALE | Colon Adenocarcinoma | C18.0 | 0.11 |
| TCGA-AA-3862 | Proficient | 82 | T3 | Transverse Colon | MALE | Colon Adenocarcinoma | C18.2 | 0.00 |
| TCGA-AA-3866 | Proficient | 78 | T2 | Cecum | FEMALE | Colon Adenocarcinoma | C18.9 | 0.02 |
| TCGA-AA-3867 | Proficient | 74 | T3 | Sigmoid Colon | MALE | Colon Adenocarcinoma | C18.7 | 0.02 |
| TCGA-AA-3869 | Proficient | 76 | T4 | Cecum | MALE | Colon Adenocarcinoma | C18.0 | 0.04 |

| | | | | | | | | |
|---|---|---|---|---|---|---|---|---|
| TCGA-AA-3870 | Proficient | 71 | T3 | Ascending Colon | FEMALE | Colon Adenocarcinoma | C18.2 | 0.04 |
| TCGA-AA-3872 | Proficient | 45 | T4 | Sigmoid Colon | MALE | Colon Adenocarcinoma | C18.9 | 0.07 |
| TCGA-AA-3875 | Proficient | 78 | T1 | Ascending Colon | FEMALE | Colon Adenocarcinoma | C18.2 | 0.02 |
| TCGA-AA-3930 | Proficient | 66 | T3 | Ascending Colon | MALE | Colon Adenocarcinoma | C18.2 | 0.09 |
| TCGA-AA-3939 | Proficient | 83 | T3 | Ascending Colon | MALE | Colon Adenocarcinoma | C18.9 | 0.07 |
| TCGA-AA-3941 | Proficient | 84 | T4 | Ascending Colon | FEMALE | Colon Adenocarcinoma | C18.9 | 0.11 |
| TCGA-AA-3952 | Proficient | 68 | T3 | Descending Colon | MALE | Colon Adenocarcinoma | C18.6 | 0.02 |
| TCGA-AA-3955 | Proficient | 38 | T2 | Descending Colon | MALE | Colon Adenocarcinoma | C18.9 | 0.02 |
| TCGA-AA-3956 | Proficient | 65 | T3 | Cecum | MALE | Colon Adenocarcinoma | C18.0 | 0.07 |
| TCGA-AA-3967 | Proficient | 77 | T3 | Sigmoid Colon | MALE | Colon Adenocarcinoma | C18.9 | 0.02 |
| TCGA-AA-3968 | Proficient | 55 | T2 | Sigmoid Colon | FEMALE | Colon Adenocarcinoma | C18.7 | 0.07 |
| TCGA-AA-3971 | Proficient | 58 | T3 | Sigmoid Colon | MALE | Colon Adenocarcinoma | C18.9 | 0.00 |
| TCGA-AA-3972 | Proficient | 72 | T3 | Sigmoid Colon | MALE | Colon Adenocarcinoma | C18.7 | 0.09 |
| TCGA-AA-3973 | Proficient | 69 | T4 | Sigmoid Colon | MALE | Colon Adenocarcinoma | C18.9 | 0.07 |
| TCGA-AA-3975 | Proficient | 80 | T2 | Sigmoid Colon | MALE | Colon Adenocarcinoma | C18.9 | 0.02 |
| TCGA-AA-3976 | Proficient | 70 | T2 | Sigmoid Colon | MALE | Colon Adenocarcinoma | C19 | 0.02 |
| TCGA-AA-3979 | Proficient | 84 | T3 | Sigmoid Colon | MALE | Colon Adenocarcinoma | C18.7 | 0.02 |
| TCGA-AA-3980 | Proficient | 89 | T2 | Sigmoid Colon | FEMALE | Colon Adenocarcinoma | C18.9 | 0.04 |
| TCGA-AA-3982 | Proficient | 75 | T3 | Sigmoid Colon | MALE | Colon Adenocarcinoma | C18.9 | 0.02 |
| TCGA-AA-3986 | Proficient | 73 | T2 | Transverse Colon | MALE | Colon Adenocarcinoma | C18.9 | 0.04 |
| TCGA-AA-3989 | Proficient | 84 | T3 | Transverse Colon | MALE | Colon Adenocarcinoma | C18.9 | 0.00 |
| TCGA-AA-3994 | Proficient | 69 | T3 | Transverse Colon | MALE | Colon Mucinous Adenocarcinoma | C18.9 | 0.07 |
| TCGA-AA-A004 | Proficient | 76 | T3 | Sigmoid Colon | MALE | Colon Adenocarcinoma | C18.7 | 0.00 |
| TCGA-AA-A017 | Proficient | 57 | T3 | Sigmoid Colon | FEMALE | Colon Adenocarcinoma | C18.7 | 0.07 |
| TCGA-AA-A01C | Proficient | 75 | T2 | Ascending Colon | MALE | Colon Adenocarcinoma | C18.2 | 0.00 |
| TCGA-AA-A01I | Proficient | 76 | T2 | Sigmoid Colon | MALE | Colon Adenocarcinoma | C18.7 | 0.02 |
| TCGA-AA-A01S | Proficient | 47 | T3 | Sigmoid Colon | FEMALE | Colon Adenocarcinoma | C18.7 | 0.04 |
| TCGA-AA-A01T | Proficient | 63 | T3 | Sigmoid Colon | FEMALE | Colon Adenocarcinoma | C18.7 | 0.02 |
| TCGA-AA-A01V | Proficient | 59 | T2 | Cecum | MALE | Colon Adenocarcinoma | C18.0 | 0.02 |
| TCGA-AA-A01X | Proficient | 80 | T2 | Sigmoid Colon | FEMALE | Colon Adenocarcinoma | C18.7 | 0.00 |
| TCGA-AA-A01Z | Proficient | 68 | T3 | Ascending Colon | MALE | Colon Adenocarcinoma | C18.2 | 0.02 |
| TCGA-AA-A024 | Proficient | 81 | T3 | Descending Colon | MALE | Colon Mucinous Adenocarcinoma | C18.6 | 0.00 |
| TCGA-AA-A02E | Proficient | 82 | T3 | Cecum | FEMALE | Colon Adenocarcinoma | C18.0 | 0.07 |
| TCGA-AA-A02F | Proficient | 68 | T3 | Sigmoid Colon | FEMALE | Colon Adenocarcinoma | C18.7 | 0.02 |
| TCGA-AA-A02H | Proficient | 74 | T3 | Sigmoid Colon | FEMALE | Colon Adenocarcinoma | C18.7 | 0.00 |
| TCGA-AA-A02K | Proficient | 50 | T4 | Ascending Colon | MALE | Colon Adenocarcinoma | C18.2 | 0.11 |

| | | | | | | | | |
|---|---|---|---|---|---|---|---|---|
| TCGA-AA-A02O | Proficient | 82 | T3 | Transverse Colon | MALE | Colon Adenocarcinoma | C18.4 | 0.00 |
| TCGA-AA-A02W | Proficient | 73 | T2 | Sigmoid Colon | FEMALE | Colon Adenocarcinoma | C18.7 | 0.00 |
| TCGA-AA-A02Y | Proficient | 73 | T2 | Cecum | MALE | Colon Adenocarcinoma | C18.0 | 0.02 |
| TCGA-AA-A03F | Proficient | 90 | T3 | Cecum | FEMALE | Colon Mucinous Adenocarcinoma | C18.0 | 0.00 |
| TCGA-AA-A03J | Proficient | 65 | T2 | Sigmoid Colon | FEMALE | Colon Adenocarcinoma | C18.7 | 0.02 |
| TCGA-AD-6548 | Proficient | 81 | T2 | Splenic Flexure | FEMALE | Colon Adenocarcinoma | C18.5 | 0.02 |
| TCGA-AD-6888 | Proficient | 73 | T3 | Hepatic Flexure | MALE | Colon Adenocarcinoma | C18.2 | 0.04 |
| TCGA-AD-6890 | Proficient | 65 | T1 | Ascending Colon | MALE | Colon Adenocarcinoma | C18.2 | 0.11 |
| TCGA-AD-6899 | Proficient | 84 | T4a | Cecum | MALE | Colon Mucinous Adenocarcinoma | C18.9 | 0.02 |
| TCGA-AD-6901 | Proficient | 78 | T3 | Cecum | MALE | Colon Adenocarcinoma | C18.0 | 0.04 |
| TCGA-AD-6963 | Proficient | 58 | T3 | Ascending Colon | MALE | Colon Adenocarcinoma | C18.2 | 0.02 |
| TCGA-AD-6965 | Proficient | 62 | T4a | Cecum | MALE | Colon Adenocarcinoma | C18.0 | 0.07 |
| TCGA-AD-A5EK | Proficient | 51 | T2 | Ascending Colon | MALE | Colon Adenocarcinoma | C18.2 | 0.02 |
| TCGA-AU-3779 | Proficient | 80 | T3 | Rectosigmoid Junction | FEMALE | Colon Adenocarcinoma | C18.7 | 0.00 |
| TCGA-AY-4070 | Proficient | 50 | T3 | Cecum | FEMALE | Colon Adenocarcinoma | C18.2 | 0.02 |
| TCGA-AY-4071 | Proficient | 63 | T1 | [Not Available] | FEMALE | [Not Available] | C18.7 | 0.00 |
| TCGA-AY-5543 | Proficient | 65 | T3 | Ascending Colon | FEMALE | Colon Adenocarcinoma | C18.2 | 0.07 |
| TCGA-AY-6196 | Proficient | 47 | T3 | Cecum | MALE | Colon Mucinous Adenocarcinoma | C18.2 | 0.09 |
| TCGA-AY-6386 | Proficient | 66 | T3 | Cecum | FEMALE | Colon Adenocarcinoma | C18.0 | 0.16 |
| TCGA-AY-A54L | Proficient | 74 | T2 | Transverse Colon | FEMALE | Colon Adenocarcinoma | C18.3 | 0.11 |
| TCGA-AY-A69D | Proficient | 55 | T3 | Transverse Colon | FEMALE | Colon Adenocarcinoma | C18.4 | 0.07 |
| TCGA-AY-A71X | Proficient | 54 | T2 | Cecum | FEMALE | Colon Adenocarcinoma | C18.0 | 0.09 |
| TCGA-AY-A8YK | Proficient | 44 | T3 | Sigmoid Colon | MALE | Colon Adenocarcinoma | C18.7 | 0.00 |
| TCGA-AZ-4308 | Proficient | 47 | T3 | Sigmoid Colon | FEMALE | Colon Adenocarcinoma | C18.7 | 0.00 |
| TCGA-AZ-4323 | Proficient | 37 | T4 | Cecum | MALE | Colon Adenocarcinoma | C18.0 | 0.02 |
| TCGA-AZ-4614 | Proficient | 71 | T4a | [Not Available] | FEMALE | Colon Adenocarcinoma | C18.0 | 0.07 |
| TCGA-AZ-4616 | Proficient | 82 | T3 | Cecum | FEMALE | Colon Adenocarcinoma | C18.0 | 0.04 |
| TCGA-AZ-4681 | Proficient | 79 | T3 | Ascending Colon | FEMALE | Colon Adenocarcinoma | C18.2 | 0.04 |
| TCGA-AZ-4682 | Proficient | 61 | T3 | Sigmoid Colon | MALE | Colon Adenocarcinoma | C19 | 0.02 |
| TCGA-AZ-4684 | Proficient | 49 | T3 | [Not Available] | MALE | Colon Adenocarcinoma | C19 | 0.00 |
| TCGA-AZ-5403 | Proficient | 43 | T3 | Sigmoid Colon | MALE | Colon Adenocarcinoma | C18.6 | 0.00 |
| TCGA-AZ-5407 | Proficient | 51 | T1 | Cecum | FEMALE | Colon Adenocarcinoma | C18.0 | 0.00 |
| TCGA-AZ-6599 | Proficient | 72 | T2 | Cecum | MALE | Colon Adenocarcinoma | C18.0 | 0.04 |
| TCGA-AZ-6600 | Proficient | 64 | T4 | Hepatic Flexure | MALE | Colon Adenocarcinoma | C18.2 | 0.13 |
| TCGA-AZ-6603 | Proficient | 77 | T2 | Sigmoid Colon | FEMALE | Colon Adenocarcinoma | C18.7 | 0.09 |
| TCGA-AZ-6605 | Proficient | 77 | T4 | Ascending Colon | MALE | Colon Adenocarcinoma | C18.2 | 0.13 |

| | | | | | | | | |
|---|---|---|---|---|---|---|---|---|
| TCGA-AZ-6606 | Proficient | 81 | T4 | Cecum | MALE | Colon Adenocarcinoma | C18.0 | 0.00 |
| TCGA-AZ-6607 | Proficient | 69 | T4 | Sigmoid Colon | MALE | Colon Adenocarcinoma | C18.7 | 0.04 |
| TCGA-AZ-6608 | Proficient | 55 | T2 | Sigmoid Colon | FEMALE | Colon Adenocarcinoma | C18.7 | 0.11 |
| TCGA-CA-5254 | Proficient | 42 | T3 | Transverse Colon | FEMALE | Colon Adenocarcinoma | C18.9 | 0.07 |
| TCGA-CA-5255 | Proficient | 45 | T3 | Ascending Colon | MALE | Colon Adenocarcinoma | C18.9 | 0.00 |
| TCGA-CA-5256 | Proficient | 54 | T3 | Hepatic Flexure | FEMALE | Colon Adenocarcinoma | C18.9 | 0.07 |
| TCGA-CA-5796 | Proficient | 52 | T3 | Ascending Colon | FEMALE | Colon Mucinous Adenocarcinoma | C18.2 | 0.09 |
| TCGA-CA-5797 | Proficient | 56 | T3 | Sigmoid Colon | MALE | Colon Adenocarcinoma | C18.9 | 0.04 |
| TCGA-CA-6715 | Proficient | 63 | T3 | Sigmoid Colon | MALE | Colon Adenocarcinoma | C18.7 | 0.04 |
| TCGA-CA-6716 | Proficient | 65 | T3 | Ascending Colon | MALE | Colon Adenocarcinoma | C18.2 | 0.16 |
| TCGA-CA-6719 | Proficient | 77 | T3 | Descending Colon | MALE | Colon Adenocarcinoma | C18.6 | 0.07 |
| TCGA-CK-4947 | Proficient | 46 | T4 | Sigmoid Colon | FEMALE | Colon Adenocarcinoma | C18.7 | 0.07 |
| TCGA-CK-4948 | Proficient | 45 | T3 | Sigmoid Colon | FEMALE | Colon Adenocarcinoma | C18.7 | 0.04 |
| TCGA-CK-4950 | Proficient | 68 | T3 | Cecum | FEMALE | Colon Mucinous Adenocarcinoma | C18.0 | 0.13 |
| TCGA-CK-4952 | Proficient | 48 | T4 | Ascending Colon | FEMALE | Colon Mucinous Adenocarcinoma | C18.2 | 0.09 |
| TCGA-CK-5912 | Proficient | 81 | T2 | Cecum | MALE | Colon Adenocarcinoma | C18.2 | 0.02 |
| TCGA-CK-5914 | Proficient | 81 | T3 | Sigmoid Colon | MALE | Colon Adenocarcinoma | C18.7 | 0.07 |
| TCGA-CK-5915 | Proficient | 63 | T2 | Sigmoid Colon | MALE | Colon Adenocarcinoma | C18.7 | 0.07 |
| TCGA-CK-6748 | Proficient | 45 | T3 | Sigmoid Colon | FEMALE | Colon Mucinous Adenocarcinoma | C18.7 | 0.04 |
| TCGA-CK-6751 | Proficient | 88 | T2 | Ascending Colon | FEMALE | Colon Mucinous Adenocarcinoma | C18.2 | 0.07 |
| TCGA-CM-4744 | Proficient | 69 | T2 | Cecum | MALE | Colon Adenocarcinoma | C18.0 | 0.16 |
| TCGA-CM-4747 | Proficient | 47 | T4a | Cecum | MALE | Colon Adenocarcinoma | C18.0 | 0.02 |
| TCGA-CM-4748 | Proficient | 53 | T4a | Transverse Colon | MALE | Colon Mucinous Adenocarcinoma | C18.4 | 0.02 |
| TCGA-CM-4750 | Proficient | 34 | T1 | [Not Available] | FEMALE | Colon Adenocarcinoma | C19 | 0.09 |
| TCGA-CM-4751 | Proficient | 62 | T3 | Cecum | MALE | Colon Adenocarcinoma | C18.0 | 0.00 |
| TCGA-CM-4752 | Proficient | 58 | T3 | Ascending Colon | MALE | Colon Adenocarcinoma | C18.2 | 0.04 |
| TCGA-CM-5341 | Proficient | 82 | T2 | Sigmoid Colon | FEMALE | Colon Adenocarcinoma | C18.7 | 0.04 |
| TCGA-CM-5344 | Proficient | 39 | T3 | Sigmoid Colon | FEMALE | Colon Adenocarcinoma | C18.7 | 0.04 |
| TCGA-CM-5348 | Proficient | 72 | T3 | Cecum | MALE | Colon Adenocarcinoma | C18.0 | 0.00 |
| TCGA-CM-5349 | Proficient | 68 | T3 | Cecum | FEMALE | Colon Adenocarcinoma | C18.0 | 0.04 |
| TCGA-CM-5860 | Proficient | 44 | T3 | Ascending Colon | MALE | Colon Adenocarcinoma | C18.2 | 0.00 |
| TCGA-CM-5862 | Proficient | 80 | T3 | Ascending Colon | MALE | Colon Adenocarcinoma | C18.2 | 0.11 |
| TCGA-CM-5863 | Proficient | 60 | T3 | Ascending Colon | FEMALE | Colon Mucinous Adenocarcinoma | C18.2 | 0.02 |
| TCGA-CM-5864 | Proficient | 60 | T2 | Cecum | MALE | Colon Adenocarcinoma | C18.0 | 0.09 |
| TCGA-CM-5868 | Proficient | 59 | T4a | Sigmoid Colon | FEMALE | Colon Adenocarcinoma | C18.7 | 0.13 |
| TCGA-CM-6161 | Proficient | 36 | T2 | Sigmoid Colon | FEMALE | Colon Adenocarcinoma | C18.7 | 0.07 |

| | | | | | | | | |
|---|---|---|---|---|---|---|---|---|
| TCGA-CM-6163 | Proficient | 74 | T1 | Sigmoid Colon | MALE | Colon Adenocarcinoma | C18.7 | 0.02 |
| TCGA-CM-6164 | Proficient | 46 | T3 | Sigmoid Colon | FEMALE | Colon Adenocarcinoma | C18.7 | 0.09 |
| TCGA-CM-6165 | Proficient | 74 | T3 | Sigmoid Colon | MALE | Colon Adenocarcinoma | C18.7 | 0.00 |
| TCGA-CM-6166 | Proficient | 48 | T2 | Ascending Colon | FEMALE | Colon Adenocarcinoma | C18.2 | 0.00 |
| TCGA-CM-6167 | Proficient | 57 | T3 | Cecum | FEMALE | Colon Adenocarcinoma | C18.0 | 0.09 |
| TCGA-CM-6169 | Proficient | 67 | T3 | Cecum | MALE | Colon Adenocarcinoma | C18.0 | 0.02 |
| TCGA-CM-6170 | Proficient | 73 | T2 | Descending Colon | FEMALE | Colon Adenocarcinoma | C18.6 | 0.07 |
| TCGA-CM-6172 | Proficient | 70 | T3 | Sigmoid Colon | FEMALE | Colon Adenocarcinoma | C18.7 | 0.00 |
| TCGA-CM-6675 | Proficient | 35 | T3 | Cecum | MALE | Colon Adenocarcinoma | C18.0 | 0.02 |
| TCGA-CM-6676 | Proficient | 82 | T2 | Sigmoid Colon | MALE | Colon Adenocarcinoma | C18.7 | 0.04 |
| TCGA-CM-6677 | Proficient | 75 | T3 | Hepatic Flexure | FEMALE | Colon Adenocarcinoma | C18.3 | 0.09 |
| TCGA-CM-6678 | Proficient | 63 | T4a | Sigmoid Colon | FEMALE | Colon Adenocarcinoma | C18.7 | 0.02 |
| TCGA-CM-6679 | Proficient | 58 | T3 | Sigmoid Colon | MALE | Colon Adenocarcinoma | C18.7 | 0.02 |
| TCGA-CM-6680 | Proficient | 78 | T3 | Cecum | FEMALE | Colon Adenocarcinoma | C18.0 | 0.04 |
| TCGA-D5-5537 | Proficient | 83 | T3 | Ascending Colon | MALE | Colon Adenocarcinoma | C18.9 | 0.09 |
| TCGA-D5-5538 | Proficient | 60 | T3 | Cecum | FEMALE | Colon Adenocarcinoma | C18.0 | 0.11 |
| TCGA-D5-5539 | Proficient | 60 | T3 | Ascending Colon | MALE | Colon Mucinous Adenocarcinoma | C18.2 | 0.04 |
| TCGA-D5-5540 | Proficient | 73 | T3 | Cecum | MALE | Colon Adenocarcinoma | C18.0 | 0.00 |
| TCGA-D5-5541 | Proficient | 63 | T3 | Sigmoid Colon | MALE | Colon Adenocarcinoma | C18.7 | 0.02 |
| TCGA-D5-6529 | Proficient | 69 | T3 | NA | MALE | Colon Adenocarcinoma | C18.9 | 0.00 |
| TCGA-D5-6531 | Proficient | 75 | T3 | Hepatic Flexure | MALE | Colon Adenocarcinoma | C18.3 | 0.07 |
| TCGA-D5-6532 | Proficient | 61 | T3 | Sigmoid Colon | MALE | Colon Adenocarcinoma | C18.7 | 0.04 |
| TCGA-D5-6533 | Proficient | 68 | T4b | Transverse Colon | FEMALE | Colon Adenocarcinoma | C18.4 | 0.09 |
| TCGA-D5-6534 | Proficient | 62 | T3 | Ascending Colon | FEMALE | Colon Mucinous Adenocarcinoma | C18.9 | 0.02 |
| TCGA-D5-6535 | Proficient | 80 | T3 | Ascending Colon | FEMALE | Colon Adenocarcinoma | C18.9 | 0.02 |
| TCGA-D5-6536 | Proficient | 73 | T3 | Sigmoid Colon | MALE | Colon Adenocarcinoma | C18.9 | 0.00 |
| TCGA-D5-6537 | Proficient | 64 | T3 | Transverse Colon | MALE | Colon Adenocarcinoma | C18.4 | 0.02 |
| TCGA-D5-6538 | Proficient | 79 | T3 | Hepatic Flexure | FEMALE | Colon Adenocarcinoma | C18.3 | 0.13 |
| TCGA-D5-6539 | Proficient | 45 | T3 | Transverse Colon | FEMALE | Colon Adenocarcinoma | C18.4 | 0.02 |
| TCGA-D5-6541 | Proficient | 49 | T3 | Splenic Flexure | MALE | Colon Adenocarcinoma | C18.5 | 0.11 |
| TCGA-D5-6898 | Proficient | 51 | T2 | Sigmoid Colon | FEMALE | Colon Adenocarcinoma | C18.9 | 0.02 |
| TCGA-D5-6920 | Proficient | 77 | T3 | Sigmoid Colon | FEMALE | Colon Adenocarcinoma | C18.7 | 0.07 |
| TCGA-D5-6922 | Proficient | 76 | T3 | Sigmoid Colon | MALE | Colon Adenocarcinoma | C18.7 | 0.02 |
| TCGA-D5-6924 | Proficient | 68 | T3 | Sigmoid Colon | MALE | Colon Adenocarcinoma | C18.7 | 0.02 |
| TCGA-D5-6926 | Proficient | 65 | T4a | Sigmoid Colon | MALE | Colon Adenocarcinoma | C18.7 | 0.07 |
| TCGA-D5-6929 | Proficient | 49 | T3 | Sigmoid Colon | FEMALE | Colon Adenocarcinoma | C18.9 | 0.04 |

| | | | | | | | | |
|---|---|---|---|---|---|---|---|---|
| TCGA-D5-6931 | Proficient | 77 | T4b | Transverse Colon | MALE | Colon Adenocarcinoma | C18.9 | 0.07 |
| TCGA-D5-6932 | Proficient | 69 | T3 | Transverse Colon | MALE | Colon Adenocarcinoma | C18.9 | 0.02 |
| TCGA-D5-7000 | Proficient | 79 | T2 | Cecum | FEMALE | Colon Mucinous Adenocarcinoma | C18.0 | 0.04 |
| TCGA-DM-A0X9 | Proficient | 71 | T3 | NA | FEMALE | Colon Adenocarcinoma | C18.2 | 0.16 |
| TCGA-DM-A0XD | Proficient | 65 | T3 | NA | MALE | Colon Adenocarcinoma | C18.0 | 0.07 |
| TCGA-DM-A0XF | Proficient | 68 | T3 | NA | FEMALE | NA | C18.7 | 0.02 |
| TCGA-DM-A1D0 | Proficient | 79 | T3 | Sigmoid Colon | FEMALE | Colon Adenocarcinoma | C18.7 | 0.04 |
| TCGA-DM-A1D4 | Proficient | 80 | T3 | Cecum | MALE | Colon Adenocarcinoma | C18.0 | 0.11 |
| TCGA-DM-A1D6 | Proficient | 88 | T3 | Splenic Flexure | MALE | Colon Mucinous Adenocarcinoma | C18.5 | 0.02 |
| TCGA-DM-A1D7 | Proficient | 82 | T3 | Sigmoid Colon | MALE | NA | C18.7 | 0.04 |
| TCGA-DM-A1D8 | Proficient | 50 | T3 | Ascending Colon | FEMALE | Colon Adenocarcinoma | C18.2 | 0.09 |
| TCGA-DM-A1D9 | Proficient | 67 | T3 | Cecum | FEMALE | Colon Adenocarcinoma | C18.0 | 0.04 |
| TCGA-DM-A1DA | Proficient | 71 | T3 | Cecum | FEMALE | Colon Adenocarcinoma | C18.0 | 0.00 |
| TCGA-DM-A1DB | Proficient | 68 | T3 | Sigmoid Colon | MALE | Colon Adenocarcinoma | C18.7 | 0.07 |
| TCGA-DM-A1HA | Proficient | 82 | T3 | Ascending Colon | MALE | Colon Adenocarcinoma | C18.2 | 0.07 |
| TCGA-DM-A280 | Proficient | 70 | T3 | Ascending Colon | FEMALE | Colon Mucinous Adenocarcinoma | C18.2 | 0.13 |
| TCGA-DM-A282 | Proficient | 60 | T3 | Hepatic Flexure | FEMALE | Colon Adenocarcinoma | C18.3 | 0.07 |
| TCGA-DM-A285 | Proficient | 71 | T3 | Ascending Colon | FEMALE | Colon Mucinous Adenocarcinoma | C18.2 | 0.02 |
| TCGA-DM-A288 | Proficient | 68 | T3 | Cecum | MALE | Colon Mucinous Adenocarcinoma | C18.0 | 0.00 |
| TCGA-DM-A28A | Proficient | 78 | T3 | Cecum | MALE | Colon Adenocarcinoma | C18.0 | 0.07 |
| TCGA-DM-A28C | Proficient | 74 | T3 | Sigmoid Colon | MALE | Colon Adenocarcinoma | C18.7 | 0.04 |
| TCGA-DM-A28E | Proficient | 72 | T3 | Sigmoid Colon | FEMALE | Colon Adenocarcinoma | C18.7 | 0.02 |
| TCGA-DM-A28F | Proficient | 73 | T3 | Sigmoid Colon | MALE | Colon Adenocarcinoma | C18.7 | 0.07 |
| TCGA-DM-A28G | Proficient | 75 | T3 | Ascending Colon | MALE | Colon Adenocarcinoma | C18.2 | 0.02 |
| TCGA-DM-A28H | Proficient | 50 | T3 | Cecum | MALE | Colon Adenocarcinoma | C18.0 | 0.02 |
| TCGA-DM-A28K | Proficient | 75 | T3 | Hepatic Flexure | MALE | Colon Mucinous Adenocarcinoma | C18.3 | 0.11 |
| TCGA-DM-A28M | Proficient | 63 | T3 | Descending Colon | MALE | Colon Adenocarcinoma | C18.6 | 0.04 |
| TCGA-F4-6459 | Proficient | 61 | T3 | Sigmoid Colon | FEMALE | Colon Adenocarcinoma | C18.7 | 0.09 |
| TCGA-F4-6460 | Proficient | 51 | T3 | Sigmoid Colon | FEMALE | Colon Adenocarcinoma | C18.7 | 0.07 |
| TCGA-F4-6461 | Proficient | 41 | T4b | Hepatic Flexure | FEMALE | Colon Adenocarcinoma | C18.9 | 0.04 |
| TCGA-F4-6463 | Proficient | 51 | T3 | Transverse Colon | MALE | Colon Mucinous Adenocarcinoma | C18.4 | 0.02 |
| TCGA-F4-6569 | Proficient | 60 | T2 | Transverse Colon | MALE | Colon Adenocarcinoma | C18.9 | 0.02 |
| TCGA-F4-6704 | Proficient | 60 | T3 | Sigmoid Colon | MALE | Colon Mucinous Adenocarcinoma | C18.7 | 0.00 |
| TCGA-F4-6805 | Proficient | 58 | T3 | Descending Colon | FEMALE | Colon Adenocarcinoma | C18.9 | 0.00 |
| TCGA-F4-6806 | Proficient | 59 | T2 | Sigmoid Colon | FEMALE | Colon Adenocarcinoma | C18.7 | 0.07 |
| TCGA-F4-6807 | Proficient | 51 | T3 | Hepatic Flexure | FEMALE | Colon Adenocarcinoma | C18.9 | 0.02 |

| | | | | | | | | |
|---|---|---|---|---|---|---|---|---|
| TCGA-F4-6808 | Proficient | 54 | T1 | Sigmoid Colon | FEMALE | Colon Adenocarcinoma | C18.7 | 0.02 |
| TCGA-F4-6809 | Proficient | 52 | T3 | Sigmoid Colon | FEMALE | Colon Adenocarcinoma | C18.7 | 0.09 |
| TCGA-F4-6854 | Proficient | 77 | T3 | Sigmoid Colon | FEMALE | Colon Adenocarcinoma | C18.7 | 0.02 |
| TCGA-F4-6855 | Proficient | 70 | T3 | Sigmoid Colon | FEMALE | Colon Adenocarcinoma | C18.7 | 0.02 |
| TCGA-G4-6293 | Proficient | 49 | T3 | Transverse Colon | FEMALE | Colon Adenocarcinoma | C18.4 | 0.04 |
| TCGA-G4-6294 | Proficient | 75 | T3 | Cecum | MALE | Colon Adenocarcinoma | C18.0 | 0.02 |
| TCGA-G4-6295 | Proficient | 70 | T3 | Cecum | FEMALE | Colon Adenocarcinoma | C18.0 | 0.07 |
| TCGA-G4-6297 | Proficient | 55 | T3 | Cecum | FEMALE | Colon Adenocarcinoma | C18.0 | 0.11 |
| TCGA-G4-6298 | Proficient | 90 | T4a | Cecum | MALE | Colon Adenocarcinoma | C18.0 | 0.00 |
| TCGA-G4-6299 | Proficient | 69 | T3 | Descending Colon | MALE | Colon Adenocarcinoma | C18.6 | 0.04 |
| TCGA-G4-6303 | Proficient | 54 | T3 | Sigmoid Colon | FEMALE | Colon Adenocarcinoma | C18.7 | 0.13 |
| TCGA-G4-6306 | Proficient | 71 | T2 | Ascending Colon | MALE | Colon Adenocarcinoma | C18.2 | 0.04 |
| TCGA-G4-6307 | Proficient | 37 | T3 | Sigmoid Colon | FEMALE | Colon Adenocarcinoma | C18.7 | 0.00 |
| TCGA-G4-6310 | Proficient | 69 | T3 | Cecum | MALE | Colon Adenocarcinoma | C18.9 | 0.04 |
| TCGA-G4-6311 | Proficient | 80 | T3 | Ascending Colon | MALE | Colon Adenocarcinoma | C18.2 | 0.04 |
| TCGA-G4-6314 | Proficient | 76 | T3 | Cecum | FEMALE | Colon Adenocarcinoma | C18.0 | 0.02 |
| TCGA-G4-6315 | Proficient | 66 | T3 | Descending Colon | MALE | Colon Adenocarcinoma | C18.6 | 0.16 |
| TCGA-G4-6317 | Proficient | 51 | T3 | Sigmoid Colon | FEMALE | Colon Adenocarcinoma | C18.7 | 0.04 |
| TCGA-G4-6321 | Proficient | 60 | T2 | Cecum | FEMALE | Colon Adenocarcinoma | C18.0 | 0.09 |
| TCGA-G4-6322 | Proficient | 65 | T3 | Descending Colon | MALE | Colon Mucinous Adenocarcinoma | C18.6 | 0.02 |
| TCGA-G4-6323 | Proficient | 50 | Tis | Cecum | MALE | Colon Adenocarcinoma | C18.0 | 0.00 |
| TCGA-G4-6625 | Proficient | 77 | T3 | Sigmoid Colon | FEMALE | Colon Adenocarcinoma | C18.7 | 0.07 |
| TCGA-G4-6626 | Proficient | 90 | T3 | Ascending Colon | MALE | Colon Adenocarcinoma | C18.2 | 0.04 |
| TCGA-G4-6627 | Proficient | 84 | T3 | Ascending Colon | MALE | Colon Adenocarcinoma | C18.2 | 0.00 |
| TCGA-NH-A50T | Proficient | 68 | T3 | Splenic Flexure | FEMALE | Colon Adenocarcinoma | C18.5 | 0.04 |
| TCGA-NH-A50U | Proficient | 42 | T4a | Cecum | MALE | Colon Mucinous Adenocarcinoma | C18.0 | 0.02 |
| TCGA-NH-A50V | Proficient | 69 | T3 | Cecum | MALE | Colon Adenocarcinoma | C18.0 | 0.11 |
| TCGA-NH-A6GA | Proficient | 58 | T4a | Ascending Colon | MALE | Colon Adenocarcinoma | C18.2 | 0.02 |
| TCGA-NH-A6GB | Proficient | 71 | T3 | Transverse Colon | FEMALE | Colon Adenocarcinoma | C18.4 | 0.04 |
| TCGA-NH-A6GC | Proficient | 66 | T4b | Descending Colon | FEMALE | Colon Mucinous Adenocarcinoma | C18.6 | 0.18 |
| TCGA-NH-A8F7 | Proficient | 53 | T3 | Sigmoid Colon | FEMALE | Colon Adenocarcinoma | C18.7 | 0.02 |
| TCGA-NH-A8F8 | Proficient | 79 | T4a | Ascending Colon | MALE | Colon Adenocarcinoma | C18.2 | 0.00 |
| TCGA-QG-A5YV | Proficient | 64 | T4b | Sigmoid Colon | FEMALE | Colon Adenocarcinoma | C18.7 | 0.09 |
| TCGA-QG-A5YW | Proficient | 55 | T3 | Cecum | FEMALE | Colon Mucinous Adenocarcinoma | C18.0 | 0.07 |
| TCGA-QG-A5YX | Proficient | 61 | T3 | Sigmoid Colon | FEMALE | Colon Adenocarcinoma | C18.7 | 0.09 |
| TCGA-QG-A5Z1 | Proficient | 71 | T3 | Sigmoid Colon | MALE | Colon Adenocarcinoma | C19 | 0.02 |

| | | | | | | | | |
|---|---|---|---|---|---|---|---|---|
| TCGA-QL-A97D | Proficient | 84 | T2 | Cecum | FEMALE | Colon Adenocarcinoma | C18.2 | 0.07 |
| TCGA-RU-A8FL | Proficient | 51 | T3 | Cecum | MALE | Colon Adenocarcinoma | C18.0 | 0.00 |
| TCGA-SS-A7HO | Proficient | 44 | T4a | Cecum | FEMALE | Colon Adenocarcinoma | C18.0 | 0.09 |
| TCGA-T9-A92H | Proficient | 82 | T3 | Sigmoid Colon | MALE | Colon Adenocarcinoma | C18.7 | 0.07 |
| TCGA-AF-2687 | Proficient | 57 | T3 | Rectum | MALE | Rectal Adenocarcinoma | C19 | 0.02 |
| TCGA-AF-2690 | Proficient | 76 | T3 | Rectum | FEMALE | Rectal Adenocarcinoma | C20 | 0.00 |
| TCGA-AF-2693 | Proficient | 75 | T2 | Sigmoid Colon | MALE | [Not Available] | C19 | 0.00 |
| TCGA-AF-3911 | Proficient | 48 | T3 | Rectum | MALE | Rectal Adenocarcinoma | C20 | 0.04 |
| TCGA-AF-3913 | Proficient | 60 | T3 | Rectosigmoid Junction | MALE | Rectal Adenocarcinoma | C19 | 0.04 |
| TCGA-AF-3914 | Proficient | 39 | T3 | Rectosigmoid Junction | MALE | Rectal Adenocarcinoma | C19 | 0.04 |
| TCGA-AF-4110 | Proficient | 77 | T4a | Rectosigmoid Junction | MALE | [Not Available] | C20 | 0.00 |
| TCGA-AF-5654 | Proficient | 73 | T2 | Rectosigmoid Junction | FEMALE | Rectal Adenocarcinoma | C19 | 0.00 |
| TCGA-AF-6136 | Proficient | 72 | T3 | Rectosigmoid Junction | FEMALE | Rectal Adenocarcinoma | C19 | 0.02 |
| TCGA-AF-6655 | Proficient | 66 | T2 | Rectum | MALE | Rectal Adenocarcinoma | C19 | 0.02 |
| TCGA-AF-6672 | Proficient | 43 | T4a | Rectosigmoid Junction | MALE | Rectal Adenocarcinoma | C19 | 0.02 |
| TCGA-AF-A56K | Proficient | 56 | T3 | Sigmoid Colon | MALE | Rectal Adenocarcinoma | C19 | 0.09 |
| TCGA-AF-A56L | Proficient | 48 | T3 | Sigmoid Colon | FEMALE | Rectal Adenocarcinoma | C19 | 0.04 |
| TCGA-AF-A56N | Proficient | 47 | T3 | Rectum | FEMALE | Rectal Adenocarcinoma | C19 | 0.02 |
| TCGA-AG-3591 | Proficient | 66 | T3 | Rectum | FEMALE | Rectal Adenocarcinoma | C19 | 0.11 |
| TCGA-AG-3592 | Proficient | 68 | T3 | Rectum | MALE | Rectal Adenocarcinoma | C19 | 0.09 |
| TCGA-AG-3725 | Proficient | 90 | T3 | Rectum | FEMALE | Rectal Adenocarcinoma | C19 | 0.00 |
| TCGA-AG-3726 | Proficient | 63 | T2 | Rectum | FEMALE | Rectal Adenocarcinoma | C20 | 0.09 |
| TCGA-AG-3727 | Proficient | 78 | T3 | Rectum | FEMALE | Rectal Adenocarcinoma | C19 | 0.09 |
| TCGA-AG-3728 | Proficient | 73 | T3 | Rectum | MALE | Rectal Adenocarcinoma | C20 | 0.00 |
| TCGA-AG-3731 | Proficient | 65 | T3 | Rectum | MALE | Rectal Adenocarcinoma | C20 | 0.04 |
| TCGA-AG-3732 | Proficient | 78 | T2 | Rectum | FEMALE | Rectal Adenocarcinoma | C19 | 0.02 |
| TCGA-AG-3742 | Proficient | 85 | T1 | Rectum | FEMALE | Rectal Adenocarcinoma | C18.9 | 0.04 |
| TCGA-AG-3878 | Proficient | 64 | T2 | Rectum | MALE | Rectal Adenocarcinoma | C20 | 0.09 |
| TCGA-AG-3881 | Proficient | 83 | T3 | Rectum | FEMALE | [Not Available] | C80.1 | 0.00 |
| TCGA-AG-3882 | Proficient | 66 | T2 | Rectum | FEMALE | Rectal Adenocarcinoma | C18.9 | 0.04 |
| TCGA-AG-3883 | Proficient | 69 | T2 | Rectum | MALE | Rectal Adenocarcinoma | C20 | 0.02 |
| TCGA-AG-3885 | Proficient | 71 | T3 | Rectum | FEMALE | Rectal Adenocarcinoma | C20 | 0.00 |
| TCGA-AG-3887 | Proficient | 68 | T3 | Rectum | MALE | Rectal Mucinous Adenocarcinoma | C20 | 0.04 |
| TCGA-AG-3890 | Proficient | 62 | T2 | Rectum | MALE | Rectal Adenocarcinoma | C20 | 0.04 |
| TCGA-AG-3893 | Proficient | 74 | T3 | Rectum | MALE | Rectal Adenocarcinoma | C19 | 0.02 |
| TCGA-AG-3894 | Proficient | 65 | T3 | Rectum | MALE | Rectal Adenocarcinoma | C20 | 0.02 |

| | | | | | | | | |
|---|---|---|---|---|---|---|---|---|
| TCGA-AG-3896 | Proficient | 85 | T2 | Rectum | FEMALE | Rectal Adenocarcinoma | C20 | 0.00 |
| TCGA-AG-3898 | Proficient | 61 | T3 | Rectum | MALE | Rectal Adenocarcinoma | C20 | 0.07 |
| TCGA-AG-3901 | Proficient | 67 | T3 | Rectum | FEMALE | Rectal Mucinous Adenocarcinoma | C19 | 0.00 |
| TCGA-AG-3902 | Proficient | 61 | T3 | Rectum | MALE | Rectal Adenocarcinoma | C19 | 0.07 |
| TCGA-AG-3909 | Proficient | 69 | T3 | Rectum | FEMALE | Rectal Adenocarcinoma | C19 | 0.04 |
| TCGA-AG-4001 | Proficient | 74 | T3 | Rectum | FEMALE | Rectal Adenocarcinoma | C19 | 0.02 |
| TCGA-AG-4008 | Proficient | 63 | T3 | Rectum | MALE | Rectal Adenocarcinoma | C18.9 | 0.02 |
| TCGA-AG-4009 | Proficient | 83 | T2 | Rectum | MALE | Rectal Adenocarcinoma | C20 | 0.07 |
| TCGA-AG-4015 | Proficient | 85 | T3 | Rectum | FEMALE | Rectal Adenocarcinoma | C18.9 | 0.02 |
| TCGA-AG-4021 | Proficient | 84 | T3 | Rectum | FEMALE | Rectal Adenocarcinoma | C20 | 0.04 |
| TCGA-AG-4022 | Proficient | 59 | T3 | Rectum | FEMALE | Rectal Adenocarcinoma | C20 | 0.02 |
| TCGA-AG-A008 | Proficient | 50 | T2 | Rectum | FEMALE | Rectal Mucinous Adenocarcinoma | C20 | 0.02 |
| TCGA-AG-A00C | Proficient | 49 | T3 | Rectum | FEMALE | Rectal Adenocarcinoma | C20 | 0.07 |
| TCGA-AG-A00Y | Proficient | 68 | T3 | Rectum | MALE | Rectal Adenocarcinoma | C20 | 0.04 |
| TCGA-AG-A011 | Proficient | 80 | T3 | Rectum | MALE | Rectal Adenocarcinoma | C20 | 0.04 |
| TCGA-AG-A014 | Proficient | 86 | T2 | Rectum | MALE | Rectal Adenocarcinoma | C20 | 0.04 |
| TCGA-AG-A015 | Proficient | 64 | T1 | Rectum | FEMALE | Rectal Adenocarcinoma | C20 | 0.02 |
| TCGA-AG-A016 | Proficient | 55 | T3 | Rectum | MALE | Rectal Adenocarcinoma | C20 | 0.00 |
| TCGA-AG-A01J | Proficient | 59 | T3 | Rectum | FEMALE | Rectal Adenocarcinoma | C20 | 0.02 |
| TCGA-AG-A01L | Proficient | 58 | T3 | Rectum | MALE | Rectal Adenocarcinoma | C20 | 0.00 |
| TCGA-AG-A01N | Proficient | 68 | T2 | Rectum | FEMALE | Rectal Adenocarcinoma | C20 | 0.04 |
| TCGA-AG-A01W | Proficient | 67 | T3 | Rectum | FEMALE | Rectal Adenocarcinoma | C20 | 0.00 |
| TCGA-AG-A01Y | Proficient | 49 | T3 | Rectum | FEMALE | Rectal Adenocarcinoma | C20 | 0.04 |
| TCGA-AG-A020 | Proficient | 57 | T3 | Rectum | FEMALE | Rectal Mucinous Adenocarcinoma | C20 | 0.02 |
| TCGA-AG-A023 | Proficient | 62 | T4 | Rectum | FEMALE | Rectal Adenocarcinoma | C20 | 0.04 |
| TCGA-AG-A026 | Proficient | 66 | T4 | Rectum | MALE | Rectal Adenocarcinoma | C20 | 0.04 |
| TCGA-AG-A02X | Proficient | 77 | T2 | Rectum | MALE | Rectal Adenocarcinoma | C20 | 0.07 |
| TCGA-AG-A032 | Proficient | 68 | T3 | Rectum | MALE | Rectal Adenocarcinoma | C20 | 0.00 |
| TCGA-AG-A036 | Proficient | 71 | T3 | Rectum | MALE | Rectal Adenocarcinoma | C20 | 0.04 |
| TCGA-AH-6544 | Proficient | 60 | T3 | Rectosigmoid Junction | MALE | Rectal Adenocarcinoma | C19 | 0.04 |
| TCGA-AH-6547 | Proficient | 79 | T3 | Rectosigmoid Junction | FEMALE | Rectal Adenocarcinoma | C19 | 0.00 |
| TCGA-AH-6549 | Proficient | 66 | T3 | Rectum | MALE | Rectal Adenocarcinoma | C20 | 0.02 |
| TCGA-AH-6643 | Proficient | 50 | T3 | Rectosigmoid Junction | MALE | Rectal Adenocarcinoma | C19 | 0.04 |
| TCGA-AH-6644 | Proficient | 73 | T3 | Rectosigmoid Junction | MALE | Rectal Adenocarcinoma | C19 | 0.07 |
| TCGA-AH-6897 | Proficient | 48 | T2 | Rectosigmoid Junction | MALE | Rectal Adenocarcinoma | C19 | 0.04 |
| TCGA-AH-6903 | Proficient | 46 | T3 | Rectosigmoid Junction | MALE | Rectal Mucinous Adenocarcinoma | C19 | 0.00 |

| | | | | | | | | |
|---|---|---|---|---|---|---|---|---|
| TCGA-BM-6198 | Proficient | 73 | T3 | Rectum | MALE | Rectal Adenocarcinoma | C20 | 0.02 |
| TCGA-CI-6619 | Proficient | 41 | T3 | Rectum | MALE | Rectal Adenocarcinoma | C20 | 0.11 |
| TCGA-CI-6620 | Proficient | 41 | T3 | Rectum | FEMALE | Rectal Adenocarcinoma | C20 | 0.11 |
| TCGA-CI-6621 | Proficient | 63 | T3 | Rectum | MALE | Rectal Adenocarcinoma | C20 | 0.00 |
| TCGA-CI-6622 | Proficient | 74 | T4 | Rectum | MALE | Rectal Adenocarcinoma | C20 | 0.02 |
| TCGA-CI-6623 | Proficient | 44 | T1 | Rectum | MALE | Rectal Adenocarcinoma | C20 | 0.09 |
| TCGA-CI-6624 | Proficient | 53 | T2 | Rectosigmoid Junction | FEMALE | Rectal Adenocarcinoma | C20 | 0.07 |
| TCGA-CL-4957 | Proficient | 79 | T3 | [Not Available] | FEMALE | Rectal Adenocarcinoma | C20 | 0.02 |
| TCGA-CL-5917 | Proficient | 71 | T3 | Rectosigmoid Junction | FEMALE | Rectal Adenocarcinoma | C19 | 0.02 |
| TCGA-CL-5918 | Proficient | 90 | T3 | Rectosigmoid Junction | FEMALE | Rectal Adenocarcinoma | C20 | 0.07 |
| TCGA-DC-4745 | Proficient | 49 | T3 | Rectosigmoid Junction | FEMALE | Rectal Adenocarcinoma | C19 | 0.02 |
| TCGA-DC-4749 | Proficient | 57 | T2 | Rectosigmoid Junction | MALE | Rectal Adenocarcinoma | C19 | 0.00 |
| TCGA-DC-5337 | Proficient | 69 | T1 | Rectosigmoid Junction | MALE | Rectal Adenocarcinoma | C19 | 0.02 |
| TCGA-DC-5869 | Proficient | 62 | T3 | Rectum | FEMALE | Rectal Adenocarcinoma | C20 | 0.02 |
| TCGA-DC-6154 | Proficient | 57 | T4a | Rectosigmoid Junction | FEMALE | Rectal Adenocarcinoma | C19 | 0.00 |
| TCGA-DC-6155 | Proficient | 31 | T2 | Rectosigmoid Junction | FEMALE | Rectal Adenocarcinoma | C19 | 0.00 |
| TCGA-DC-6157 | Proficient | 48 | T2 | Rectosigmoid Junction | MALE | Rectal Adenocarcinoma | C19 | 0.04 |
| TCGA-DC-6158 | Proficient | 70 | T2 | Rectum | MALE | Rectal Adenocarcinoma | C20 | 0.00 |
| TCGA-DC-6160 | Proficient | 68 | T2 | Rectosigmoid Junction | MALE | Rectal Adenocarcinoma | C19 | 0.13 |
| TCGA-DC-6681 | Proficient | 70 | T3 | Rectosigmoid Junction | FEMALE | Rectal Adenocarcinoma | C19 | 0.02 |
| TCGA-DC-6682 | Proficient | 57 | T3 | Rectosigmoid Junction | MALE | Rectal Adenocarcinoma | C19 | 0.13 |
| TCGA-DC-6683 | Proficient | 43 | T3 | Rectosigmoid Junction | MALE | Rectal Adenocarcinoma | C19 | 0.00 |
| TCGA-DT-5265 | Proficient | 51 | T3 | Rectum | MALE | Rectal Mucinous Adenocarcinoma | C20 | 0.02 |
| TCGA-DY-A0XA | Proficient | 57 | T3 | Rectosigmoid Junction | FEMALE | Rectal Adenocarcinoma | C19 | 0.09 |
| TCGA-DY-A1DC | Proficient | 72 | T3 | Rectosigmoid Junction | FEMALE | Rectal Adenocarcinoma | C19 | 0.04 |
| TCGA-DY-A1DD | Proficient | 77 | T3 | Rectosigmoid Junction | FEMALE | Rectal Adenocarcinoma | C19 | 0.04 |
| TCGA-DY-A1DF | Proficient | 73 | T3 | Rectosigmoid Junction | FEMALE | Rectal Adenocarcinoma | C19 | 0.04 |
| TCGA-DY-A1DG | Proficient | 75 | T3 | Rectum | MALE | Rectal Adenocarcinoma | C20 | 0.02 |
| TCGA-DY-A1H8 | Proficient | 77 | T2 | Rectum | FEMALE | Rectal Adenocarcinoma | C20 | 0.04 |
| TCGA-EF-5830 | Proficient | 54 | T4a | Rectum | MALE | Rectal Adenocarcinoma | C20 | 0.04 |
| TCGA-EF-5831 | Proficient | 72 | T3 | Rectum | MALE | Rectal Adenocarcinoma | C20 | 0.04 |
| TCGA-EI-6506 | Proficient | 78 | T3 | Rectosigmoid Junction | FEMALE | Rectal Adenocarcinoma | C20 | 0.04 |
| TCGA-EI-6508 | Proficient | 48 | T3 | Rectosigmoid Junction | FEMALE | Rectal Adenocarcinoma | C20 | 0.07 |
| TCGA-EI-6509 | Proficient | 53 | T3 | Rectosigmoid Junction | MALE | Rectal Adenocarcinoma | C20 | 0.02 |
| TCGA-EI-6510 | Proficient | 77 | T2 | Rectosigmoid Junction | FEMALE | Rectal Adenocarcinoma | C20 | 0.02 |
| TCGA-EI-6511 | Proficient | 52 | T3 | Rectum | MALE | Rectal Adenocarcinoma | C20 | 0.07 |

| | | | | | | | | |
|---|---|---|---|---|---|---|---|---|
| TCGA-EI-6512 | Proficient | 64 | T3 | Rectosigmoid Junction | FEMALE | Rectal Adenocarcinoma | C20 | 0.11 |
| TCGA-EI-6513 | Proficient | 59 | T3 | Rectosigmoid Junction | MALE | Rectal Adenocarcinoma | C20 | 0.07 |
| TCGA-EI-6514 | Proficient | 59 | T3 | Sigmoid Colon | FEMALE | Rectal Adenocarcinoma | C20 | 0.02 |
| TCGA-EI-6881 | Proficient | 60 | T3 | Rectosigmoid Junction | MALE | Rectal Adenocarcinoma | C20 | 0.02 |
| TCGA-EI-6883 | Proficient | 63 | T3 | Rectosigmoid Junction | MALE | Rectal Adenocarcinoma | C20 | 0.11 |
| TCGA-EI-6884 | Proficient | 71 | T3 | Rectosigmoid Junction | MALE | Rectal Adenocarcinoma | C20 | 0.07 |
| TCGA-EI-6885 | Proficient | 57 | T3 | Rectosigmoid Junction | FEMALE | Rectal Adenocarcinoma | C20 | 0.04 |
| TCGA-EI-7002 | Proficient | 58 | T3 | Rectum | MALE | Rectal Adenocarcinoma | C20 | 0.00 |
| TCGA-EI-7004 | Proficient | 37 | T4a | Rectosigmoid Junction | FEMALE | Rectal Mucinous Adenocarcinoma | C19 | 0.04 |
| TCGA-F5-6464 | Proficient | 77 | T4b | Rectum | FEMALE | Rectal Adenocarcinoma | C19 | 0.02 |
| TCGA-F5-6465 | Proficient | 64 | T3 | Rectosigmoid Junction | FEMALE | Rectal Adenocarcinoma | C19 | 0.00 |
| TCGA-F5-6571 | Proficient | 62 | T3 | Rectum | FEMALE | Rectal Adenocarcinoma | C20 | 0.04 |
| TCGA-F5-6702 | Proficient | 71 | T3 | Rectosigmoid Junction | MALE | Rectal Adenocarcinoma | C19 | 0.00 |
| TCGA-F5-6810 | Proficient | 71 | NA | NA | MALE | Rectal Adenocarcinoma | NA | 0.02 |
| TCGA-F5-6811 | Proficient | 72 | T3 | Rectosigmoid Junction | FEMALE | Rectal Adenocarcinoma | C19 | 0.04 |
| TCGA-F5-6812 | Proficient | 67 | T3 | Rectum | MALE | Rectal Adenocarcinoma | C49.4 | 0.04 |
| TCGA-F5-6813 | Proficient | 70 | T4a | Rectum | MALE | Rectal Adenocarcinoma | C49.4 | 0.02 |
| TCGA-F5-6861 | Proficient | 60 | T3 | Rectum | FEMALE | Rectal Adenocarcinoma | C20 | 0.00 |
| TCGA-F5-6863 | Proficient | 71 | T4a | Rectum | FEMALE | Rectal Adenocarcinoma | C20 | 0.02 |
| TCGA-F5-6864 | Proficient | 74 | T3 | Rectum | FEMALE | Rectal Adenocarcinoma | C20 | 0.11 |
| TCGA-G5-6233 | Proficient | 74 | T3 | Sigmoid Colon | MALE | Rectal Adenocarcinoma | C20 | 0.07 |
| TCGA-G5-6235 | Proficient | 72 | T3 | Rectosigmoid Junction | MALE | Rectal Adenocarcinoma | C19 | 0.04 |
| TCGA-G5-6572 | Proficient | 56 | [Not Available] | Rectosigmoid Junction | MALE | Rectal Adenocarcinoma | C19 | 0.04 |
| TCGA-G5-6641 | Proficient | 67 | T1 | Rectosigmoid Junction | MALE | Rectal Mucinous Adenocarcinoma | C19 | 0.11 |
| TCGA-A6-2672 | Deficient | 82 | T3 | NA | FEMALE | Colon Adenocarcinoma | C18.2 | 4.01 |
| TCGA-A6-2686 | Deficient | 81 | T3 | Cecum | FEMALE | Colon Adenocarcinoma | C18.9 | 11.77 |
| TCGA-A6-3809 | Deficient | 71 | T4 | Transverse Colon | FEMALE | Colon Mucinous Adenocarcinoma | C18.9 | 8.20 |
| TCGA-A6-5661 | Deficient | 80 | T3 | Ascending Colon | FEMALE | Colon Adenocarcinoma | C18.2 | 6.70 |
| TCGA-A6-5665 | Deficient | 84 | T3 | Ascending Colon | FEMALE | Colon Adenocarcinoma | C18.2 | 11.35 |
| TCGA-A6-6653 | Deficient | 82 | T2 | Ascending Colon | MALE | Colon Adenocarcinoma | C18.9 | 6.72 |
| TCGA-AA-3492 | Deficient | 90 | T3 | Ascending Colon | FEMALE | Colon Adenocarcinoma | C18.2 | 10.31 |
| TCGA-AA-3663 | Deficient | 42 | T3 | Cecum | MALE | Colon Adenocarcinoma | C18.0 | 11.73 |
| TCGA-AA-3672 | Deficient | 90 | T3 | Transverse Colon | FEMALE | Colon Adenocarcinoma | C18.4 | 11.91 |
| TCGA-AA-3713 | Deficient | 68 | T3 | Ascending Colon | MALE | Colon Adenocarcinoma | C18.9 | 7.21 |
| TCGA-AA-3715 | Deficient | 77 | T3 | Ascending Colon | MALE | Colon Adenocarcinoma | C18.2 | 11.86 |

| | | | | | | | | |
|---|---|---|---|---|---|---|---|---|
| TCGA-AA-3811 | Deficient | 84 | T3 | Hepatic Flexure | FEMALE | Colon Adenocarcinoma | C18.9 | 5.88 |
| TCGA-AA-3815 | Deficient | 65 | T3 | Cecum | FEMALE | Colon Adenocarcinoma | C18.0 | 4.46 |
| TCGA-AA-3821 | Deficient | 81 | T2 | Hepatic Flexure | FEMALE | Colon Mucinous Adenocarcinoma | C18.2 | 4.57 |
| TCGA-AA-3833 | Deficient | 63 | T3 | Ascending Colon | FEMALE | Colon Adenocarcinoma | C18.9 | 3.15 |
| TCGA-AA-3845 | Deficient | 86 | T3 | Ascending Colon | FEMALE | Colon Adenocarcinoma | C18.2 | 4.35 |
| TCGA-AA-3877 | Deficient | 83 | T1 | Transverse Colon | FEMALE | Colon Mucinous Adenocarcinoma | C18.4 | 5.32 |
| TCGA-AA-3947 | Deficient | 60 | T4 | Ascending Colon | FEMALE | Colon Mucinous Adenocarcinoma | C18.9 | 11.64 |
| TCGA-AA-3949 | Deficient | 87 | T3 | Ascending Colon | FEMALE | Colon Mucinous Adenocarcinoma | C18.2 | 4.57 |
| TCGA-AA-3950 | Deficient | 79 | T3 | Ascending Colon | FEMALE | Colon Mucinous Adenocarcinoma | C18.2 | 4.97 |
| TCGA-AA-3966 | Deficient | 89 | T3 | Hepatic Flexure | FEMALE | Colon Mucinous Adenocarcinoma | C18.9 | 5.65 |
| TCGA-AA-A01P | Deficient | 80 | T3 | Ascending Colon | FEMALE | Colon Adenocarcinoma | C18.2 | 4.66 |
| TCGA-AA-A022 | Deficient | 88 | T4 | Cecum | FEMALE | Colon Adenocarcinoma | C18.0 | 8.91 |
| TCGA-AA-A02R | Deficient | 84 | T3 | Cecum | FEMALE | Colon Adenocarcinoma | C18.0 | 8.25 |
| TCGA-AD-5900 | Deficient | 67 | T2 | Ascending Colon | MALE | Colon Mucinous Adenocarcinoma | C18.2 | 8.38 |
| TCGA-AD-6889 | Deficient | 76 | T3 | Ascending Colon | MALE | Colon Adenocarcinoma | C18.2 | 11.84 |
| TCGA-AD-6895 | Deficient | 84 | T3 | Cecum | MALE | Colon Adenocarcinoma | C18.0 | 5.88 |
| TCGA-AD-A5EJ | Deficient | 74 | T3 | Cecum | FEMALE | Colon Adenocarcinoma | C18.0 | 8.78 |
| TCGA-AM-5821 | Deficient | 68 | T3 | Sigmoid Colon | FEMALE | Colon Adenocarcinoma | C18.7 | 5.34 |
| TCGA-AU-6004 | Deficient | 69 | T2 | Cecum | FEMALE | Colon Adenocarcinoma | C18.0 | 6.87 |
| TCGA-AY-6197 | Deficient | 60 | T3 | Cecum | MALE | Colon Adenocarcinoma | C18.2 | 6.39 |
| TCGA-AZ-4615 | Deficient | 84 | T3 | [Not Available] | MALE | Colon Adenocarcinoma | C18.0 | 5.88 |
| TCGA-AZ-6598 | Deficient | 77 | T3 | NA | FEMALE | Colon Adenocarcinoma | C18.2 | 16.34 |
| TCGA-CK-4951 | Deficient | 79 | T3 | Ascending Colon | FEMALE | Colon Mucinous Adenocarcinoma | C18.0 | 6.32 |
| TCGA-CK-5913 | Deficient | 58 | T3 | Cecum | FEMALE | Colon Adenocarcinoma | C18.2 | 5.50 |
| TCGA-CK-5916 | Deficient | 71 | T1 | Cecum | FEMALE | Colon Adenocarcinoma | C18.2 | 9.38 |
| TCGA-CK-6746 | Deficient | 84 | T4b | Cecum | FEMALE | Colon Adenocarcinoma | C18.0 | 8.18 |
| TCGA-CM-4743 | Deficient | 69 | T3 | Hepatic Flexure | MALE | Colon Adenocarcinoma | C18.2 | 6.96 |
| TCGA-CM-5861 | Deficient | 63 | T3 | Cecum | FEMALE | Colon Adenocarcinoma | C18.0 | 7.45 |
| TCGA-CM-6171 | Deficient | 77 | T2 | Ascending Colon | FEMALE | Colon Adenocarcinoma | C18.2 | 7.16 |
| TCGA-D5-6540 | Deficient | 66 | T2 | Cecum | MALE | Colon Mucinous Adenocarcinoma | C18.0 | 8.12 |
| TCGA-D5-6928 | Deficient | 80 | T3 | Ascending Colon | MALE | Colon Mucinous Adenocarcinoma | C18.3 | 7.10 |
| TCGA-D5-6930 | Deficient | 67 | T3 | Ascending Colon | MALE | Colon Mucinous Adenocarcinoma | C18.0 | 6.59 |
| TCGA-DM-A1HB | Deficient | 75 | T3 | Transverse Colon | MALE | NA | C18.4 | 6.30 |
| TCGA-F4-6570 | Deficient | 78 | T3 | Transverse Colon | FEMALE | Colon Adenocarcinoma | C18.9 | 8.29 |
| TCGA-G4-6302 | Deficient | 90 | T3 | Cecum | FEMALE | Colon Mucinous Adenocarcinoma | C18.0 | 5.52 |
| TCGA-G4-6304 | Deficient | 66 | T4 | Transverse Colon | FEMALE | Colon Adenocarcinoma | C18.4 | 4.99 |

| | | | | | | | | |
|---|---|---|---|---|---|---|---|---|
| TCGA-G4-6586 | Deficient | 73 | T3 | Ascending Colon | FEMALE | Colon Adenocarcinoma | C18.2 | 6.74 |
| TCGA-G4-6588 | Deficient | 58 | T3 | Cecum | FEMALE | Colon Adenocarcinoma | C18.0 | 11.11 |
| TCGA-G4-6628 | Deficient | 78 | T2 | Cecum | MALE | Colon Adenocarcinoma | C18.0 | 9.51 |
| TCGA-QG-A5Z2 | Deficient | 61 | T2 | Cecum | MALE | Colon Adenocarcinoma | C18.0 | 5.46 |
| TCGA-WS-AB45 | Deficient | 52 | T3 | Cecum | FEMALE | Colon Mucinous Adenocarcinoma | C18.0 | 13.17 |

**Supplementary Table 3.** Summary of the hereditary CRC and polyposis syndromes investigated in this study, including their underlying gene defect, previously reported mutational signature associations and the number of individuals and CRCs tested by WES for each group. In addition to CRCs from the ACCFR, OCCFR, and WEHI studies, non-hereditary CRCs from the TCGA COAD and READ studies were included as a separate group of controls for validation.

| Hereditary | Defective gene/s | DNA repair mechanism | Associated Signatures | Study Group | Individuals | CRCs | CRC Study IDs |
|---|---|---|---|---|---|---|---|
| **Hereditary CRCs** | | | | | | | |
| MUTYH-associated polyposis (MAP) | Biallelic *MUTYH* | Base excision repair | SBS18, SBS36 | Biallelic *MUTYH* carrier | 8 | 12 | M01-M12 |
| | | | | Monoallelic *MUTYH* carrier | 8 | 8 | W01-W08 |
| | | | | *MUTYH* VUS carrier | 1 | 1 | W09 |
| MMR PV Carriers (Lynch syndrome) | *MLH1, MSH2, MSH6, PMS2* | Mismatch repair | SBS6, SBS14, SBS15, SBS20, SBS21, SBS26, SBS44, ID2, ID7 | *MLH1* carriers | 14 | 14 | L30-L43 |
| | | | | *MSH2* carriers | 12 | 12 | L01-L12 |
| | | | | *MSH6* carriers | 7 | 7 | L20-L26 |
| **Non-hereditary CRCs** | | | | | | | |
| Sporadic MMR-deficient CRC | *MLH1* tumour methylation | Mismatch repair | | MMRd controls | 25 | 25 | K01-K25 |
| Sporadic MMR-proficient CRC | | | | MMRp controls | 160 | 160 | C01-C162 |
| **TOTAL** | | | | | **235** | **239** | |
| **TCGA Non-hereditary CRCs** | | | | | | | |
| Sporadic MMR-deficient CRC | *MLH1* tumour methylation | Mismatch repair | | TCGA MMRd controls | 52 | 52 | |
| Sporadic MMR-proficient CRC | | | | TCGA MMRp controls | 446 | 446 | |
| **TOTAL TCGA** | | | | | **498** | **498** | |

**Supplementary Table 4**. The accuracy (acc), sensitivity (sens), specificity (spec), positive predictive value (PPV), and negative predictive value (NPV) for each comparison in the study, across the discovery, validation and combined datasets, for a range of possible TMS thresholds.

| **MUTYH Biallelic v MMRp all** | | | | | | **Discovery** | | | | | **Validation** | | | | |
|---|---|---|---|---|---|---|---|---|---|---|---|---|---|---|---|
| **Threshold SBS18+SBS36** | **Acc** | **Sens** | **Spec** | **PPV** | **NPV** | **Acc** | **Sens** | **Spec** | **PPV** | **NPV** | **Acc** | **Sens** | **Spec** | **PPV** | **NPV** |
| 5% | 82.6% | 100.0% | 81.2% | 28.6% | 100.0% | 83.0% | 100.0% | 81.6% | 30.8% | 100.0% | 81.8% | 100.0% | 80.6% | 25.0% | 100.0% |
| 10% | 94.8% | 100.0% | 94.4% | 57.1% | 100.0% | 93.4% | 100.0% | 92.9% | 53.3% | 100.0% | 97.0% | 100.0% | 96.8% | 66.7% | 100.0% |
| 15% | 97.1% | 100.0% | 96.9% | 70.6% | 100.0% | 96.2% | 100.0% | 95.9% | 66.7% | 100.0% | 98.5% | 100.0% | 98.4% | 80.0% | 100.0% |
| 20% | 98.8% | 100.0% | 98.8% | 85.7% | 100.0% | 98.1% | 100.0% | 98.0% | 80.0% | 100.0% | 100.0% | 100.0% | 100.0% | 100.0% | 100.0% |
| 25% | 99.4% | 100.0% | 99.4% | 92.3% | 100.0% | 99.1% | 100.0% | 99.0% | 88.9% | 100.0% | 100.0% | 100.0% | 100.0% | 100.0% | 100.0% |
| **30%** | **100.0%** | **100.0%** | **100.0%** | **100.0%** | **100.0%** | **100.0%** | **100.0%** | **100.0%** | **100.0%** | **100.0%** | **100.0%** | **100.0%** | **100.0%** | **100.0%** | **100.0%** |
| 35% | 98.8% | 83.3% | 100.0% | 100.0% | 98.8% | 99.1% | 87.5% | 100.0% | 100.0% | 99.0% | 98.5% | 75.0% | 100.0% | 100.0% | 98.4% |
| 40% | 98.8% | 83.3% | 100.0% | 100.0% | 98.8% | 99.1% | 87.5% | 100.0% | 100.0% | 99.0% | 98.5% | 75.0% | 100.0% | 100.0% | 98.4% |
| 45% | 98.8% | 83.3% | 100.0% | 100.0% | 98.8% | 99.1% | 87.5% | 100.0% | 100.0% | 99.0% | 98.5% | 75.0% | 100.0% | 100.0% | 98.4% |
| 50% | 97.7% | 66.7% | 100.0% | 100.0% | 97.6% | 98.1% | 75.0% | 100.0% | 100.0% | 98.0% | 97.0% | 50.0% | 100.0% | 100.0% | 96.9% |
| 55% | 97.7% | 66.7% | 100.0% | 100.0% | 97.6% | 98.1% | 75.0% | 100.0% | 100.0% | 98.0% | 97.0% | 50.0% | 100.0% | 100.0% | 96.9% |
| 60% | 97.1% | 58.3% | 100.0% | 100.0% | 97.0% | 97.2% | 62.5% | 100.0% | 100.0% | 97.0% | 97.0% | 50.0% | 100.0% | 100.0% | 96.9% |
| 65% | 95.3% | 33.3% | 100.0% | 100.0% | 95.2% | 94.3% | 25.0% | 100.0% | 100.0% | 94.2% | 97.0% | 50.0% | 100.0% | 100.0% | 96.9% |
| 70% | 93.6% | 8.3% | 100.0% | 100.0% | 93.6% | 92.5% | 0.0% | 100.0% | 0.0% | 92.5% | 95.5% | 25.0% | 100.0% | 100.0% | 95.4% |
| 75% | 93.0% | 0.0% | 100.0% | 0.0% | 93.0% | 92.5% | 0.0% | 100.0% | 0.0% | 92.5% | 93.9% | 0.0% | 100.0% | 0.0% | 93.9% |
| 80% | 93.0% | 0.0% | 100.0% | 0.0% | 93.0% | 92.5% | 0.0% | 100.0% | 0.0% | 92.5% | 93.9% | 0.0% | 100.0% | 0.0% | 93.9% |
| 85% | 93.0% | 0.0% | 100.0% | 0.0% | 93.0% | 92.5% | 0.0% | 100.0% | 0.0% | 92.5% | 93.9% | 0.0% | 100.0% | 0.0% | 93.9% |
| 90% | 93.0% | 0.0% | 100.0% | 0.0% | 93.0% | 92.5% | 0.0% | 100.0% | 0.0% | 92.5% | 93.9% | 0.0% | 100.0% | 0.0% | 93.9% |
| 95% | 93.0% | 0.0% | 100.0% | 0.0% | 93.0% | 92.5% | 0.0% | 100.0% | 0.0% | 92.5% | 93.9% | 0.0% | 100.0% | 0.0% | 93.9% |
| **Lynch v MMRp all** | | | | | | **Discovery** | | | | | **Validation** | | | | |
| **Threshold ID2+ID7** | **Acc** | **Sens** | **Spec** | **PPV** | **NPV** | **Acc** | **Sens** | **Spec** | **PPV** | **NPV** | **Acc** | **Sens** | **Spec** | **PPV** | **NPV** |
| 5% | 26.4% | 100.0% | 11.3% | 18.9% | 100.0% | 19.1% | 100.0% | 5.1% | 15.5% | 100.0% | 37.2% | 100.0% | 21.0% | 24.6% | 100.0% |
| 10% | 34.7% | 100.0% | 21.2% | 20.8% | 100.0% | 31.3% | 100.0% | 19.4% | 17.7% | 100.0% | 39.7% | 100.0% | 24.2% | 25.4% | 100.0% |
| 15% | 46.1% | 100.0% | 35.0% | 24.1% | 100.0% | 42.6% | 100.0% | 32.7% | 20.5% | 100.0% | 51.3% | 100.0% | 38.7% | 29.6% | 100.0% |
| 20% | 54.4% | 100.0% | 45.0% | 27.3% | 100.0% | 49.6% | 100.0% | 40.8% | 22.7% | 100.0% | 61.5% | 100.0% | 51.6% | 34.8% | 100.0% |
| 25% | 62.2% | 100.0% | 54.4% | 31.1% | 100.0% | 61.7% | 100.0% | 55.1% | 27.9% | 100.0% | 62.8% | 100.0% | 53.2% | 35.6% | 100.0% |
| 30% | 70.5% | 100.0% | 64.4% | 36.7% | 100.0% | 68.7% | 100.0% | 63.3% | 32.1% | 100.0% | 73.1% | 100.0% | 66.1% | 43.2% | 100.0% |

| | | | | | | | | | | | | | | | |
|---|---|---|---|---|---|---|---|---|---|---|---|---|---|---|---|
| 35% | 76.7% | 100.0% | 71.9% | 42.3% | 100.0% | 76.5% | 100.0% | 72.4% | 38.6% | 100.0% | 76.9% | 100.0% | 71.0% | 47.1% | 100.0% |
| 40% | 79.8% | 100.0% | 75.6% | 45.8% | 100.0% | 80.9% | 100.0% | 77.6% | 43.6% | 100.0% | 78.2% | 100.0% | 72.6% | 48.5% | 100.0% |
| 45% | 87.6% | 100.0% | 85.0% | 57.9% | 100.0% | 89.6% | 100.0% | 87.8% | 58.6% | 100.0% | 84.6% | 100.0% | 80.6% | 57.1% | 100.0% |
| 50% | 91.7% | 100.0% | 90.0% | 67.3% | 100.0% | 93.9% | 100.0% | 92.9% | 70.8% | 100.0% | 88.5% | 100.0% | 85.5% | 64.0% | 100.0% |
| 55% | 94.3% | 100.0% | 93.1% | 75.0% | 100.0% | 96.5% | 100.0% | 95.9% | 81.0% | 100.0% | 91.0% | 100.0% | 88.7% | 69.6% | 100.0% |
| 60% | 96.9% | 100.0% | 96.3% | 84.6% | 100.0% | 97.4% | 100.0% | 96.9% | 85.0% | 100.0% | 96.2% | 100.0% | 95.2% | 84.2% | 100.0% |
| 65% | 97.9% | 100.0% | 97.5% | 89.2% | 100.0% | 98.3% | 100.0% | 98.0% | 89.5% | 100.0% | 97.4% | 100.0% | 96.8% | 88.9% | 100.0% |
| 70% | 99.0% | 100.0% | 98.8% | 94.3% | 100.0% | 99.1% | 100.0% | 99.0% | 94.4% | 100.0% | 98.7% | 100.0% | 98.4% | 94.1% | 100.0% |
| **75%** | **99.5%** | **100.0%** | **99.4%** | **97.1%** | **100.0%** | **100.0%** | **100.0%** | **100.0%** | **100.0%** | **100.0%** | **98.7%** | **100.0%** | **98.4%** | **94.1%** | **100.0%** |
| 80% | 99.0% | 97.0% | 99.4% | 97.0% | 99.4% | 100.0% | 100.0% | 100.0% | 100.0% | 100.0% | 97.4% | 93.8% | 98.4% | 93.8% | 98.4% |
| 85% | 90.7% | 48.5% | 99.4% | 94.1% | 90.3% | 90.4% | 35.3% | 100.0% | 100.0% | 89.9% | 91.0% | 62.5% | 98.4% | 90.9% | 91.0% |
| 90% | 85.0% | 15.2% | 99.4% | 83.3% | 85.0% | 86.1% | 5.9% | 100.0% | 100.0% | 86.0% | 83.3% | 25.0% | 98.4% | 80.0% | 83.6% |
| 95% | 82.4% | 0.0% | 99.4% | 0.0% | 82.8% | 85.2% | 0.0% | 100.0% | 0.0% | 85.2% | 78.2% | 0.0% | 98.4% | 0.0% | 79.2% |
| | **MMRd v MMRp** | | | | | **Discovery** | | | | | **Validation** | | | | |
| **Threshold ID2+ID7** | **Acc** | **Sens** | **Spec** | **PPV** | **NPV** | **Acc** | **Sens** | **Spec** | **PPV** | **NPV** | **Acc** | **Sens** | **Spec** | **PPV** | **NPV** |
| 5% | 34.9% | 100.0% | 11.3% | 29.0% | 100.0% | 27.3% | 100.0% | 5.1% | 24.4% | 100.0% | 45.6% | 100.0% | 21.0% | 36.4% | 100.0% |
| 10% | 42.2% | 100.0% | 21.2% | 31.5% | 100.0% | 38.3% | 100.0% | 19.4% | 27.5% | 100.0% | 47.8% | 100.0% | 24.2% | 37.3% | 100.0% |
| 15% | 52.3% | 100.0% | 35.0% | 35.8% | 100.0% | 48.4% | 100.0% | 32.7% | 31.2% | 100.0% | 57.8% | 100.0% | 38.7% | 42.4% | 100.0% |
| 20% | 59.6% | 100.0% | 45.0% | 39.7% | 100.0% | 54.7% | 100.0% | 40.8% | 34.1% | 100.0% | 66.7% | 100.0% | 51.6% | 48.3% | 100.0% |
| 25% | 66.5% | 100.0% | 54.4% | 44.3% | 100.0% | 65.6% | 100.0% | 55.1% | 40.5% | 100.0% | 67.8% | 100.0% | 53.2% | 49.1% | 100.0% |
| 30% | 73.9% | 100.0% | 64.4% | 50.4% | 100.0% | 71.9% | 100.0% | 63.3% | 45.5% | 100.0% | 76.7% | 100.0% | 66.1% | 57.1% | 100.0% |
| 35% | 79.4% | 100.0% | 71.9% | 56.3% | 100.0% | 78.9% | 100.0% | 72.4% | 52.6% | 100.0% | 80.0% | 100.0% | 71.0% | 60.9% | 100.0% |
| 40% | 82.1% | 100.0% | 75.6% | 59.8% | 100.0% | 82.8% | 100.0% | 77.6% | 57.7% | 100.0% | 81.1% | 100.0% | 72.6% | 62.2% | 100.0% |
| 45% | 89.0% | 100.0% | 85.0% | 70.7% | 100.0% | 90.6% | 100.0% | 87.8% | 71.4% | 100.0% | 86.7% | 100.0% | 80.6% | 70.0% | 100.0% |
| 50% | 92.7% | 100.0% | 90.0% | 78.4% | 100.0% | 94.5% | 100.0% | 92.9% | 81.1% | 100.0% | 90.0% | 100.0% | 85.5% | 75.7% | 100.0% |
| 55% | 95.0% | 100.0% | 93.1% | 84.1% | 100.0% | 96.9% | 100.0% | 95.9% | 88.2% | 100.0% | 92.2% | 100.0% | 88.7% | 80.0% | 100.0% |
| 60% | 97.2% | 100.0% | 96.3% | 90.6% | 100.0% | 97.7% | 100.0% | 96.9% | 90.9% | 100.0% | 96.7% | 100.0% | 95.2% | 90.3% | 100.0% |
| 65% | 98.2% | 100.0% | 97.5% | 93.5% | 100.0% | 98.4% | 100.0% | 98.0% | 93.8% | 100.0% | 97.8% | 100.0% | 96.8% | 93.3% | 100.0% |
| 70% | 98.6% | 98.3% | 98.8% | 96.6% | 99.4% | 99.2% | 100.0% | 99.0% | 96.8% | 100.0% | 97.8% | 96.4% | 98.4% | 96.4% | 98.4% |
| **75%** | **99.1%** | **98.3%** | **99.4%** | **98.3%** | **99.4%** | **100.0%** | **100.0%** | **100.0%** | **100.0%** | **100.0%** | **97.8%** | **96.4%** | **98.4%** | **96.4%** | **98.4%** |
| 80% | 98.2% | 94.8% | 99.4% | 98.2% | 98.1% | 100.0% | 100.0% | 100.0% | 100.0% | 100.0% | 95.6% | 89.3% | 98.4% | 96.2% | 95.3% |
| 85% | 88.5% | 58.6% | 99.4% | 97.1% | 86.9% | 90.6% | 60.0% | 100.0% | 100.0% | 89.1% | 85.6% | 57.1% | 98.4% | 94.1% | 83.6% |
| 90% | 79.8% | 25.9% | 99.4% | 93.8% | 78.7% | 85.2% | 36.7% | 100.0% | 100.0% | 83.8% | 72.2% | 14.3% | 98.4% | 80.0% | 71.8% |

| | | | | | | | | | | | | | | | |
|---|---|---|---|---|---|---|---|---|---|---|---|---|---|---|---|
| 95% | 73.9% | 3.4% | 99.4% | 66.7% | 74.0% | 78.1% | 6.7% | 100.0% | 100.0% | 77.8% | 67.8% | 0.0% | 98.4% | 0.0% | 68.5% |
| **Lynch v MMRd** | | | | | | **Discovery** | | | | | **Validation** | | | | |
| **Threshold SBS1** | **Acc** | **Sens** | **Spec** | **PPV** | **NPV** | **Acc** | **Sens** | **Spec** | **PPV** | **NPV** | **Acc** | **Sens** | **Spec** | **PPV** | **NPV** |
| 5% | 58.6% | 100.0% | 4.0% | 57.9% | 100.0% | 56.7% | 100.0% | 0.0% | 56.7% | 0.0% | 60.7% | 100.0% | 8.3% | 59.3% | 100.0% |
| 10% | 65.5% | 97.0% | 24.0% | 62.7% | 85.7% | 60.0% | 100.0% | 7.7% | 58.6% | 100.0% | 71.4% | 93.8% | 41.7% | 68.2% | 83.3% |
| 15% | 69.0% | 93.9% | 36.0% | 66.0% | 81.8% | 63.3% | 94.1% | 23.1% | 61.5% | 75.0% | 75.0% | 93.8% | 50.0% | 71.4% | 85.7% |
| **20%** | **74.1%** | **87.9%** | **56.0%** | **72.5%** | **77.8%** | **73.3%** | **88.2%** | **53.8%** | **71.4%** | **77.8%** | **75.0%** | **87.5%** | **58.3%** | **73.7%** | **77.8%** |
| 25% | 72.4% | 75.8% | 68.0% | 75.8% | 68.0% | 76.7% | 82.4% | 69.2% | 77.8% | 75.0% | 67.9% | 68.8% | 66.7% | 73.3% | 61.5% |
| 30% | 67.2% | 60.6% | 76.0% | 76.9% | 59.4% | 66.7% | 58.8% | 76.9% | 76.9% | 58.8% | 67.9% | 62.5% | 75.0% | 76.9% | 60.0% |
| 35% | 63.8% | 42.4% | 92.0% | 87.5% | 54.8% | 66.7% | 47.1% | 92.3% | 88.9% | 57.1% | 60.7% | 37.5% | 91.7% | 85.7% | 52.4% |
| 40% | 60.3% | 33.3% | 96.0% | 91.7% | 52.2% | 63.3% | 41.2% | 92.3% | 87.5% | 54.5% | 57.1% | 25.0% | 100.0% | 100.0% | 50.0% |
| 45% | 56.9% | 24.2% | 100.0% | 100.0% | 50.0% | 60.0% | 29.4% | 100.0% | 100.0% | 52.0% | 53.6% | 18.8% | 100.0% | 100.0% | 48.0% |
| 50% | 51.7% | 15.2% | 100.0% | 100.0% | 47.2% | 56.7% | 23.5% | 100.0% | 100.0% | 50.0% | 46.4% | 6.2% | 100.0% | 100.0% | 44.4% |
| 55% | 48.3% | 9.1% | 100.0% | 100.0% | 45.5% | 50.0% | 11.8% | 100.0% | 100.0% | 46.4% | 46.4% | 6.2% | 100.0% | 100.0% | 44.4% |
| 60% | 44.8% | 3.0% | 100.0% | 100.0% | 43.9% | 46.7% | 5.9% | 100.0% | 100.0% | 44.8% | 42.9% | 0.0% | 100.0% | 0.0% | 42.9% |
| 65% | 44.8% | 3.0% | 100.0% | 100.0% | 43.9% | 46.7% | 5.9% | 100.0% | 100.0% | 44.8% | 42.9% | 0.0% | 100.0% | 0.0% | 42.9% |
| 70% | 43.1% | 0.0% | 100.0% | 0.0% | 43.1% | 43.3% | 0.0% | 100.0% | 0.0% | 43.3% | 42.9% | 0.0% | 100.0% | 0.0% | 42.9% |
| 75% | 43.1% | 0.0% | 100.0% | 0.0% | 43.1% | 43.3% | 0.0% | 100.0% | 0.0% | 43.3% | 42.9% | 0.0% | 100.0% | 0.0% | 42.9% |
| 80% | 43.1% | 0.0% | 100.0% | 0.0% | 43.1% | 43.3% | 0.0% | 100.0% | 0.0% | 43.3% | 42.9% | 0.0% | 100.0% | 0.0% | 42.9% |
| 85% | 43.1% | 0.0% | 100.0% | 0.0% | 43.1% | 43.3% | 0.0% | 100.0% | 0.0% | 43.3% | 42.9% | 0.0% | 100.0% | 0.0% | 42.9% |
| 90% | 43.1% | 0.0% | 100.0% | 0.0% | 43.1% | 43.3% | 0.0% | 100.0% | 0.0% | 43.3% | 42.9% | 0.0% | 100.0% | 0.0% | 42.9% |
| 95% | 43.1% | 0.0% | 100.0% | 0.0% | 43.1% | 43.3% | 0.0% | 100.0% | 0.0% | 43.3% | 42.9% | 0.0% | 100.0% | 0.0% | 42.9% |

**Supplementary Table 5**. Each individual tumour mutational signature (TMS) was evaluated for its ability to identify the hereditary CRC groups, measured by the AUC, LD and mean difference. The number of CRCs included in each test are shown in the positive (Pos) and negative (Neg) columns. Each TMS found to have an unadjusted significant p-value (P) is included. The TMS that remained significant (adjusted p-value < 4.6 x $10^{-5}$) after Bonferroni correction (column AdjP) are in bold and are included in Table 1. Where LD is marked as n/a it could not be calculated due to all values from one group exhibiting the same TMS value.

| | All | | | | | | | Discovery | | | | | | | Validation | | | | | | |
|---|---|---|---|---|---|---|---|---|---|---|---|---|---|---|---|---|---|---|---|---|---|
| Classifier | AUC | LD | Mean Diff | Pos | Neg | P | AdjP | AUC | LD | Mean Diff | Pos | Neg | P | AdjP | AUC | LD | Mean Diff | Pos | Neg | P | AdjP |
| Biallelic MUTYH v non-MUTYH | | | | | | | | | | | | | | | | | | | | | |
| ID12 | 0.68 | 0.16 | 0.12 | 12 | 218 | 2E-03 | 1 | | | | | | | | 0.58 | 0.23 | 0.20 | 4 | 90 | 3E-03 | 1 |
| ID16 | 0.53 | 0.07 | 0.03 | 12 | 218 | 3E-02 | 1 | | | | | | | | 0.49 | 0.21 | 0.09 | 4 | 90 | 2E-03 | 1 |
| SBS14 | | | | | | | | | | | | | | | 0.67 | n/a | 0.01 | 4 | 90 | 7E-03 | 1 |
| **SBS18** | **0.98** | **1.93** | **0.26** | **12** | **218** | 1E-37 | **1E-34** | **0.96** | **1.11** | **0.22** | **8** | **128** | 5E-16 | **5.5E-13** | **1.00** | **12.34** | **0.34** | **4** | **90** | 3E-34 | **3E-31** |
| **SBS18,SBS36** | **1.00** | **16.17** | **0.55** | **12** | **218** | 4E-101 | **5E-98** | **1.00** | **20.81** | **0.56** | **8** | **128** | 4E-61 | **4.3E-58** | **1.00** | **10.99** | **0.53** | **4** | **90** | 7E-41 | **7E-38** |
| SBS24 | 0.72 | n/a | 0.01 | 12 | 218 | 2E-02 | 1 | | | | | | | | 0.78 | n/a | 0.02 | 4 | 90 | 2E-02 | 1 |
| **SBS36** | **0.96** | **2.35** | **0.29** | **12** | **218** | 2E-59 | **2E-56** | **0.94** | **2.99** | **0.34** | **8** | **128** | 8E-41 | **8.6E-38** | **1.00** | **2.28** | **0.19** | **4** | **90** | 8E-24 | **9E-21** |
| **SBS38** | **0.74** | **n/a** | **0.02** | **12** | **218** | 9E-18 | **1E-14** | **0.74** | **n/a** | **0.02** | **8** | **128** | 2E-12 | **1.7E-09** | **0.74** | **n/a** | **0.03** | **4** | **90** | 5E-08 | **6E-05** |
| MUTYH Monoallelic v Non-MUTYH | | | | | | | | | | | | | | | | | | | | | |
| ID11 | 0.62 | 0.18 | 0.04 | 9 | 218 | 0.02 | 1 | 0.58 | 0.15 | 0.04 | 6 | 128 | 4E-02 | 1 | | | | | | | |
| ID15 | | | | | | | | 0.64 | 0.33 | 0.06 | 6 | 128 | 7E-04 | 0.72 | | | | | | | |
| SBS12 | | | | | | | | | | | | | | | 0.77 | n/a | 0.01 | 3 | 90 | 2E-02 | 1 |
| SBS13 | | | | | | | | 0.76 | n/a | 0.01 | 6 | 128 | 2E-02 | 1 | | | | | | | |
| **SBS16** | **0.66** | **n/a** | **0.01** | **9** | **218** | 0.02 | **1** | | | | | | | | **0.88** | **0.87** | **0.04** | **3** | **90** | 1E-05 | **0.012** |
| SBS18 | 0.54 | n/a | 0.03 | 9 | 218 | 0.04 | 1 | 0.63 | 0.12 | 0.05 | 6 | 128 | 2E-02 | 1 | | | | | | | |
| SBS25 | | | | | | | | | | | | | | | 0.63 | n/a | 0.01 | 3 | 90 | 5E-02 | 1 |
| SBS29 | 0.63 | 0.33 | 0.03 | 9 | 218 | 2E-04 | 0.17 | 0.66 | 0.48 | 0.04 | 6 | 128 | 1E-04 | 0.16 | | | | | | | |
| SBS30 | | | | | | | | 0.70 | n/a | 0.02 | 6 | 128 | 5E-04 | 0.60 | | | | | | | |

| | | | | | | | | | | | | | | | | | | | | | |
|---|---|---|---|---|---|---|---|---|---|---|---|---|---|---|---|---|---|---|---|---|---|
| **SBS36** | 0.64 | n/a | 0.03 | 9 | 218 | 2.0E-09 | 2E-06 | 0.66 | 0.32 | 0.03 | 6 | 128 | 1E-08 | 2E-05 | | | | | | | |
| **SBS40** | 0.61 | n/a | 0.02 | 9 | 218 | 9.6E-08 | 1E-04 | 0.58 | n/a | 0.01 | 6 | 128 | 2E-02 | 1 | 0.67 | 0.50 | 0.04 | 3 | 90 | 1E-09 | 1E-06 |
| **SBS5** | 0.60 | n/a | 0.03 | 9 | 218 | 2E-03 | 1 | | | | | | | | 0.83 | 1.59 | 0.10 | 3 | 90 | 3E-16 | 3E-13 |
| SBS9 | 0.59 | n/a | 0.01 | 9 | 218 | 0.05 | 1 | | | | | | | | 0.81 | n/a | 0.02 | 3 | 90 | 1E-03 | 1 |
| **MMR PV Carrier v MMRp** | | | | | | | | | | | | | | | | | | | | | |
| **ID2** | 0.97 | 4.37 | 0.43 | 33 | 180 | 3.6E-31 | 4E-28 | 0.99 | 7.10 | 0.43 | 17 | 112 | 1E-21 | 1E-18 | 0.94 | 2.88 | 0.43 | 16 | 68 | 9E-12 | 9E-09 |
| **ID2, ID7** | 0.99 | 10.52 | 0.59 | 33 | 180 | 1.4E-47 | 2E-44 | 1.00 | 13.21 | 0.58 | 17 | 112 | 2E-30 | 3E-27 | 0.99 | 8.14 | 0.61 | 16 | 68 | 5E-19 | 6E-16 |
| **ID7** | 0.91 | 1.08 | 0.16 | 33 | 180 | 1.1E-17 | 1E-14 | 0.92 | 2.03 | 0.14 | 17 | 112 | 7E-11 | 8E-08 | 0.91 | 0.88 | 0.18 | 16 | 68 | 2E-08 | 2E-05 |
| **SBS1** | 0.67 | 0.20 | 0.08 | 33 | 180 | 3.2E-04 | 4E-01 | 0.69 | 0.27 | 0.10 | 17 | 112 | 1E-03 | 1 | 0.65 | 0.14 | 0.06 | 16 | 68 | 3E-02 | 1 |
| **SBS14** | 0.75 | n/a | 0.01 | 33 | 180 | 8.4E-06 | 9E-03 | 0.70 | n/a | 0.00 | 17 | 112 | 3E-03 | 1 | 0.79 | n/a | 0.01 | 16 | 68 | 1E-03 | 1 |
| **SBS15** | 0.84 | 0.73 | 0.13 | 33 | 180 | 3.8E-14 | 4E-11 | 0.86 | 1.13 | 0.11 | 17 | 112 | 5E-09 | 6E-06 | 0.80 | 0.62 | 0.15 | 16 | 68 | 1E-06 | 0.0015 |
| **SBS20** | 0.91 | 1.41 | 0.06 | 33 | 180 | 2.0E-35 | 2E-32 | 0.91 | 1.54 | 0.06 | 17 | 112 | 4E-23 | 5E-20 | 0.92 | 1.33 | 0.05 | 16 | 68 | 2E-14 | 2E-11 |
| **SBS21** | 0.73 | n/a | 0.01 | 33 | 180 | 9.4E-07 | 1E-03 | 0.73 | n/a | 0.01 | 17 | 112 | 1E-04 | 1E-01 | 0.73 | n/a | 0.01 | 16 | 68 | 1E-03 | 1 |
| **SBS26** | 0.59 | n/a | 0.01 | 33 | 180 | 1.9E-04 | 2E-01 | 0.60 | n/a | 0.01 | 17 | 112 | 2E-04 | 2E-01 | | | | | | | |
| **SBS6** | 0.80 | 0.66 | 0.10 | 33 | 180 | 9.3E-09 | 1E-05 | 0.80 | 0.66 | 0.09 | 17 | 112 | 2E-05 | 3E-02 | 0.78 | 0.66 | 0.10 | 16 | 68 | 7E-05 | 0.081 |
| **MMRd v MMRp** | | | | | | | | | | | | | | | | | | | | | |
| **ID2** | 0.97 | 4.12 | 0.42 | 58 | 180 | 2.6E-43 | 3E-40 | 0.97 | 5.16 | 0.40 | 30 | 112 | 2E-27 | 2E-24 | 0.95 | 3.32 | 0.44 | 28 | 68 | 2E-17 | 2E-14 |
| **ID2,ID7** | 0.99 | 10.50 | 0.60 | 58 | 180 | 3.1E-69 | 3E-66 | 1.00 | 13.85 | 0.61 | 30 | 112 | 4E-45 | 4E-42 | 0.99 | 7.63 | 0.60 | 28 | 68 | 2E-26 | 2E-23 |
| **ID7** | 0.93 | 1.44 | 0.19 | 58 | 180 | 6.0E-29 | 7E-26 | 0.95 | 2.27 | 0.21 | 30 | 112 | 3E-22 | 3E-19 | 0.92 | 0.99 | 0.16 | 28 | 68 | 3E-10 | 3E-07 |
| **SBS14** | 0.75 | n/a | 0.01 | 58 | 180 | 5.6E-09 | 6E-06 | 0.80 | n/a | 0.01 | 30 | 112 | 5E-09 | 5E-06 | 0.70 | n/a | 0.00 | 28 | 68 | 2E-02 | 1 |
| **SBS15** | 0.85 | 0.78 | 0.16 | 58 | 180 | 2.1E-20 | 2E-17 | 0.85 | 1.00 | 0.13 | 30 | 112 | 3E-13 | 3E-10 | 0.83 | 0.73 | 0.18 | 28 | 68 | 4E-09 | 5E-06 |
| **SBS20** | 0.93 | 1.80 | 0.07 | 58 | 180 | 8.4E-45 | 9E-42 | 0.95 | 2.58 | 0.08 | 30 | 112 | 3E-34 | 3E-31 | 0.90 | 1.33 | 0.05 | 28 | 68 | 3E-15 | 3E-12 |
| **SBS21** | 0.79 | n/a | 0.02 | 58 | 180 | 1.1E-13 | 1E-10 | 0.78 | n/a | 0.02 | 30 | 112 | 3E-09 | 4E-06 | 0.81 | n/a | 0.02 | 28 | 68 | 3E-06 | 3E-03 |
| **SBS26** | 0.60 | n/a | 0.01 | 58 | 180 | 2.0E-06 | 2E-03 | 0.60 | n/a | 0.01 | 30 | 112 | 7E-05 | 7E-02 | 0.60 | n/a | 0.01 | 28 | 68 | 5E-03 | 1 |
| **SBS6** | 0.74 | 0.40 | 0.08 | 58 | 180 | 3.4E-09 | 4E-06 | 0.77 | 0.48 | 0.09 | 30 | 112 | 1E-06 | 1E-03 | 0.72 | 0.33 | 0.07 | 28 | 68 | 3E-04 | 0.34 |
| **MMR PV Carrier v MLH1 Methylated** | | | | | | | | | | | | | | | | | | | | | |
| **ID12** | 0.66 | n/a | 0.02 | 33 | 25 | 0.02 | 1 | 0.95 | 3.51 | 0.07 | 17 | 13 | | 2E-04 | | | | | | | |
| ID2 | | | | | | | | 0.85 | 0.78 | 0.09 | 17 | 13 | | 1 | | | | | | | |

| | | | | | | | | | | | | | | | | | | | | | |
|---|---|---|---|---|---|---|---|---|---|---|---|---|---|---|---|---|---|---|---|---|---|
| **SBS1** | **0.80** | **0.76** | **0.15** | **33** | **25** | 1.9E-05 | **0.02** | **0.82** | **0.83** | **0.16** | **17** | **13** | | **1** | **0.79** | **0.73** | **0.14** | **16** | **12** | 3E-03 | **1** |
| SBS14 | | | | | | | | | | | | | | | 0.78 | n/a | 0.01 | 16 | 12 | 4E-03 | 1 |
| SBS37 | | | | | | | | 0.73 | n/a | 0.01 | 17 | 13 | | 1 | | | | | | | |
| SBS6 | | | | | | | | | | | | | | | 0.68 | 0.33 | 0.06 | 16 | 12 | 3E-02 | 1 |
| SBS7c | 0.68 | n/a | 0.01 | 33 | 25 | 5E-03 | 1 | 0.83 | n/a | 0.01 | 17 | 13 | | 0.78 | | | | | | | |

**Figures**

**Supplementary Figure 1.** The tumour mutational signature (TMS) profiles based on the COSMIC v3 signature set for each of the 97 CRCs tested by whole exome sequencing and included in the validation group: (a) Single base substitution (SBS)-derived TMS and (b) insertion and deletion (ID)-derived TMS profiles. CRCs were grouped by subtype: (i) biallelic *MUTYH* pathogenic variant carriers, (ii) monoallelic *MUTYH* pathogenic variant carriers, (iii) mismatch repair (MMR) gene pathogenic variant carriers (Lynch syndrome), (iv) MMR-deficient (MMRd) controls related to *MLH1* gene promoter hypermethylation and (v) MMR-proficient (MMRp) controls. Individual SBS or ID TMS with proportional values below 5% across all the CRC samples were excluded.

**Supplementary Figure 2.** Assessment of somatic loss of heterozygosity (LOH) across *MUTYH* in monoallelic samples W01 (monoallelic *MUTYH* germline pathogenic variant carrier), C20 (POLE somatic pathogenic variant) and L01 (*MSH2* germline pathogenic variant carrier) **(a-c)** as well as monoallelic *MUTYH* sample W07 which exhibited a combined SBS18 and SBS36 signature profile indicative of biallelic *MUTYH* inactivation **(d)**. Each dot indicates a variant seen at a given allele fraction in the tumour, with "+" indicating the equivalent germline allele fraction. LOH is unlikely in regions containing heterozygous variants (red), while somatic homozygous variants seen as heterozygous in the germline sequence are indicative of LOH (green). Samples W01, C20 and L01 did not exhibit LOH, nor any somatic pathogenic variant in *MUTYH*, while W07 exhibits LOH across the entire *MUTYH* gene. This suggests that LOH causes loss of the wildtype allele and accounts for the high SBS18 and SBS36 signature profile observe for this germline monoallelic *MUTYH* CRC.

10 Wellcome Sanger Institute. COSMIC Signatures of Mutational Processes in Human

Cancer. Signatures of Mutational Processes in Human Cancer. 2019.https://cancer.sanger.ac.uk/cosmic/signatures (accessed 31 May2019).

11 Huang MN, McPherson JR, Cutcutache I, *et al.* MSIseq: Software for Assessing Microsatellite Instability from Catalogs of Somatic Mutations. *Sci Rep* 2015;**5**:13321. doi:10.1038/srep13321

12 Webb AR. *Statistical Pattern Recognition*. 2nd ed. West Sussex, England: : Wiley 2002.

13 Bland JM, Altman DG. Multiple significance tests: the Bonferroni method. *BMJ* 1995;**310**:170.

14 Hanley JA, McNeil BJ. The meaning and use of the area under a receiver operating characteristic (ROC) curve. *Radiology* 1982;**143**:29–36. doi:10.1148/radiology.143.1.7063747

15 Obuchowski NA, Lieber ML. Confidence bounds when the estimated ROC area is 1.01. *Acad Radiol* 2002;**9**:526–30.

16 Marshall AW, Olkin I. Distributions with Bounded Support. In: *Life Distributions*. New York, NY: : Springer New York 2007. 473–95. doi:10.1007/978-0-387-68477-2_14

17 Johnson NL, Kotz S, Balakrishnan N. *Continuous Univariate Distributions*. John Wiley & Sons, Ltd 1995.

18 de Carvalho JF, Draper NR, Smith H. Applied Regression Analysis (2nd ed). *J Am Stat Assoc* 1981;**76**:1012. doi:10.2307/2287608

19 Tibshirani R. Regression shrinkage and selection via the lasso. *Journal of the Royal Statistical Society: Series B (Methodological)* 1996;**58**:267–88. doi:10.1111/j.2517-6161.1996.tb02080.x

20 Lynch HT, de la Chapelle A. Hereditary colorectal cancer. *N Engl J Med* 2003;**348**:919–32. doi:10.1056/NEJMra012242

21 Cancer Genome Atlas Network. Comprehensive molecular characterization of human colon and rectal cancer. *Nature* 2012;**487**:330–7. doi:10.1038/nature11252

22 Cortes-Ciriano I, Lee S, Park W-Y, *et al.* A molecular portrait of microsatellite instability across multiple cancers. *Nat Commun* 2017;**8**:15180. doi:10.1038/ncomms15180

23 Harris CR, Millman KJ, van der Walt SJ, *et al.* Array programming with NumPy. *Nature* 2020;**585**:357–62. doi:10.1038/s41586-020-2649-2

24 Virtanen P, Gommers R, Oliphant TE, *et al.* SciPy 1.0: fundamental algorithms for scientific computing in Python. *Nat Methods* 2020;**17**:261–72. doi:10.1038/s41592-019-0686-2

25 scikit-learn: machine learning in Python — scikit-learn 0.23.2 documentation. https://scikit-learn.org/stable/ (accessed 21 Oct2020).
