## Supplementary figures and images for "Evaluating the utility of tumour mutational signatures for identifying hereditary colorectal cancer and polyposis syndrome carriers"

### Supplementary Figure 1

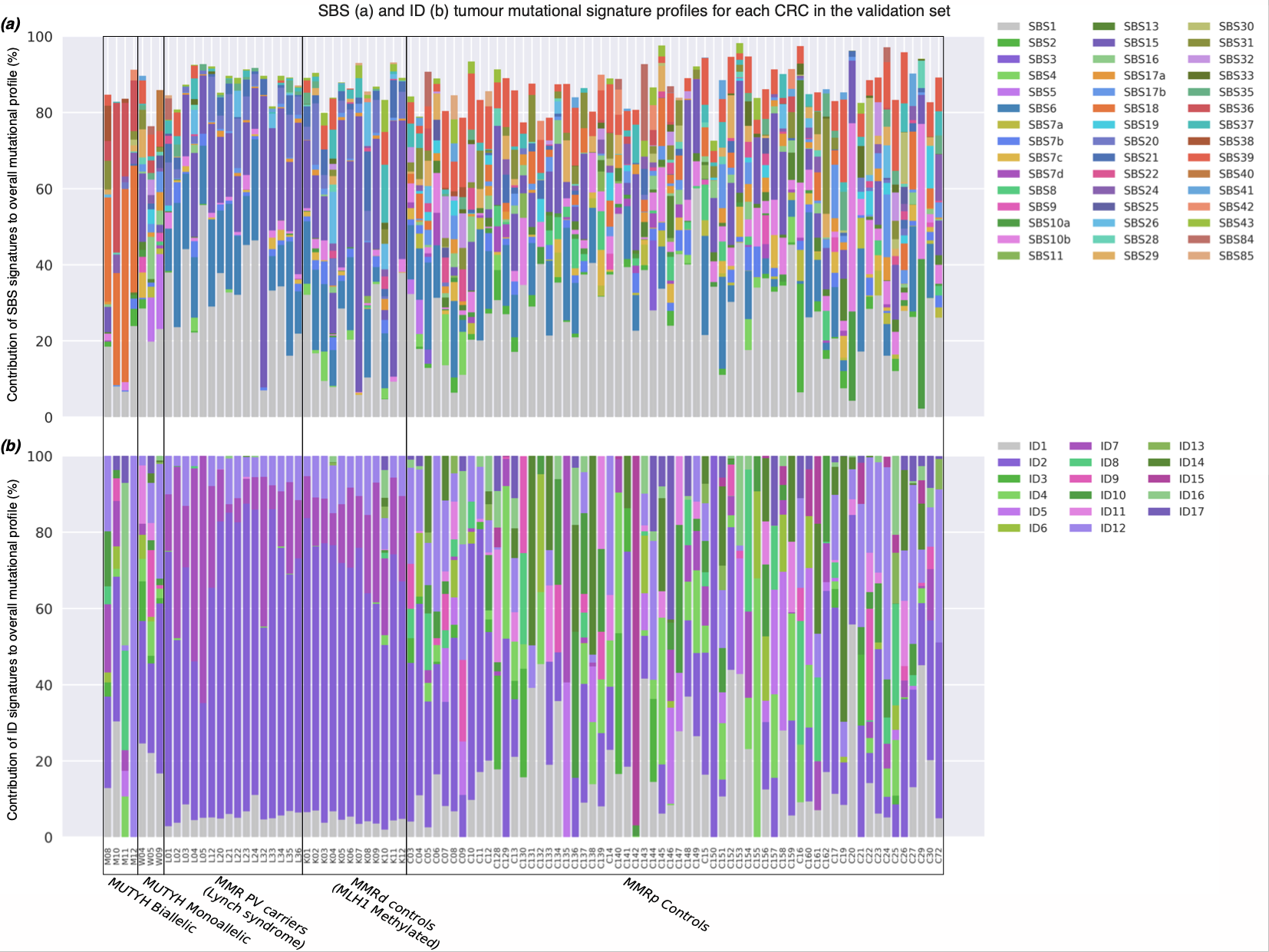

### Supplementary Figure 2

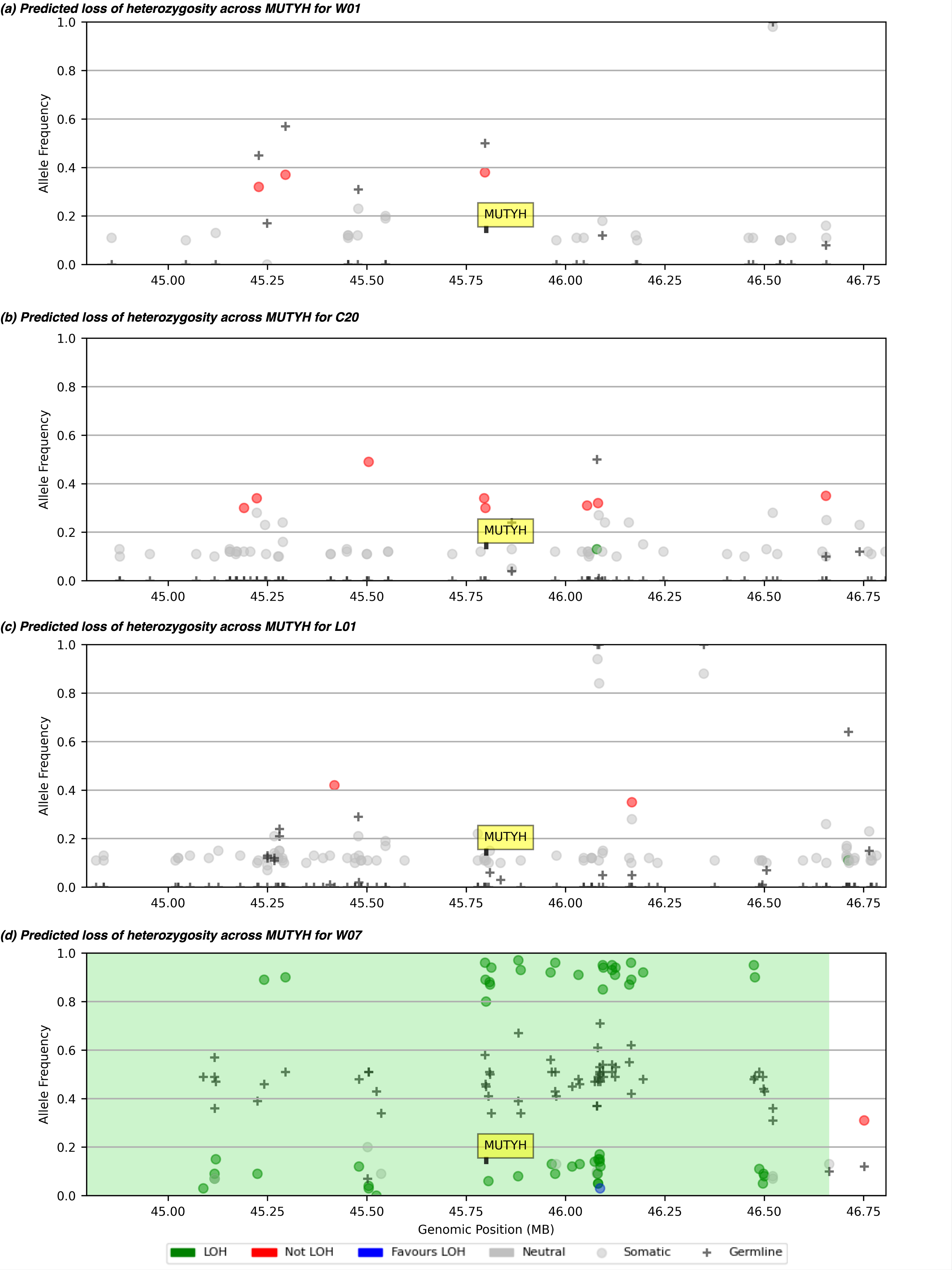
